# Trends in knowledge of HIV status and efficiency of HIV testing services in Sub-Saharan Africa (2000-2020): a modelling study of survey and HIV testing program data

**DOI:** 10.1101/2020.10.20.20216283

**Authors:** Katia Giguère, Jeffrey W. Eaton, Kimberly Marsh, Leigh F. Johnson, Cheryl C. Johnson, Eboi Ehui, Andreas Jahn, Ian Wanyeki, Francisco Mbofana, Fidèle Bakiono, Mary Mahy, Mathieu Maheu-Giroux

## Abstract

**Background:** Knowledge of HIV status (KOS) among people living with HIV (PLHIV) is essential for an effective national HIV response. This study estimates progress and gaps in reaching the UNAIDS 2020 target of 90% KOS, and the efficiency of HIV testing services (HTS) in sub-Saharan Africa (SSA), where two thirds of all PLHIV live.

**Methods:** We used data from 183 population-based surveys (N=2.7 million participants) and national HTS programs (N=315 country-years) from 40 countries as inputs into a mathematical model to examine trends in KOS among PLHIV, median time from HIV infection to diagnosis, HIV testing positivity, and proportion of new diagnoses among all positive tests, adjusting for retesting.

**Findings:** Across SSA, KOS steadily increased from 6% (95% credible interval [95%CrI]: 5% to 7%) in 2000 to 84% (95%CrI: 82% to 86%) in 2020. Twelve countries and one region, Southern Africa, reached the 90% target. In 2020, KOS was lower among men (79%) than women (87%) across SSA. PLHIV aged 15-24 years were the least likely to know their status (65%), but the largest gap in terms of absolute numbers was among men aged 35-49 years, with over 700,000 left undiagnosed. As KOS increased from 2000 to 2020, the median time to diagnosis decreased from 10 to 3 years, HIV testing positivity declined from 9% to 3%, and the proportion of first-time diagnoses among all positive tests dropped from 89% to 42%.

**Interpretation:** On the path towards the next UNAIDS target of 95% diagnostic coverage by 2030, and in a context of declining positivity and yield of first-time diagnoses, we need to focus on addressing disparities in KOS. Increasing KOS and treatment coverage among older men could be critical to reduce HIV incidence among women in SSA, and by extension, reducing mother-to-child transmission.

## Research in Context

### Evidence before this study

One of the major health policy objective of the last decade in the global HIV response has been the adoption of targets to end the AIDS epidemic by 2030. UNAIDS and its partners put forth in 2014 the 90-90-90 objective to increase HIV diagnosis, treatment, and viral load suppression. They called for 90% of all people living with HIV to have knowledge of their HIV status (KOS) by 2020. There is clear evidence of increases in treatment coverage in sub-Saharan Africa (SSA), but little attention has been devoted to the “*first 90*” and trends in KOS have not been systematically reviewed and compared.

We searched PubMed from inception to March 2020 without language restriction with the terms “HIV”[Title/Abstract] AND (“test*”[Title/Abstract] OR “diagnos*”[Title/Abstract] OR “knowledge”[Title/Abstract]) AND (“Africa”[MeSH] OR “Africa”[Title/Abstract]), the websites from the *Joint United Nations Programme on HIV/AIDS* (UNAIDS), and the *World Health Organization* for HIV testing reports and guidelines. Several studies and reports present KOS estimates for selected countries but none comprehensively examined KOS trends by country, age, and sex, or provided estimates of HIV testing services (HTS) efficiency.

### Added value of this study

Due to incomplete HIV surveillance data, and to non-disclosure of HIV positive status in most population-based surveys, assessment of KOS is challenging and not uniform in SSA. By triangulating household survey data about the proportion of adults ever tested for HIV and HTS program data on the total annual number of HIV tests performed among adults using a mathematical model of testing behaviors, this study is the first to systematically and comprehensively assess how KOS and HTS efficiency evolved in SSA over 20 years, and with stratification by sex, age, and region.

### Implications of the available evidence

The last two decades witnessed remarkable increases in KOS across SSA, but stark sex, age, and regional disparities remain, even in countries that have met the 90% target overall. Concomitant decreases in median time to diagnosis, HIV testing positivity, and proportion of new diagnoses among all positive tests highlight one of the major challenges faced by testing programs – targeting of HTS to achieve greatest yield of new diagnoses as the undiagnosed population shrinks and diagnosis delays are reduced. With national HIV control programs now contemplating how to reach the next UNAIDS target of 95% diagnostic coverage by 2030, there is a need to focus on addressing disparities in KOS and to better understand retesting patterns.

## Introduction

Efficient and effective HIV testing services (HTS) are a key component to efforts to end the AIDS epidemic. A positive diagnosis enables people living with HIV (PLHIV) to receive life-saving antiretroviral therapy (ART)^1^ and, for pregnant women living with HIV, risk of mother-to-child HIV transmission can be almost entirely prevented.^2^ At the population level, early diagnosis and treatment could reduce incidence by dramatically lowering viremia such that those with a suppressed viral load are unable to contribute to onward transmission.^3^ HTS also helps identify people who are vulnerable to HIV acquisition and link them to effective HIV prevention services (e.g., voluntary male medical circumcision, pre-exposure prophylaxis).^4^

In sub-Saharan Africa (SSA), where more than two-thirds of PLHIV reside,^5^ HTS were initially provided through voluntary counselling and testing upon request in stand-alone sites.^6^ As HIV treatment became more widely available, provider-initiated HIV testing and counselling emerged, expanding HIV testing services to all patients in health facilities. HTS was also integrated into antenatal care, which greatly increased testing coverage among pregnant and postpartum women.^6^ Such facility-based services were gradually expanded and implementation of community-based services enabled underserved rural and marginalized key populations to be reached by HTS and treatment.^7-9^ The development of new testing technologies and strategies — including point-of-care rapid diagnostic tests, self-testing, partner testing and home-based testing — provided opportunities to accelerate delivery of results and linkage to care.^10^

Recognizing the individual and population benefits of HIV testing and treatment, in 2014, the *Joint United Nations Program for HIV/AIDS* (UNAIDS) proposed ambitious targets to strengthen the HIV treatment and care cascade such that, by 2020, 90% of PLHIV know their status, 90% of those diagnosed receive ART, and 90% of those treated have a suppressed viral load; with each target increasing to 95% by 2030.^11^ These targets are widely adopted globally, and have motivated shifts in the delivery of HTS, especially in SSA countries with the greatest epidemic burden. Countries monitor and report annually to UNAIDS their progress towards these targets.

However, the proportion of PLHIV who know their status is particularly challenging to monitor in SSA because neither the number of PLHIV, nor the number who are diagnosed, are directly counted. Estimates for PLHIV typically come from mathematical models synthesising HIV serosurvey and antenatal testing data ̲ for example, the UNAIDS-supported Spectrum model.^12^ Aggregate HTS data including the number of HIV tests conducted and number of HIV diagnoses, are routinely collected, but reports are often not deduplicated and rates of retesting and re-diagnosis can be high.^13,14^ Household surveys provide cross-sectional data about testing history by HIV status at intervals roughly every five years in most countries, but only a few surveys directly ask respondents if they are aware of their HIV status, a sensitive question that has high potential for non-disclosure.^15-17^ These challenges are compounded by imprecise estimates for the number of new infections by age, sex and geographical area, and by incomplete ascertainment of mortality among the previously diagnosed and undiagnosed population.

Nearing the UNAIDS’ interim 2020 target deadline, we sought to evaluate progress towards the ‘first 90’ HIV diagnosis target in SSA, describe the impact of HTS programs on knowledge of HIV positive status (KOS) and timeliness of HIV diagnosis over the 2000-2020 period, and identify remaining gaps in who is being reached by HTS. We synthesized data from 40 SSA countries about HIV testing history from population-based surveys, HTS program data, and HIV epidemic indicators using a validated mathematical model specifically designed to estimate KOS.^13^ In addition to trends in KOS and diagnosis gaps, we estimated time from HIV infection to diagnosis, probability of getting tested within one year of infection or before reaching a CD4 cell count threshold lower than 350 CD4 cells per µL, positivity (proportion of HIV-positive tests among all tests), diagnosis yield (proportion of new diagnoses among all tests), and proportion of new diagnoses among positive tests.

## Methods

### Overview

We previously developed and validated a compartmental deterministic mathematical model (named *Shiny90*), to synthesize multiple data sources into a coherent framework to longitudinally estimate KOS. This model has been described in detail elsewhere.^13^ Briefly, *Shiny90* models the transition of individuals aged ≥15 years between six stages: 1) HIV-susceptible who have never been tested, 2) HIV-susceptible ever tested, 3) PLHIV who have never been tested, 4) PLHIV unaware who have ever been tested, 5) PLHIV aware (not on ART), and 6) PLHIV on ART. Household surveys and HTS program data are used to estimate the rates of HIV testing among adults not living with HIV and those living with HIV, where HIV testing rates vary with calendar time, sex, age, previous HIV testing status, awareness of status, and, for PLHIV, CD4 cell count category (as a marker of risk of AIDS-related symptoms motivating care-seeking and HIV testing).^13^ In this way, the proportion of PLHIV who know their status estimated by *Shiny90* is bound by ART coverage (minimum) and the proportion of PLHIV who have ever been tested and received the results (maximum).

### Data sources and model calibration

*Shiny90* uses inputs for HIV incidence, mortality, and ART coverage estimated and reported by national governments using the UNAIDS-supported *Spectrum* modeling software and its *Estimation and Projection Package*.^12^ The *Spectrum* model calculates epidemic statistics stratified by age, sex, CD4 cell count category, and ART status. Parameter estimates for HIV disease progression and mortality, as well as demographic rates, are also informed by *Spectrum*.

Two main data sources are used for estimation of HIV testing rates during model calibration:

1. The proportion of individuals (≥15 years old) who self-report having ever been tested for HIV and received the result of the last HIV test from national household surveys conducted between 2000-2019. Estimates were stratified by sex, age (15-24, 25-34, and 35-49 years old), and, if available, HIV sero-status. Sources of national household surveys included *Demographic and Health Surveys* (DHS; https://dhsprogram.com/Data/), *AIDS Indicator Surveys* (AIS), *Multiple Indicator Cluster Surveys* (MICS; www.mics.unicef.org/surveys), *Population-based HIV Impact Assessments* (PHIA; https://phia-data.icap.columbia.edu/files), and other country-specific surveys (Figure 1). The model was calibrated to data on the proportion ever tested for HIV, but we did not calibrate to self-reported awareness of status data, due to evidence of non-disclosure.^15-17^
2. Data on the total annual number of HIV tests performed among individuals aged ≥15 years and, where available, total number of positive HIV tests (2000-2019) reported by national HIV testing programs. HTS program data are particularly informative about changes in testing levels after the most recent available population-based survey.^13^

**Figure 1:**
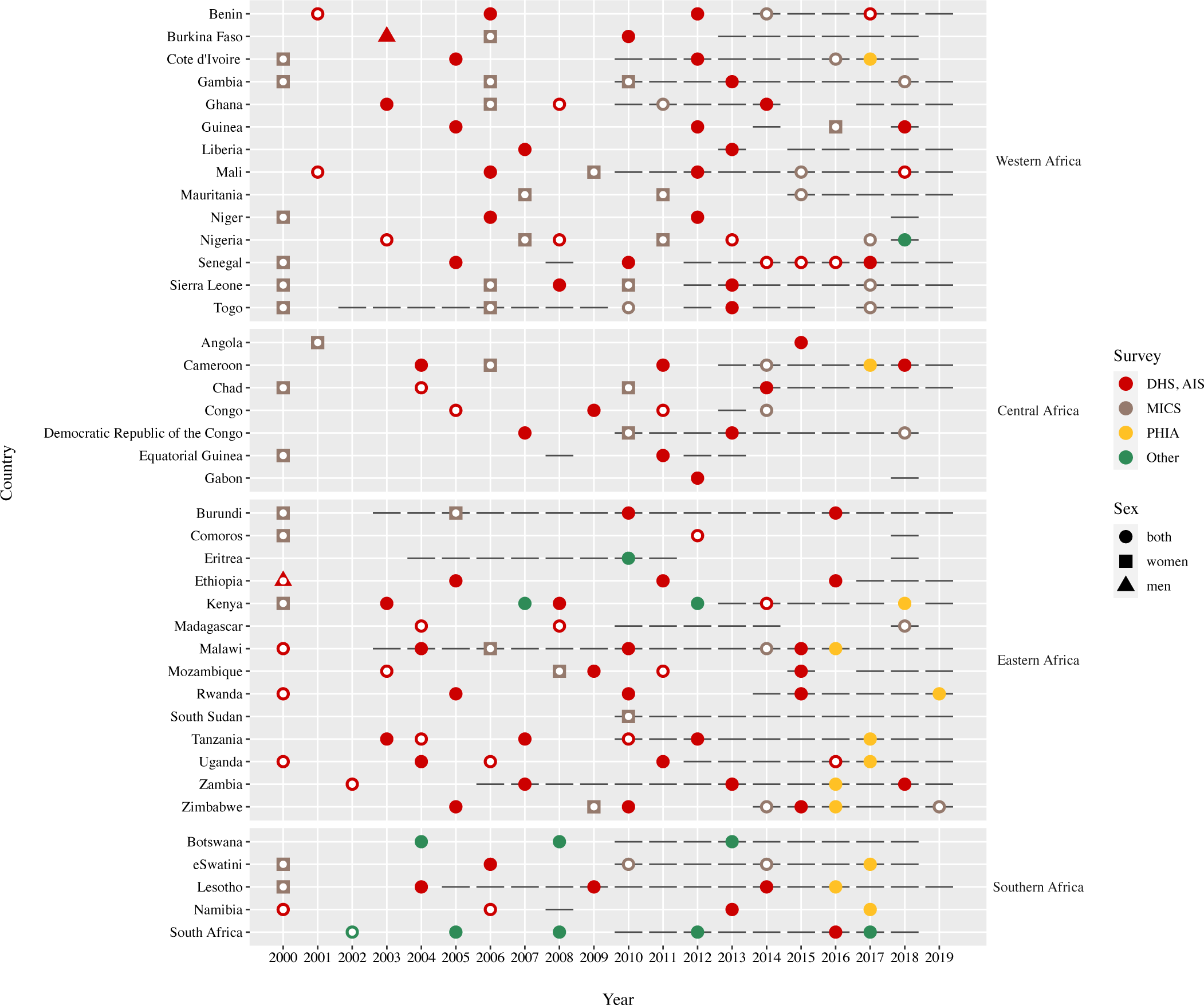
Summary of included surveys and HIV testing services program data by country and year, 2000-2019. Circles represent surveys that were conducted among both men and women, while squares and triangles represent surveys that were conducted among women or among men only, respectively. White dots indicate surveys where results on the proportion of individuals who self-report having ever been tested for HIV are not available by HIV status. Horizontal lines represent HIV testing services program data. DHS: Demographic Health Survey. AIS: AIDS Indicator Survey; MICS: Multiple Indicator Cluster Survey; PHIA: Population-based HIV Impact Assessment Survey. Other types of surveys include: Population Health Survey from Eritrea, South African National HIV Prevalence, Incidence, Behaviour and Communication Surveys, and Botswanan, Kenyan and Nigerian AIDS Indicator Surveys.

For the analyses reported here, we used the *Shiny90* country files submitted to UNAIDS in 2020 (www.unaids.org/en/dataanalysis/datatools/spectrum-epp), including *Spectrum*, surveys, and program data. Additional programme data sources are listed in appendix (pp 47-50).

Sub-saharan African countries with at least one available survey stratified by HIV sero-status, or countries with surveys not stratified by HIV sero-status but having at least one HTS program data set including total number of positive tests between 2000-2019 were included in analyses. These were the minimal set of survey and HTS program data that were required to calibrate the model for a given country. Countries with a population under 250,000 people, or without available survey data, or with only survey data not stratified by HIV sero-status and no HTS program data between 2000-2019 were excluded from analyses.

For each country, the model estimates rates of HIV testing by sex, age, HIV status, and testing and treatment history were estimated from the household survey and HTS program data in a Bayesian framework. The mode of the posterior distribution was estimated via optimisation with the Broyden-Fletcher-Goldfarb-Shanno algorithm^18^ and the posterior density was approximated via Laplace approximation around the posterior mode.^13^ Conceptually, the HTS program data inform rates of HIV testing in the population, while changes in the proportion ever tested by HIV status, sex, and age, alongside estimates of HIV incidence and mortality, inform the proportion of tests conducted among those being HIV tested or diagnosed for the first time versus repeat testing.^13^

### Estimating knowledge of HIV status, positivity, and yield

Using *Shiny90* we calculated annual (2000-2020) proportions of PLHIV with KOS (% of all PLHIV who have ever tested HIV-positive and are thus aware of their HIV status), positivity (% of all HIV tests that are positive), yield of new diagnoses (% of new diagnoses out of all HIV tests), and the proportion of new diagnoses out of all positive tests. For post-2019 model predictions, rates of HIV testing were assumed to remain constant at their 2019 values, but with amplified uncertainty guided by variation in historical testing rates. These projections were also guided by historical increases in ART coverage, with coverage achieved in 2020 extrapolated from the 2016-2019 rates of ART initiation. No adjustments were made for the possible impact of coronavirus disease 2019 (COVID-19) disruptions. All indicators can be stratified by sex and age group, and aggregated to regional level by weighting each country’s indicator by the number of estimated PLHIV from *Spectrum* for that calendar year.

### Estimating time to diagnosis

From the annual sex-, age-, HIV testing history-, and CD4 cell count-specific testing rates, we calculated several cross-sectional indicators using period life table methods^19^ that account for the competing risk of AIDS-related death. These include: time from HIV infection to diagnosis, probability of getting tested within one year following infection and before reaching a CD4 count threshold lower than 350 cells per µL. These indicators were calculated annually by constructing individual period life tables for each of the 16 baseline strata of sex (men, women), age groups (15-24, 25-34, 35-49, and 50+ years), and HIV testing history (never tested, ever tested). Because the estimates are from period life tables, they reflect the distribution of time to diagnosis if a person who seroconverted in a given year was to experience that year’s HIV testing rates by age and CD4 category for their remaining lifetime. Details of the calculations are presented as supplementary materials (appendix p 1). For each calendar year between 2000 and 2020, we estimated these indicators for the 16 age/sex/testing history strata separately. They were then aggregated to the desired demographic or geographic level (e.g., age, sex, country, region [Western, Central, Eastern, and Southern Africa]) by weighting each stratum by the estimated number of new HIV infections in that stratum for that year (obtained from *Spectrum*).

### Uncertainty

We obtained uncertainty intervals by drawing 1,000 samples from the posterior distribution of the testing rates estimated by *Shiny90*. We summarized all indicators using the median, 2.5^th^ and 97.5^th^ percentile of their posterior distribution. We performed analyses using R version 3.5.1 and the Rcpp packages.^20^ The code for *Shiny90* is available on a public repository (www.github.com/mrc-ide/first90release). We followed the *Guidelines for Accurate and Transparent Health Estimates Reporting* (GATHER, appendix p 51).^21^

### Ethical approval

All analyses were performed on anonymized and deidentified data. All DHS/AIS survey protocols have been approved by the Internal Review Board of ICF International in Calverton (USA) and by the relevant country authorities for other surveys (MICS and PHIA). Further information on the ethics approval can be found in the individual country reports. Ethics approval for secondary data analyses was obtained from McGill University’s Faculty of Medicine Institutional Review Board (A10-E72-17B).

### Role of the Funding Source

The funders of the study played no role in study design, data collection, data analysis, data interpretation, or writing of the report. The corresponding author had full access to all the data in the study and had final responsibility for the decision to submit for publication.

## Results

A total of 40 countries, 183 population-based surveys (>2.7 millions surveyed individuals), and 315 country-years of HTS program data reports informed our model (Figure 1). Four SSA countries (Cabo Verde, Central African Republic, Guinea-Bissau, Mauritius) were excluded from the analyses due to insufficient data inputs for model calibration, and one (Sao Tome and Principe), because of high uncertainty in epidemic statistics for small population sizes (<250,000 people). Results of the *Shiny90* model calibration are presented in Text S2 (appendix pp 2-42).

Across SSA, the proportion of adults 15 years and older (both those living and not living with HIV) estimated to have been tested increased by 48 percentage points from 2000 to 2020 (Table 1). Testing coverage was highest in Southern Africa with 85% (95%CrI: 83 to 88%) of adults projected to have ever been tested in the region in 2020.

**Table 1:** Regional progress in HIV testing related outcomes among adults 15 years and older in sub-Saharan Africa, 2000-2020.

| Outcome | Year | Sub-Saharan Africa |  | Western Africa |  | Central Africa |  | Eastern Africa |  | Southern Africa |  |
| --- | --- | --- | --- | --- | --- | --- | --- | --- | --- | --- | --- |
| Proportion of individuals (≥15 years old) ever tested for HIV among overall population (%) | 2000 | 3.6 | (3.0 to 4.4) | 3.1 | (2.6 to 3.8) | 1.9 | (1.3 to 2.6) | 3.3 | (2.8 to 3.9) | 10 | (8.3 to 13) |
|  | 2005 | 11 | (10 to 12) | 7.2 | (6.6 to 7.8) | 7.8 | (6.6 to 9.2) | 13 | (12 to 14) | 30 | (29 to 32) |
|  | 2010 | 30 | (29 to 32) | 19 | (18 to 19) | 19 | (17 to 22) | 41 | (39 to 42) | 59 | (58 to 61) |
|  | 2015 | 41 | (40 to 42) | 28 | (27 to 29) | 29 | (28 to 30) | 53 | (52 to 54) | 75 | (74 to 75) |
|  | 2020 | 51 | (49 to 54) | 36 | (34 to 39) | 42 | (39 to 48) | 64 | (62 to 66) | 85 | (83 to 88) |
| Proportion of PLHIV who know their HIV status (%) | 2000 | 5.7 | (4.6 to 7.0) | 4.0 | (2.8 to 5.2) | 3.2 | (2.0 to 4.6) | 4.8 | (4.0 to 5.8) | 9.3 | (7.5 to 12) |
|  | 2005 | 20 | (18 to 22) | 10 | (7.7 to 12) | 15 | (12 to 18) | 19 | (17 to 21) | 27 | (25 to 30) |
|  | 2010 | 53 | (50 to 55) | 33 | (28 to 35) | 37 | (32 to 41) | 56 | (53 to 58) | 60 | (58 to 63) |
|  | 2015 | 71 | (69 to 73) | 52 | (48 to 55) | 53 | (47 to 56) | 74 | (72 to 75) | 79 | (77 to 80) |
|  | 2020 | 84 | (82 to 86) | 67 | (65 to 69) | 70 | (64 to 76) | 86 | (85 to 88) | 90 | (88 to 92) |
| Time to diagnosis or AIDS-related death, median (year) | 2000 | 9.6 | (9.1 to 10) | 11 | (10 to 11) | 11 | (10 to 12) | 11 | (10 to 11) | 7.7 | (7.0 to 8.4) |
|  | 2005 | 7.2 | (6.3 to 8.0) | 10 | (9.4 to 11) | 8.2 | (7.1 to 9.3) | 7.3 | (6.3 to 8.2) | 5.9 | (5.1 to 6.6) |
|  | 2010 | 3.6 | (3.2 to 4.1) | 5.5 | (4.9 to 6.5) | 6.1 | (5.2 to 7.3) | 3.2 | (2.8 to 3.6) | 2.8 | (2.5 to 3.1) |
|  | 2015 | 3.0 | (2.7 to 3.4) | 5.2 | (4.5 to 6.0) | 4.7 | (3.9 to 6.0) | 2.6 | (2.4 to 2.9) | 2.2 | (1.9 to 2.4) |
|  | 2020 | 2.6 | (1.8 to 3.5) | 5.4 | (4.1 to 6.5) | 3.9 | (2.2 to 6.0) | 2.0 | (1.5 to 2.7) | 1.5 | (0.9 to 2.3) |
| Probability of getting tested within 1 year following infection (%) | 2000 | 2.6 | (2.1 to 3.3) | 1.6 | (1.1 to 2.1) | 1.5 | (1.0 to 2.1) | 1.7 | (1.4 to 2.1) | 4.2 | (3.3 to 5.3) |
|  | 2005 | 5.9 | (4.5 to 7.7) | 1.8 | (1.1 to 2.6) | 5.1 | (3.6 to 7.1) | 5.8 | (4.4 to 7.6) | 7.5 | (5.8 to 9.7) |
|  | 2010 | 21 | (18 to 25) | 11 | (7.7 to 14) | 9.3 | (6.5 to 13) | 25 | (21 to 29) | 23 | (20 to 26) |
|  | 2015 | 26 | (23 to 30) | 13 | (9.3 to 17) | 12 | (7.8 to 18) | 31 | (28 to 35) | 28 | (25 to 32) |
|  | 2020 | 33 | (23 to 46) | 12 | (7.0 to 19) | 16 | (7.6 to 34) | 40 | (29 to 52) | 40 | (26 to 60) |
| Probability of getting tested before reaching a CD4 count lower than 350 cells/μL (%) | 2000 | 19 | (16 to 23) | 13 | (9.3 to 16) | 12 | (8.3 to 17) | 14 | (11 to 16) | 30 | (25 to 35) |
|  | 2005 | 35 | (29 to 42) | 14 | (9.2 to 19) | 30 | (22 to 38) | 35 | (29 to 42) | 43 | (37 to 50) |
|  | 2010 | 63 | (58 to 66) | 45 | (36 to 51) | 43 | (34 to 51) | 66 | (62 to 70) | 69 | (66 to 72) |
|  | 2015 | 67 | (63 to 70) | 47 | (38 to 53) | 52 | (41 to 60) | 71 | (68 to 74) | 75 | (72 to 77) |
|  | 2020 | 71 | (62 to 79) | 44 | (32 to 56) | 59 | (39 to 75) | 77 | (70 to 83) | 81 | (72 to 89) |
| Positivity (% of positive tests among all tests) | 2000 | 9.0 | (7.7 to 10) | 3.0 | (2.1 to 3.5) | 4.6 | (3.5 to 5.2) | 11 | (9.6 to 12) | 15 | (13 to 19) |
|  | 2005 | 11 | (9.2 to 14) | 4.0 | (3.3 to 4.5) | 5.4 | (4.2 to 6.7) | 10 | (9.1 to 12) | 20 | (16 to 26) |
|  | 2010 | 5.9 | (4.3 to 8.3) | 2.6 | (2.0 to 3.2) | 5.0 | (3.6 to 6.9) | 5.6 | (4.2 to 7.5) | 13 | (9.0 to 22) |
|  | 2015 | 4.3 | (3.5 to 5.2) | 2.2 | (1.7 to 2.7) | 3.4 | (2.4 to 4.7) | 4.0 | (3.5 to 4.6) | 9.2 | (7.1 to 13) |
|  | 2020 | 2.8 | (2.1 to 3.9) | 1.9 | (1.3 to 2.7) | 2.2 | (1.4 to 3.3) | 2.5 | (1.9 to 3.3) | 5.5 | (3.8 to 8.4) |
| Diagnosis yield (% of new diagnoses among all tests) | 2000 | 7.9 | (7.0 to 8.6) | 2.6 | (1.8 to 2.9) | 4.2 | (3.2 to 4.7) | 9.7 | (8.8 to 11) | 13 | (12 to 14) |
|  | 2005 | 7.8 | (7.0 to 8.4) | 3.2 | (2.7 to 3.6) | 3.8 | (3.1 to 4.3) | 7.6 | (7.0 to 8.1) | 14 | (12 to 15) |
|  | 2010 | 2.8 | (2.4 to 3.3) | 1.5 | (1.2 to 1.7) | 2.6 | (2.1 to 3.0) | 2.4 | (2.1 to 2.9) | 6.9 | (6.2 to 7.5) |
|  | 2015 | 1.9 | (1.7 to 2.1) | 1.1 | (0.9 to 1.3) | 1.6 | (1.2 to 1.8) | 1.7 | (1.6 to 1.8) | 4.4 | (4.1 to 4.6) |
|  | 2020 | 1.2 | (0.9 to 1.5) | 1.0 | (0.7 to 1.5) | 0.9 | (0.6 to 1.3) | 1.0 | (0.8 to 1.3) | 2.2 | (1.6 to 2.9) |
| Proportion of new HIV diagnoses among all positive tests (%) | 2000 | 89 | (77 to 96) | 86 | (79 to 94) | 93 | (85 to 97) | 91 | (85 to 95) | 87 | (70 to 97) |
|  | 2005 | 72 | (55 to 86) | 79 | (71 to 90) | 71 | (55 to 86) | 74 | (61 to 84) | 70 | (48 to 87) |
|  | 2010 | 48 | (33 to 65) | 56 | (46 to 73) | 52 | (36 to 71) | 44 | (32 to 59) | 53 | (31 to 76) |
|  | 2015 | 44 | (37 to 53) | 48 | (38 to 61) | 46 | (33 to 59) | 42 | (37 to 48) | 47 | (34 to 60) |
|  | 2020 | 42 | (30 to 55) | 52 | (38 to 68) | 42 | (24 to 57) | 41 | (30 to 52) | 39 | (25 to 55) |

PLHIV: people living with HIV, HTS: HIV testing services. Numbers in parentheses correspond to 95% credible intervals

The proportion of adult PLHIV with KOS increased steadily from 5.7% (95%CrI: 4.6 to 7.0%) in 2000 to 84% (95%CrI: 82 to 86%) in 2020 in SSA (Table 1). While KOS increased dramatically in all four SSA regions, KOS was consistently lower in Western and Central Africa as compared to Eastern and Southern Africa (Figure 2A; Figure S1: appendix p 43). Within the regions, national estimates were also highly heterogeneous, especially in Eastern Africa with a 77 percentage point difference between the countries with the lowest and highest KOS estimates. Overall, we projected that 12 countries (Figure 3) and one region, Southern Africa, will have reached at least 90% KOS in 2020. Countries with higher KOS tended to be those in which the annual number of tests relative to the total population aged ≥15 years was highest (Figure S2: appendix p 44).

**Figure 2:**
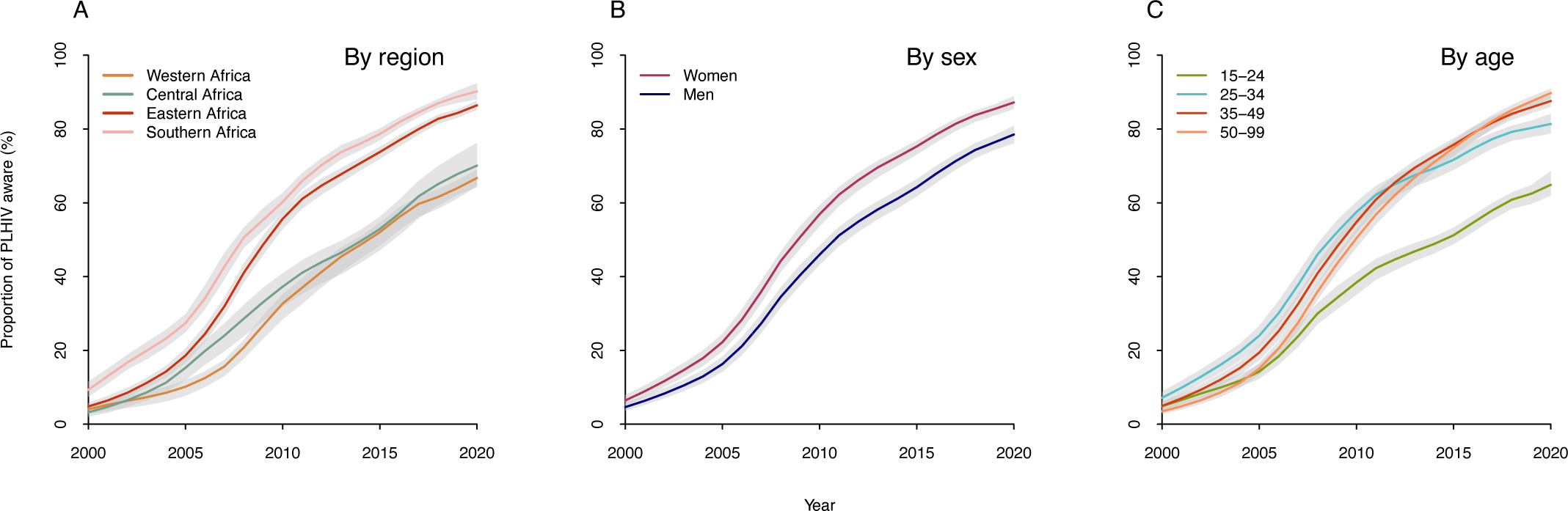
Progress and disparities in knowledge of HIV status in sub-Saharan Africa, 2000-2020. Panels A to C show trends in proportion of people living with HIV (PLHIV) who are aware of their HIV status in sub-Saharan Africa by region (A), by sex (B), or by age group (C). The shaded areas correspond to the 95% credible intervals.

**Figure 3:**
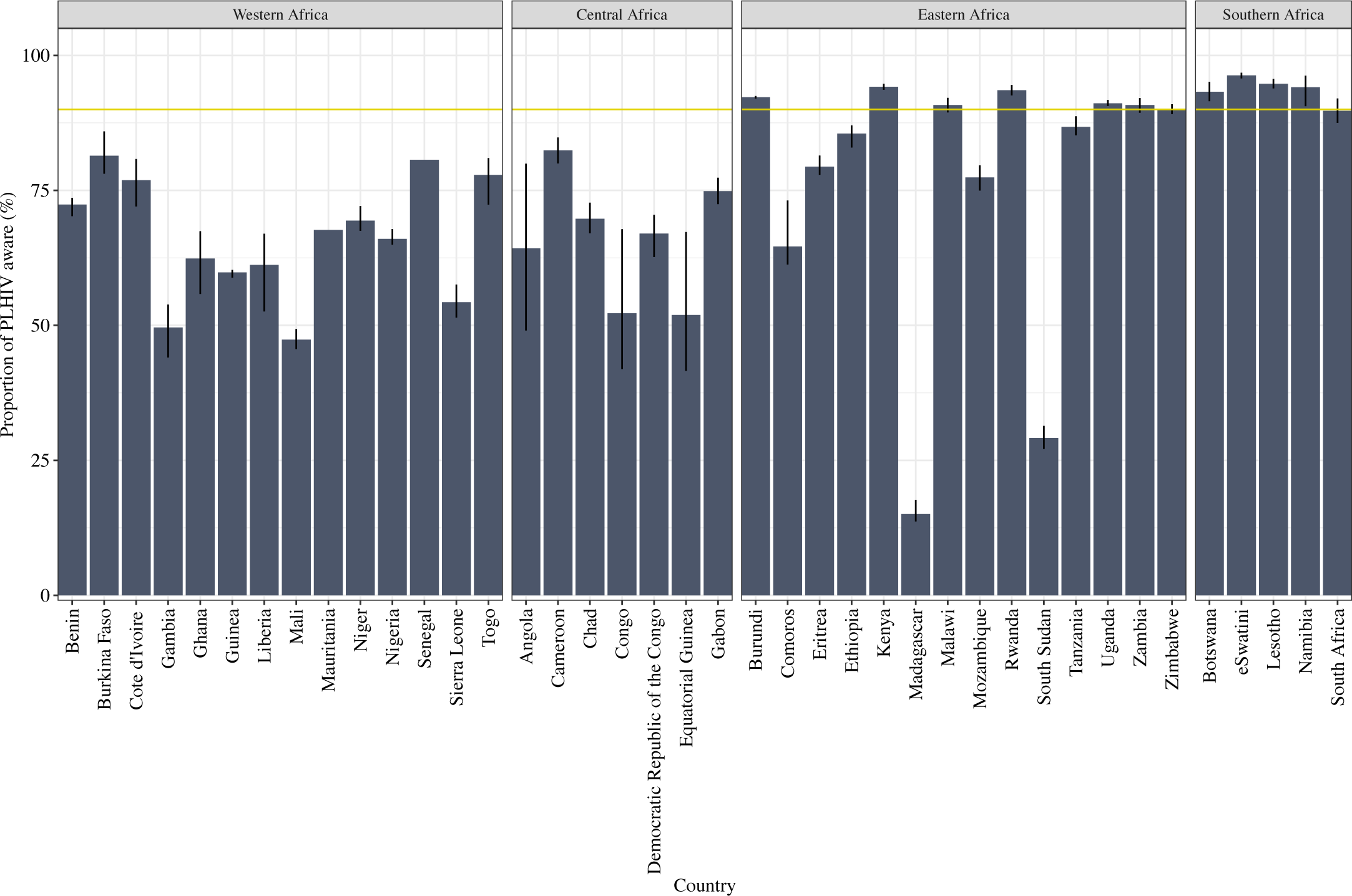
National estimates of knowledge of HIV status in sub-Saharan Africa, 2020. Proportion of PLHIV who know their HIV status. The horizontal yellow line represents a threshold of 90% and vertical lines correspond to the 95% credible intervals.

Our results also suggest disparities in KOS by sex and age. Across SSA in 2020, men had lower KOS (79%, 95%CrI: 76 to 81%) than women (87%, 95%CrI: 85 to 89%), and 15-24 year-olds were the least likely to know their status (65%, 95%CrI: 62 to 69%; Figure 2B-C, Table S4: appendix p 53). Such disparities were also observed among the 12 countries projected to achieve at least 90% of KOS overall in 2020. Of these countries, only six are projected to achieve 90% of KOS among men, and none are projected to do so among the 15-24 year-olds.

While the proportion of PLHIV aware of their status was lower among younger adults, the absolute number of PLHIV was also lower. Consequently, in absolute numbers, the largest group of undiagnosed PLHIV in SSA were men aged 35-49 years, with >700,000 left undiagnosed and 305,000 diagnoses needed to reach 90% awareness of status (Figure 4, Table S3: appendix p 52).

**Figure 4:**
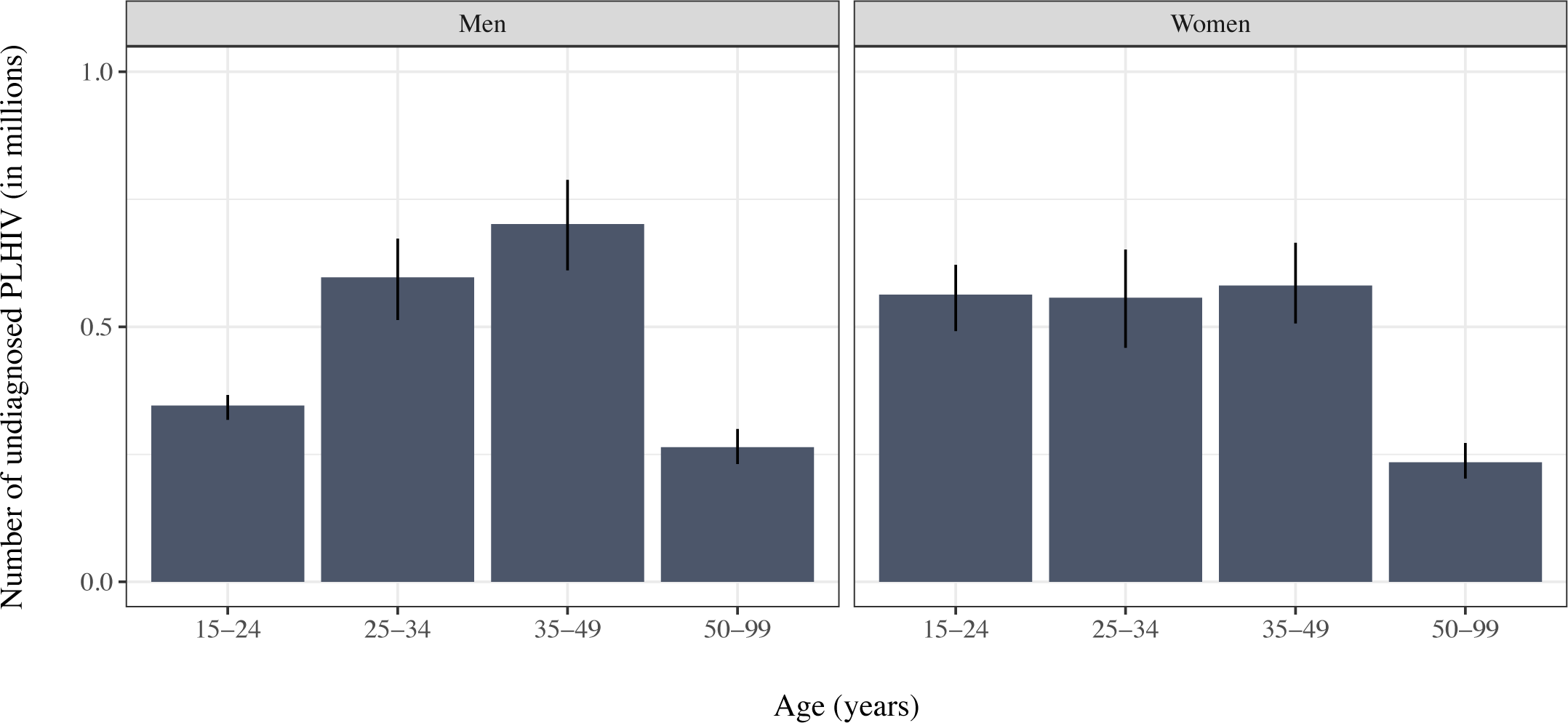
Absolute diagnosis gaps in sub-Saharan Africa, 2020. Each bar shows the total number of undiagnosed people living with HIV (PLHIV) by sex- and age-stratification. Vertical lines correspond to the 95% credible intervals.

The median time from HIV infection to diagnosis (or death) decreased by 7 years from 2000 to 2020 for all of SSA (Table 1; Figure 5A). That is, if projected HIV testing rates in 2020 persisted into the future, 50% of people infected in 2020 would be diagnosed (or, with small probability, suffer AIDS-related mortality) within 2.6 years of seroconverting. National trends are presented in Figure S3 (appendix p 45).

**Figure 5:**
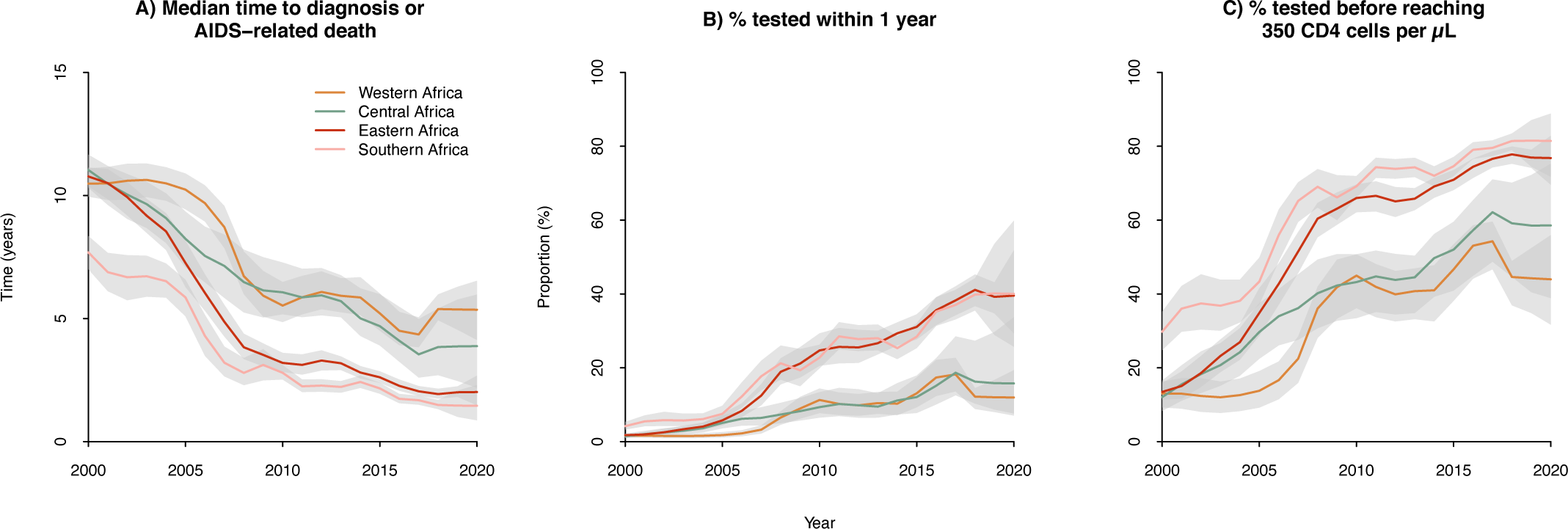
Progress in timeliness of HIV diagnosis in sub-Saharan Africa, 2000-2020. Regional trends in median time to diagnosis or AIDS-related death (A), in the probability to get tested within one year (B) or before reaching a CD4 count threshold lower than 350 cells per µL (C) were assessed through period life-table analyses. The shaded areas correspond to the 95% credible intervals.

Consistent with the estimated decreases in median time to diagnosis, the probability of receiving an HIV test within 1 year following infection or before reaching a CD4 count threshold lower than 350 cells per µL increased respectively by 31 and 52 percentage points from 2000 to 2020 in SSA (Table 1; Figure 5B-C).

The proportion of all HIV tests that are positive (positivity) decreased by 6 percentage points from 2000 to 2020 (Table 1; Figure S4: appendix p 46), and the proportion of new diagnoses among all tests (diagnosis yield) decreased by 7 percentage points. Concommitantly, the proportion of new diagnoses among positive tests decreased by 47 percentage points over the study period (Table 1). That is, we project that 58% of PLHIV undergoing testing in 2020 will have been previously diagnosed with HIV. For each of the previous outcomes, five-yearly estimates are presented by sex and age stratification and by region in Tables S4 to S8 (appendix pp 53-57).

## Discussion

Across SSA, impressive gains were achieved in KOS with 84% (95%CrI: 82 to 86%) of PLHIV being aware of their HIV positive status, and 12 countries and the region of Southern Africa that are projected to reach the 90% KOS target in 2020. Concomitant with these improvements, we estimated that median time from HIV acquisition to diagnosis would be reduced to 2.6 years (95%CrI: 1.8 to 3.5) over that period.

Despite this progress, our results highlight substantial regional, national, sex, and age disparities in KOS in SSA. KOS was consistently lower in Western and Central Africa than Eastern and Southern Africa. In those regions, HIV prevalence is lower but key populations — including sex workers, men who have sex with men, and people who inject drugs — account for a higher HIV burden. For example, they generally represent a small fraction of the population but accounted for 42% of all new HIV infections in 2019 in the region of Western and Central Africa.^5^ Stigma and discrimination towards key populations are common in many health facilities, which may lead to delayed HIV testing, concealment of HIV positive status, and/or poor uptake of HIV services.^22^ A recent systematic review and meta-analysis has shown that, among men who have sex with men in Africa, lower testing and KOS were associated with more hostile legislation, and that KOS remained low in the region.^23^ To improve coverage of HIV health services in Western and Central Africa, antidiscrimination and protective laws to eliminate stigma and discrimination among key populations should be implemented and enforced, health workers trained and sensitized, and key population-friendly services provided.^22^ Eastern Africa, despite having high KOS across the region, includes the two countries with the lowest KOS—South Sudan and Madagascar. While new HIV infections declined overall in Eastern African countries between 2010 and 2019, new infections in South Sudan and Madagascar are estimated to have increased by 17% and 191%, respectively.^5^ This underscores that reducing new HIV infections, ‘*turning off the tap*’ of undiagnosed PLHIV, is key to reaching KOS targets.

In all four SSA regions, and consistent with previous studies, men are less likely to know their HIV status compared to women.^24-28^ Overall, the diagnosis gap is such that there is a 8% point difference between men and women in 2020. Large differences in KOS are also observed between age groups, with the lowest proportion diagnosed being among PLHIV aged 15-24 years. Importantly, all countries have yet to reach 90% KOS in this younger group. This gap between age groups is the natural consequence of HIV transmission dynamics. HIV incidence is highest and average time since infection is short — and thus cumulative exposure to testing is lower — in this age group compared to older ones.^12^ To achieve 90% KOS among 15-24 years-old would require a simultaneous increase in testing with greater investment in HIV prevention to increase coverage of high impact prevention interventions.

While we found that KOS was proportionally the lowest among men aged 15-24 years old, the largest group of undiagnosed PLHIV was men aged 35-49 years, with >700,000 projected to be undiagnosed in 2020. Lower uptake of HIV testing among men may be explained by fewer opportunities for testing as well as other social and system-wide barriers such as harmful gender norms^7,29^ and inaccessible or unfriendly services.^30^ Engaging men in HIV prevention efforts is critically important, not only for their own needs, but also for their sexual partners. An increase in KOS and of treatment coverage among older men could be critical to reduce HIV acquisition rates among women, and by extension, reducing mother-to-child transmission. Among different testing approaches, community-based testing, door-to-door HTS, home-based couples testing, workplace programs, mobile testing services, social network interventions, incentives to test, self-testing, and partner notification have shown success in increasing diagnostic coverage among men.^31^ As part of these efforts, facilitating linkage and retaining men with HIV care remains a key challenge for further progress towards HIV testing and treatment targets.

Despite improvements, especially in Southern Africa where the median time to diagnosis (or AIDS-death) was estimated at 1.5 years (95%CrI: 0.9 to 2.3) in 2020, we projected that across SSA and at current testing levels, 50% of PLHIV will not be diagnosed within 3 years following their infection, and 29% will not get tested before reaching a CD4 count threshold lower than 350 cells per µL in 2020. These diagnostic delays impede rapid ART initiation at high CD4 counts which, in its absence, contribute to increased HIV morbidity and onward HIV transmission.^1, 32-34^ Reducing diagnostic delays on their own will likely not be enough to improve individual and population health outcomes. Earlier diagnosis should be accompanied by rapid ART linkage and long-term adherence to ART — these are crucial to minimizing morbidity and reducing HIV incidence.^32,33,35^

As the undiagnosed population shrinks and diagnosis delays are reduced, targeting of HTS to achieve greatest yield of new diagnoses is one of the major challenges faced by testing programs.^36^ Although we noted an ecological correlation between a country’s testing volume with respect to its population of reproductive age and KOS, we also estimated a decline in positivity and in the yield of new diagnoses. Such declining yields are an inevitable consequence of reaching saturation in testing programs — as long as testing rates are lower in previously-diagnosed individuals than in undiagnosed, we can expect yields to decline as KOS increases. Our analyses also highlight substantial retesting of PLHIV already aware of their status. We projected that 58% of positive tests will be performed on previously diagnosed PLHIV in SSA in 2020. In previous studies conducted in SSA between 2004 and 2018, retesting among PLHIV with known HIV status was also common, ranging from 13% to 68%.^14, 37-41^ Retesting can be motivated by multiple factors, one of them being the ability to confirm the accuracy of the initial test result.^41-43^ Another important driver of retesting may be avoiding disclosing prior knowledge of HIV positive status due to societal stigma or denial. A recent study conducted among persons undergoing HIV testing at a health facility in South Africa found that 50% of patients testing HIV-positive had previously been in HIV care (and hence previously diagnosed). Among these, half did not disclose prior knowledge of HIV status to their health care provider.^14^ Further research is needed to assess the potential benefits of retesting for reengaging PLHIV in care.

This analysis has some limitations. First, *Shiny90* does not provide estimates of diagnosis coverage among <15 years-old, nor can it disaggregate metrics by key population groups. Second, we could have overestimated KOS in some low HIV prevalence countries where key populations are disproportionately affected by HIV if these groups are underrepresented in population-based surveys. Third, uncertainty in the PLHIV denominator, HIV incidence estimates, and ART coverage are not accounted for. This does not affect the validity of point estimates, but their precision could be overestimated. Fourth, we assumed that HIV testing does not result in false negative or false positive results. The assumption of no false negative HIV test result may have slightly over-estimated KOS and probability of getting tested within 1 year or before reaching a CD4 count threshold lower than 350 cells per µL, and under-estimated median time to diagnosis or (AIDS-related death). The number of HIV diagnoses reported in HTS programme data could be inflated if WHO-recommended retesting to verify HIV diagnosis before ART initiation was incorrectly counted as separate HIV diagnoses, which our model would not be able to identify from routinely reported data. Fifth, we also assumed that self-reporting of HIV testing histories was accurate but social desirability and recall biases could result in underestimation of the proportion ever tested and, ultimately, of KOS.^44^ However, validation of self-reported HIV testing histories by mean of antiretroviral biomarkers data from PHIA surveys from eSwatini, Malawi, Tanzania, and Zambia using Bayesian latent class model suggest that, self-reported HIV testing history being highly sensitive, underestimation of the proportion ever tested and of KOS should be low (*Xia et al. preprint*).^45^ Sixth, earlier estimates of diagnosis delays are informed by relatively few population-based survey estimates and HTS program data. Given the cross-sectional nature of these metrics, they could be more sensitive to the elicited model’s prior distributions in early years. Finally, the impact of measures taken to prevent the spread of COVID-19 in some countries could have affected both HIV incidence and HTS.^46^ Such unacounted factors could potentially lead to slightly lower KOS estimates than those projected in 2020, although a notable decrease would be unlikely since already diagnosed PLHIV would remain so.

Although previous studies examined HIV testing uptake or self-reported KOS at community or country level, the present analysis is believed to be the first to systematically and comprehensively assess how HTS efficiency evolved in SSA over 20 years. By using a unified framework to compare HTS metrics, consistency and comparability of results between the different outcomes, countries, and regions is improved. A second strength is the large number of surveys and program data used for triangulation, improving the precision and robustness of our results. Third, in assessing time to diagnosis (or AIDS-related death) and other related metrics, we provide valuable information to help programs optimize HTS efficiency.^47^ With clear individual and population-health benefits of early treatment initiation, reducing diagnostic delays and improving linkage to care will contribute towards the ultimate goal to end AIDS epidemics by 2030.

## Conclusion

In 2014, the world adopted the goal of achieving 90% HIV diagnosis by 2020. Sub-Saharan Africa, the most affected region, is close to reaching this target and we project that 12 countries and one region, Southern Africa, will have reached that goal among adults in 2020. However, reaching 90% diagnosis coverage remains challenging and our results shed light on stark sex and age gaps in KOS. None of the 12 countries projected to reach the 90% target overall are projected to do so in all age and sex groups. National HIV control programs are now contemplating how to reach the next UNAIDS target of 95% diagnostic coverage by 2030 in a context of declining positivity, declining yields of “true” new diagnoses, and COVID-19 disruption. Reaching this objective will require a better understanding of retesting patterns and a focus on addressing disparities among older men and young people in KOS.

## Data Availability

For the analyses reported here, we used the Shiny90 country files submitted to UNAIDS in 2020 (www.unaids.org/en/dataanalysis/datatools/spectrum-epp), including Spectrum, surveys, and program data. Sources of national household surveys included Demographic and Health Surveys (DHS; https://dhsprogram.com/Data/), AIDS Indicator Surveys (AIS), Multiple Indicator Cluster Surveys (MICS; www.mics.unicef.org/surveys), Population-based HIV Impact Assessments (PHIA; https://phia-data.icap.columbia.edu/files), and other country-specific surveys (Figure 1).

http://www.mics.unicef.org/surveys

https://phia-data.icap.columbia.edu/files

## Contributors

JWE, KG, KM, LFJ, and MMG conceived the study. AJ, JWE, KM, LFJ, and MMG developed the mathematical model. KG performed the analyses. AJ, CCJ, EE, FB, FM, IW, KM, and MM contributed data and helped with result interpretation. KG and MMG wrote the initial draft. AJ, CCJ, EE, FB, FM, IW, KM, LFJ, MM, and IW provided expert input to inform background, context, and local epidemiology. All authors contributed to and approved the final manuscript.

## Declaration of interests

We acknowledge funding from the *Steinberg Fund for Interdisciplinary Global Health Research* (McGill University), the *Canadian Institutes of Health Research*, and the *Bill and Melinda Gates Foundation*. MMG holds a *Canada Research Chair* (Tier 2) in *Population Health Modeling* and reports other from UNAIDS, other from WHO, grants from Gilead Sciences Inc., outside the submitted work; KG reports a Postdoctoral Fellowship from *Fonds de recherche du Québec – Santé*, during the conduct of the study; personal fees from UNAIDS, outside the submitted work; JWE reports grands from *Bill and Melinda Gates Foundation*, grants from UNAIDS, grants from UK Medical Research Council, during the conduct of the study; grants from NIH, grants from UNAIDS, grants from WHO, personal fees from WHO, grants from USAID, outside the submitted work; All other authors has nothing to disclose. The contents in this article are those of the authors and do not necessarily reflect the view of the *World Health Organization*.

