## Appendix for "Trends in knowledge of HIV status and efficiency of HIV testing services in Sub-Saharan Africa (2000-2020): a modelling study of survey and HIV testing program data"

#### Supplemental Material

***Text S1.* Estimating diagnosis delays**

From the annual sex- , age- , HIV testing history- , and CD4 cell count-specific testing rates, we calculated several cross-sectional indicators using period life table methods[17] that account for the competing risk of AIDS-related death. For each stratum, we proceed by initializing the life table at infection (time 0: $t_0$) using the age-specific CD4 cell count distribution at infection (either ≥500 or 350-499 CD4 cells per µL) as is used in Spectrum. We then calculate the number of individuals that will either receive a positive test result or die of AIDS-related cause at each time step ($\Delta t = 0.1$ year), accounting for CD4 cell count progression. As these are period life tables, estimates reflect the distribution of time to diagnosis if a person who seroconverted in year y experienced the HIV testing rates by age and CD4 category estimated for the that year for their remaining lifetime. Specifically, the life tables are constructed using the following equations (for $t \geq t_0$):

$$I_{y(t+\Delta t)}^{kisu} = \begin{cases} I_{yt}^{kisu} - \left(\left(\tau_y^{ki_{[t]}s_{[t]}u} + \epsilon_{is_{[t]}} + \gamma^{is}\right) I_{yt}^{kisu}\right)\Delta t; \quad if\ s = 1 \\ I_{yt}^{kisu} + \left(\gamma^{i(s-1)} I_{yt}^{ki(s-1)u} - \left(\tau_y^{ki_{[t]}s_{[t]}u} + \epsilon_{is_{[t]}} + \gamma^{is}\right) I_{yt}^{kisu}\right)\Delta t; \quad if\ 1 < s < 7 \\ I_{yt}^{kisu} + \left(\gamma^{i(s-1)} I_{yt}^{ki(s-1)u} - \left(\tau_y^{ki_{[t]}s_{[t]}u} + \epsilon_{is_{[t]}}\right) I_{yt}^{kisu}\right)\Delta t; \quad if\ s = 7 \end{cases}$$

where $I_{yt}^{kisu}$ is the number of individuals who seroconverted during calendar year y, from sex k, age i, CD4 cell count s (where 1 indexes CD4 counts of ≥500 cells per µL, 2 for 350-499, 3 for 250-349, 4 for 200-249, 5 for 100-199, 6 for 50-99, and 7 for <50 CD4 cells per µL), and testing history u, at time t since infection; $\gamma^{is}$ is the CD4 cell count progression rates; $\tau_y^{ki_{[t]}s_{[t]}u}$ is the HIV testing rate specific to sex k, age ($i_{[t]}$, accounting for aging as a function of time t), CD4 cell count ($s_{[t]}$, as CD4 cell count decreases as a function of time t), and testing history (u) that were estimated for the calendar year y in which an individual seroconverted; and $\epsilon_{is_{[t]}}$ is the age- (at infection) and CD4 cell count-specific AIDS-related mortality rate.

After constructing the life table, we calculated the median time between HIV infection and diagnosis or AIDS-related death ($m_y^{kiu}$) by finding the time t after which more than half of the newly seroconverted individuals had either been diagnosed or died of HIV ($d_y^{kiu}(t)$):

$$d_y^{kiu}(t) = \frac{\sum_{t_0}^{t} \sum_s \left(\left(\left(\tau_y^{ki_{[t]}s_{[t]}u} + \epsilon_{is_{[t]}}\right) I_{yt}^{kisu}\right)\Delta t\right)}{\sum_s I_{yt_0}^{kisu}}$$

$$m_y^{kiu} = \min_{t>0}\left\{\left|d_y^{kiu}(t) - 0.5\right|\right\}$$

Finally, the probability of getting tested within one year of infection ($p_y^{kiu}$), and of getting tested before reaching a CD4 cell count threshold lower than 350 cells per µL (s=3) ($c_y^{kiu}$) were estimated by accounting for the competing risk of AIDS-related death using, respectively:

$$p_y^{kiu} = \frac{\sum_{t_0}^{1} \sum_s \left(\tau_y^{ki_{[t]}s_{[t]}u} I_{yt}^{kisu}\Delta t\right)}{\sum_s I_{yt_0}^{kisu}}$$

$$c_y^{kiu} = \frac{\sum_s^3 \sum_t \left(\tau_y^{ki_{[t]}s_{[t]}u} I_{yt}^{kisu}\Delta t\right)}{\sum_s I_{yt_0}^{kisu}}$$

For each calendar year between 2000 and 2020, we estimated these indicators for the 16 age/sex/testing history strata separately. They were then aggregated to the desired level of aggregation (e.g., age, sex, country, region) by weighting each stratum by the estimated number of new HIV infections in that stratum for that year (obtained from Spectrum).

***Text S2.* Comparison of calibrated *Shiny90* model fits with programmatic and survey data for each of the 40 included sub-Saharan countries, 2000-2020**

For each country, *Shiny90* posterior estimates of the total number of HIV tests (A), the total number of positive tests (B), the proportion reporting having ever been tested among the HIV negative population (C), among people living with HIV (PLHIV; D), and overall (E) are compared to available HTS program and survey data. On all graphs, the lines correspond to the median of the posterior estimates, whereas the shaded areas correspond to the 95% credible intervals. Black dots on graphs A and B correspond to the HIV testing services program data for the overall number of tests, and for the number of positive tests, respectively. The points on the graphs C to E represent the survey estimates of the proportion of men (circles) and women (squares) reporting having ever been tested among the HIV negative population (C), the PLHIV (D), and overall (E). The lines crossing the points correspond to the 95% confidence intervals of the surveys estimates. For countries denoted by *, surveys for which datasets are not available but are included in knowledge of status estimates reported to UNAIDS are not plotted.

**Contents**


Angola 3
Benin 4
Botswana 5
Burkina Faso 6
Burundi 7
Cameroon* 8
Chad 9
Comoros 10
Congo 11
Cote d'Ivoire* 12
Democratic Republic of the Congo 13
Equatorial Guinea 14
Eritrea 15
eSwatini 16
Ethiopia 17
Gabon 18
Gambia 19
Ghana 20
Guinea 21
Kenya* 22
Lesotho* 23
Liberia 24
Madagascar 25
Malawi 26
Mali 27
Mauritania 28
Mozambique 29
Namibia* 30
Niger 31
Nigeria 32
Rwanda* 33
Senegal 34
Sierra Leone 35
South Africa 36
South Sudan 37
Tanzania 38
Togo 39
Uganda* 40
Zambia 41
Zimbabwe* 42

### Angola

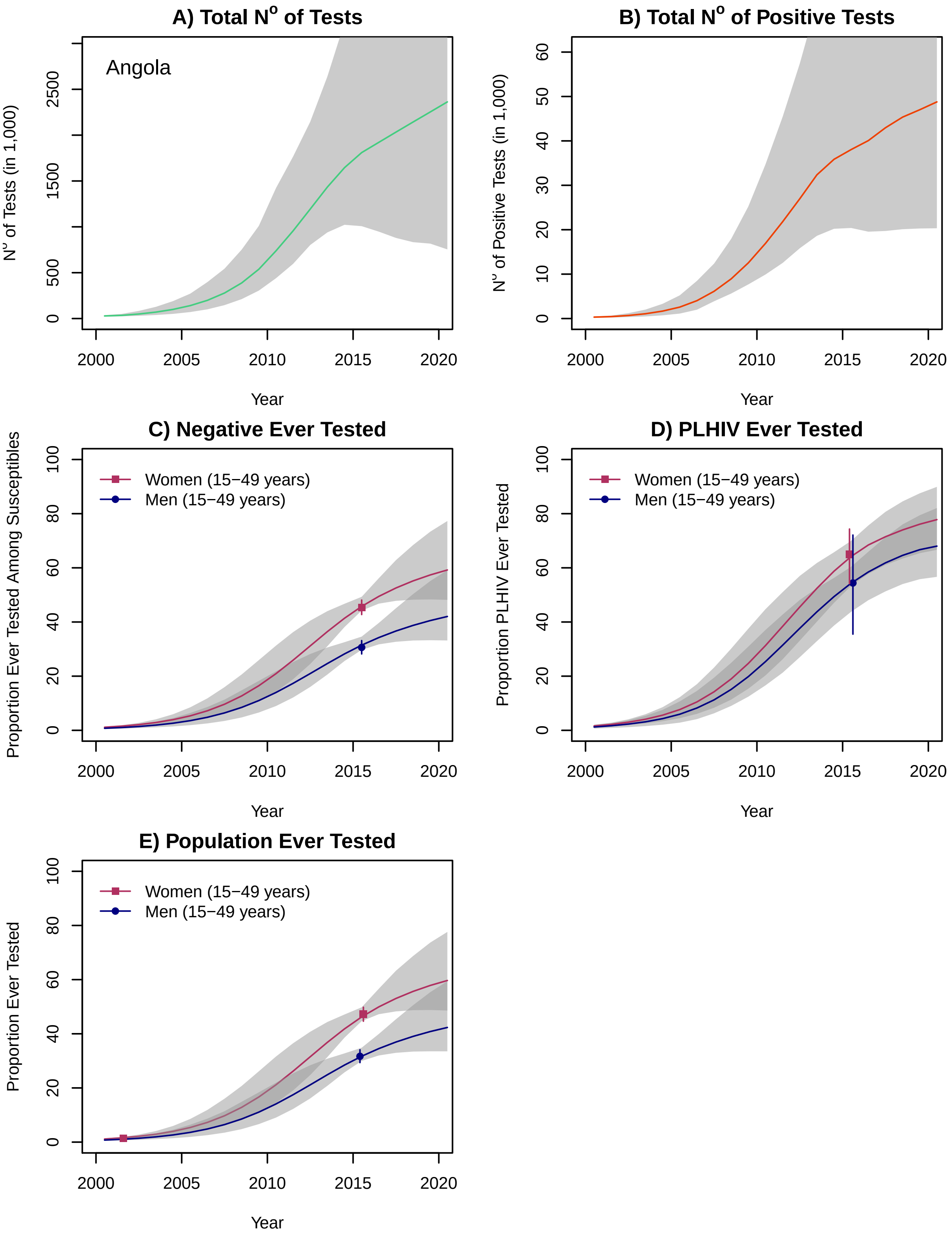
A) Total N$^o$ of Tests
Angola
N$^o$ of Tests (in 1,000)
0
500
1500
2500
2000
2005
2010
2015
2020
Year
B) Total N$^o$ of Positive Tests
N$^o$ of Positive Tests (in 1,000)
0
10
20
30
40
50
60
2000
2005
2010
2015
2020
Year
C) Negative Ever Tested
Proportion Ever Tested Among Susceptibles
Women (15−49 years)
Men (15−49 years)
0
20
40
60
80
100
2000
2005
2010
2015
2020
Year
D) PLHIV Ever Tested
Proportion PLHIV Ever Tested
Women (15−49 years)
Men (15−49 years)
0
20
40
60
80
100
2000
2005
2010
2015
2020
Year
E) Population Ever Tested
Proportion Ever Tested
Women (15−49 years)
Men (15−49 years)
0
20
40
60
80
100
2000
2005
2010
2015
2020
Year

### Benin

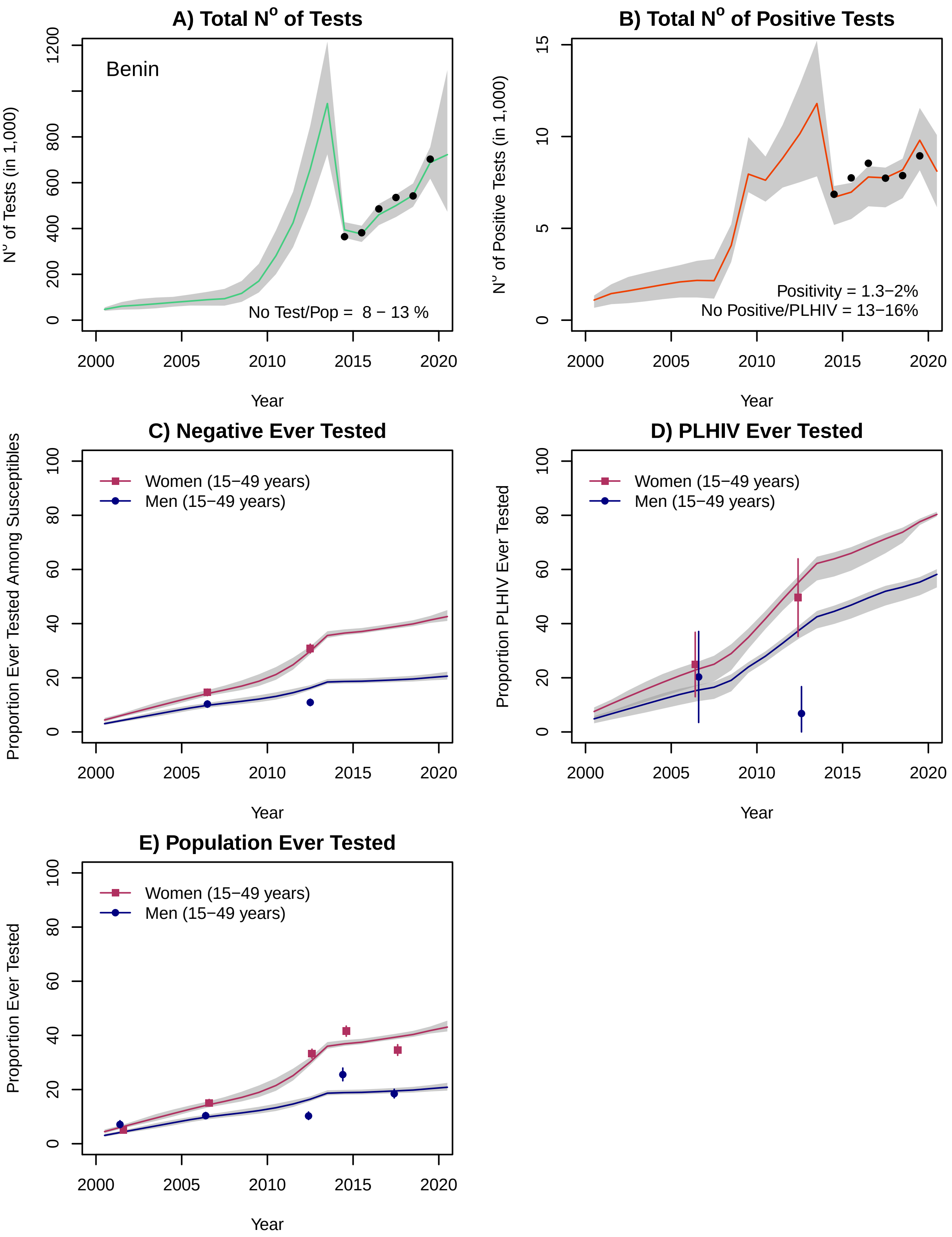
A) Total N° of Tests
Benin
N° of Tests (in 1,000)
No Test/Pop = 8 − 13 %
Year
B) Total N° of Positive Tests
N° of Positive Tests (in 1,000)
Positivity = 1.3−2%
No Positive/PLHIV = 13−16%
Year
C) Negative Ever Tested
Proportion Ever Tested Among Susceptibles
Women (15−49 years)
Men (15−49 years)
Year
D) PLHIV Ever Tested
Proportion PLHIV Ever Tested
Women (15−49 years)
Men (15−49 years)
Year
E) Population Ever Tested
Proportion Ever Tested
Women (15−49 years)
Men (15−49 years)
Year

### Botswana

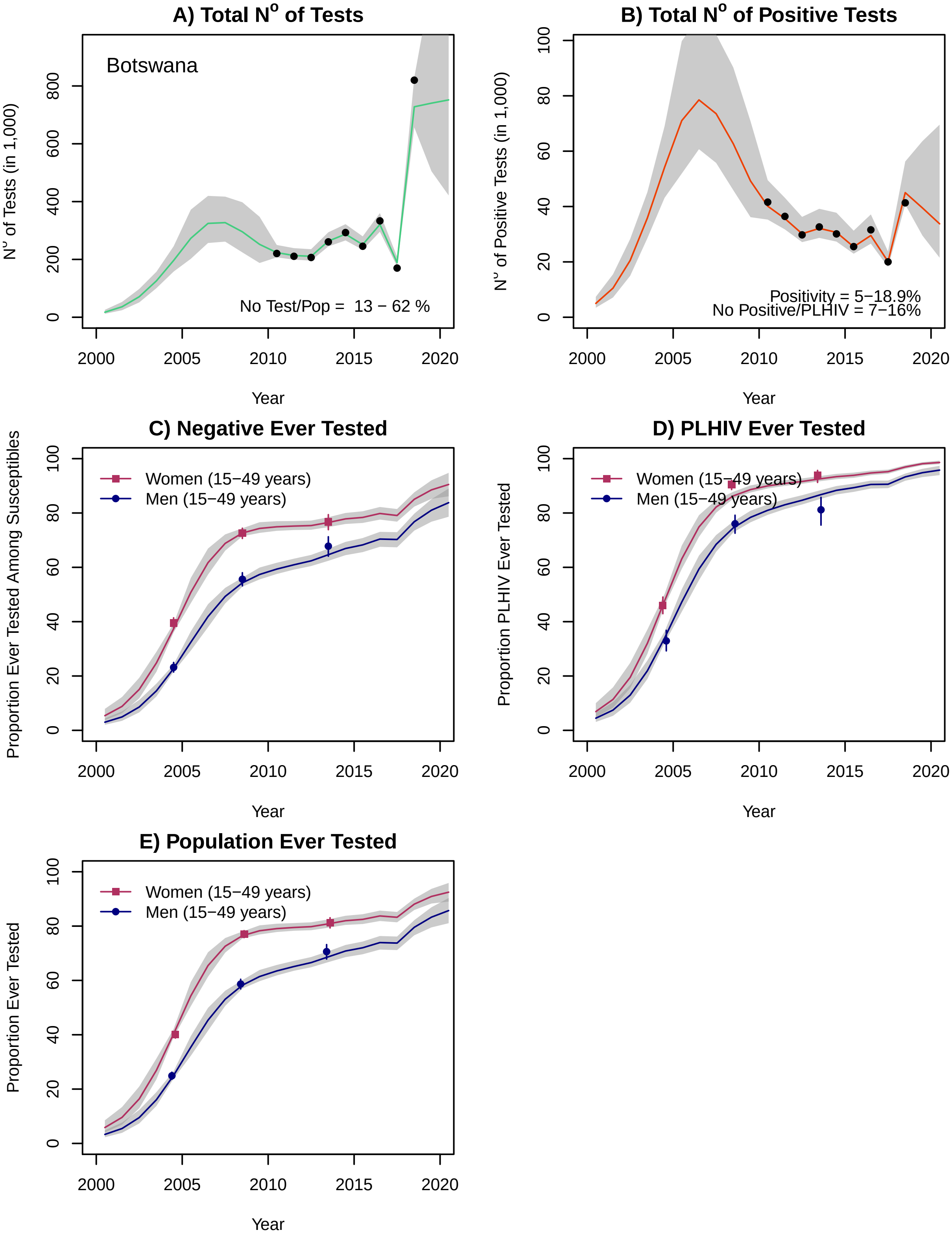
A) Total N° of Tests
Botswana
N° of Tests (in 1,000)
No Test/Pop = 13 − 62 %
Year
B) Total N° of Positive Tests
N° of Positive Tests (in 1,000)
Positivity = 5−18.9%
No Positive/PLHIV = 7−16%
Year
C) Negative Ever Tested
Proportion Ever Tested Among Susceptibles
Women (15−49 years)
Men (15−49 years)
Year
D) PLHIV Ever Tested
Proportion PLHIV Ever Tested
Women (15−49 years)
Men (15−49 years)
Year
E) Population Ever Tested
Proportion Ever Tested
Women (15−49 years)
Men (15−49 years)
Year

#### Burkina Faso

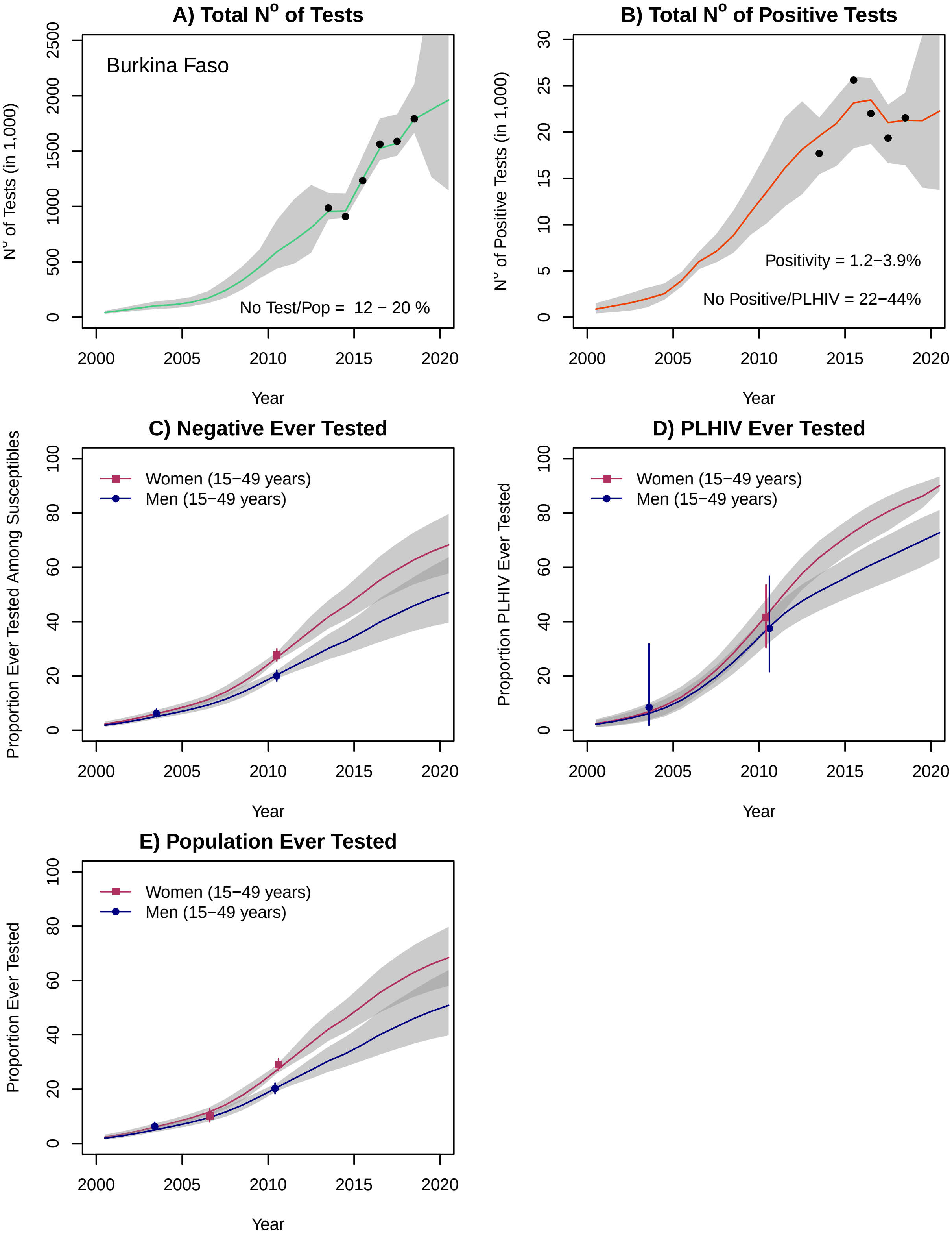

### Burundi

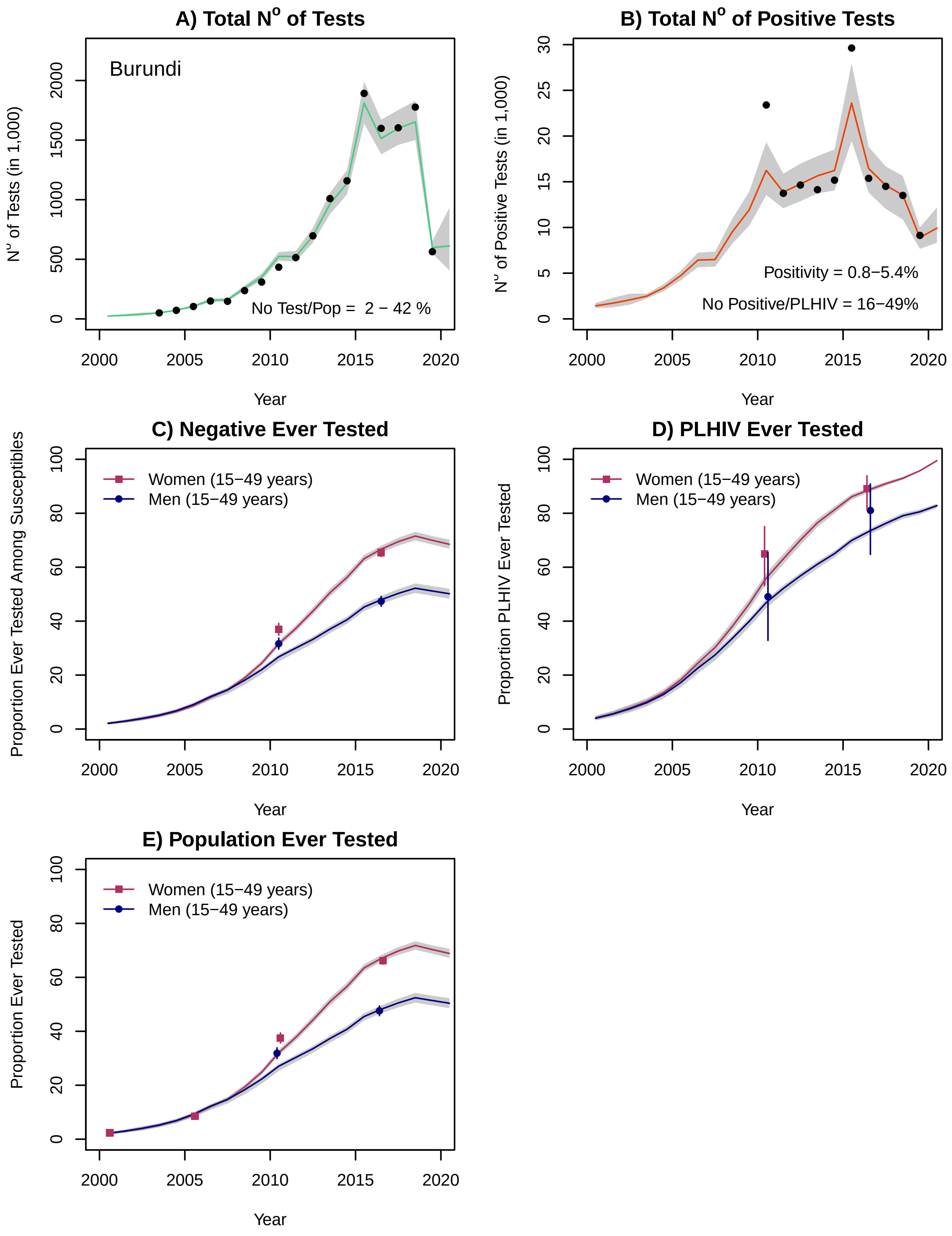

A) Total N° of Tests
Burundi
No Test/Pop = 2 − 42 %
N° of Tests (in 1,000)
Year
B) Total N° of Positive Tests
Positivity = 0.8−5.4%
No Positive/PLHIV = 16−49%
N° of Positive Tests (in 1,000)
Year
C) Negative Ever Tested
Women (15−49 years)
Men (15−49 years)
Proportion Ever Tested Among Susceptibles
Year
D) PLHIV Ever Tested
Women (15−49 years)
Men (15−49 years)
Proportion PLHIV Ever Tested
Year
E) Population Ever Tested
Women (15−49 years)
Men (15−49 years)
Proportion Ever Tested
Year

### Cameroon*

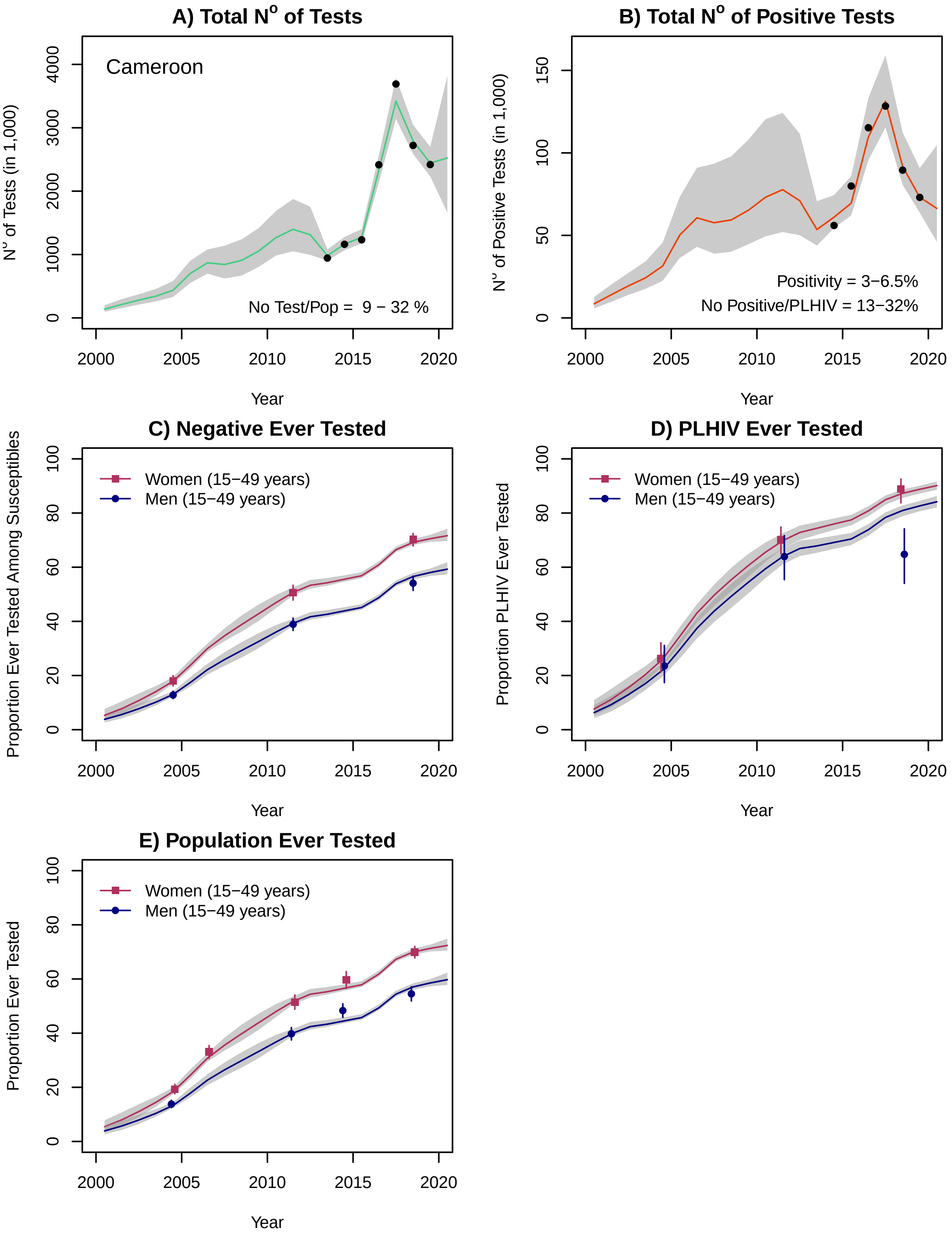
A) Total N° of Tests
Cameroon
No Test/Pop = 9 − 32 %
N° of Tests (in 1,000)
Year
B) Total N° of Positive Tests
Positivity = 3−6.5%
No Positive/PLHIV = 13−32%
N° of Positive Tests (in 1,000)
Year
C) Negative Ever Tested
Women (15−49 years)
Men (15−49 years)
Proportion Ever Tested Among Susceptibles
Year
D) PLHIV Ever Tested
Women (15−49 years)
Men (15−49 years)
Proportion PLHIV Ever Tested
Year
E) Population Ever Tested
Women (15−49 years)
Men (15−49 years)
Proportion Ever Tested
Year

### Chad

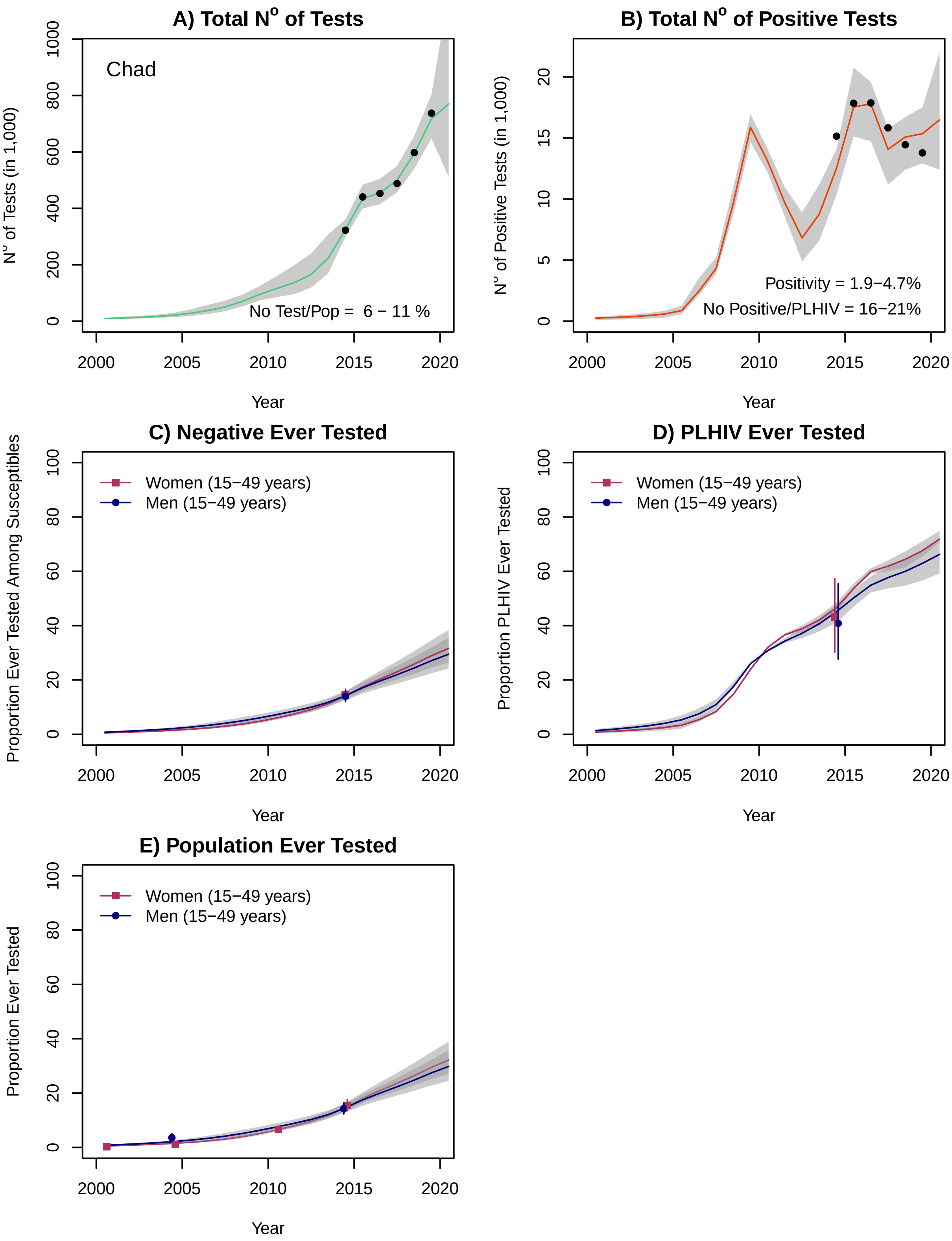
A) Total N° of Tests
Chad
No Test/Pop = 6 − 11 %
N° of Tests (in 1,000)
Year
B) Total N° of Positive Tests
Positivity = 1.9−4.7%
No Positive/PLHIV = 16−21%
N° of Positive Tests (in 1,000)
Year
C) Negative Ever Tested
Women (15−49 years)
Men (15−49 years)
Proportion Ever Tested Among Susceptibles
Year
D) PLHIV Ever Tested
Women (15−49 years)
Men (15−49 years)
Proportion PLHIV Ever Tested
Year
E) Population Ever Tested
Women (15−49 years)
Men (15−49 years)
Proportion Ever Tested
Year

### Comoros

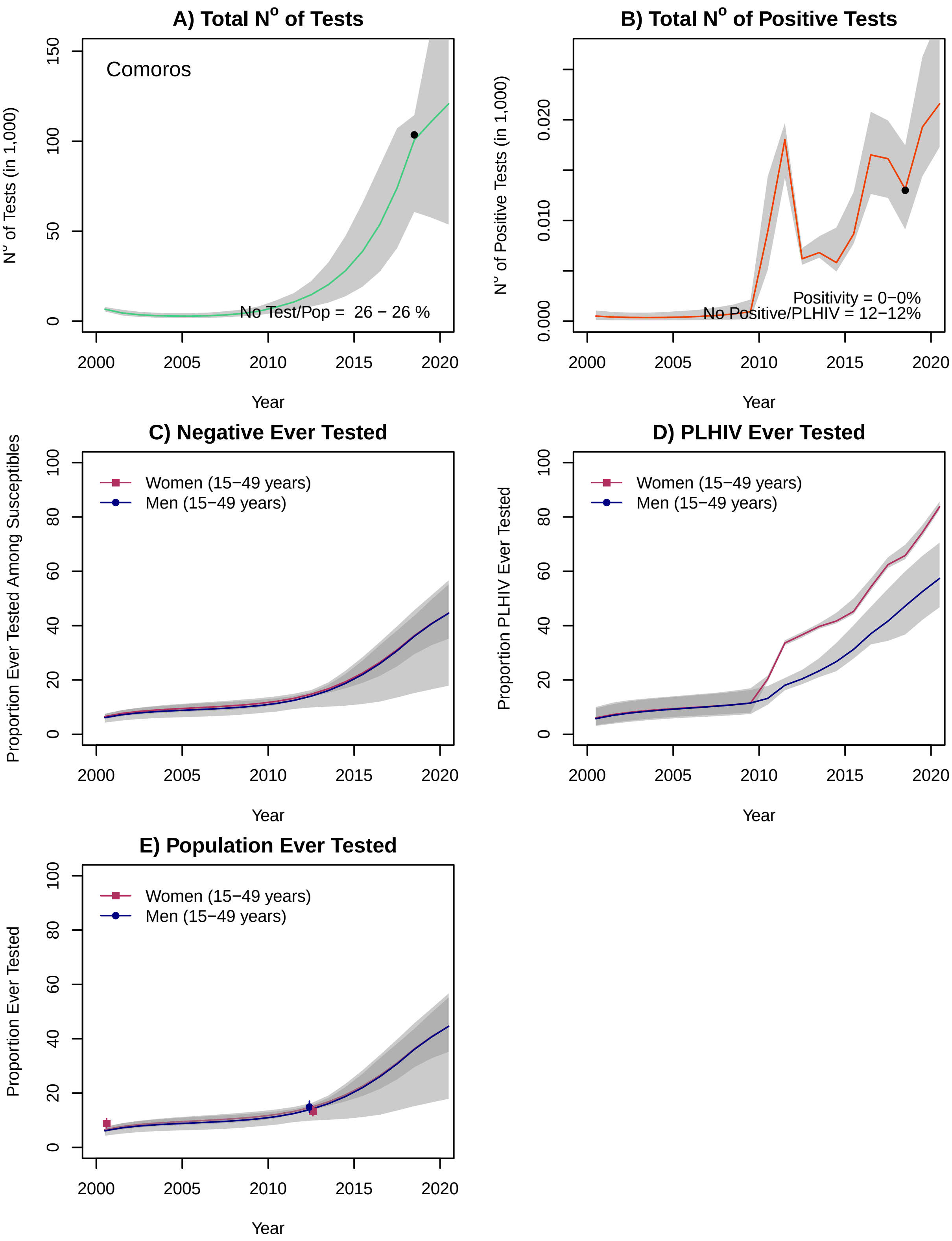

A) Total N° of Tests
Comoros
N° of Tests (in 1,000)
No Test/Pop = 26 − 26 %
B) Total N° of Positive Tests
N° of Positive Tests (in 1,000)
Positivity = 0−0%
No Positive/PLHIV = 12−12%
C) Negative Ever Tested
Proportion Ever Tested Among Susceptibles
D) PLHIV Ever Tested
Proportion PLHIV Ever Tested
E) Population Ever Tested
Proportion Ever Tested
Women (15−49 years)
Men (15−49 years)
Year

### Congo

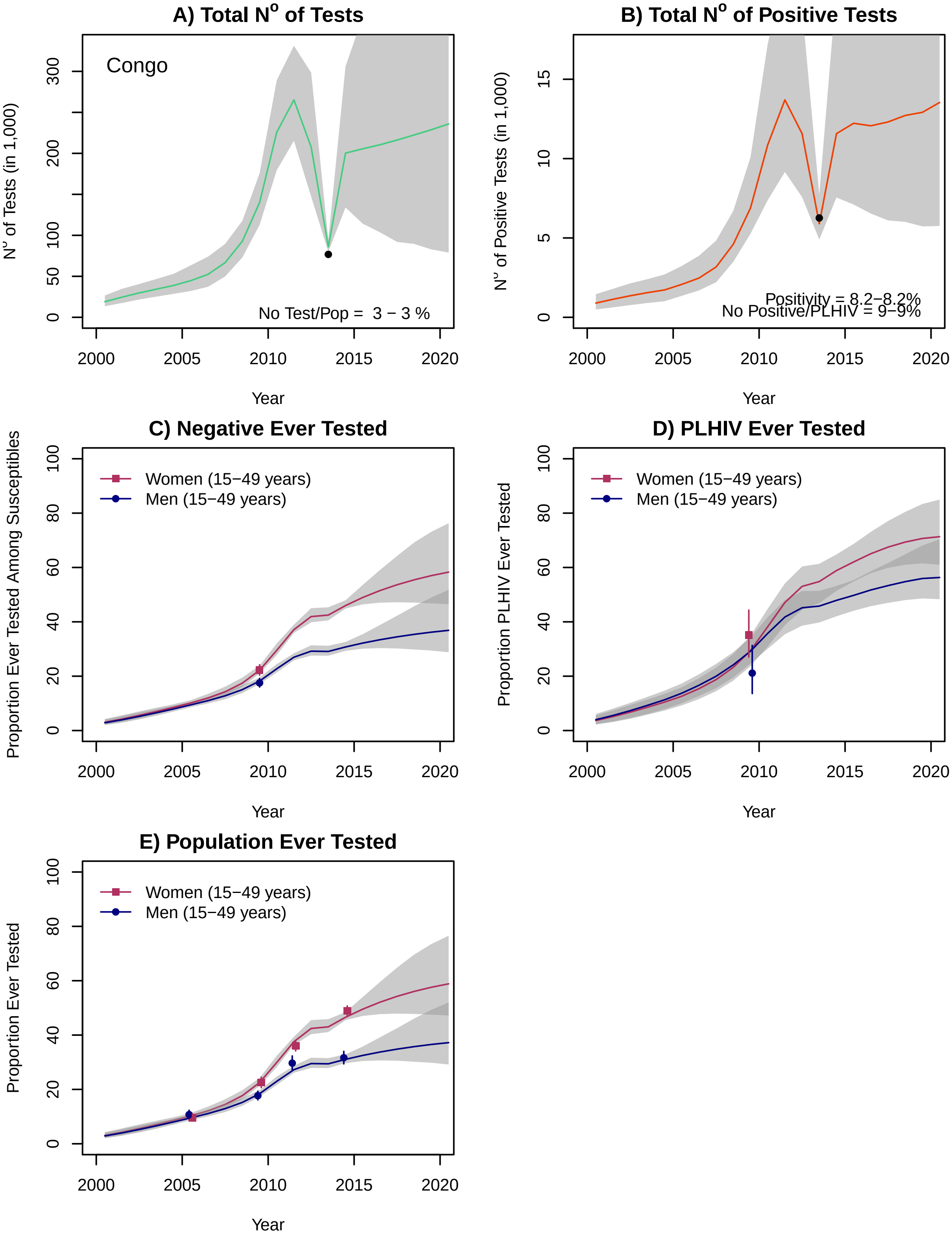
A) Total N$^o$ of Tests
Congo
N$^o$ of Tests (in 1,000)
No Test/Pop = 3 − 3 %
Year
B) Total N$^o$ of Positive Tests
N$^o$ of Positive Tests (in 1,000)
Positivity = 8.2−8.2%
No Positive/PLHIV = 9−9%
Year
C) Negative Ever Tested
Proportion Ever Tested Among Susceptibles
Women (15−49 years)
Men (15−49 years)
Year
D) PLHIV Ever Tested
Proportion PLHIV Ever Tested
Women (15−49 years)
Men (15−49 years)
Year
E) Population Ever Tested
Proportion Ever Tested
Women (15−49 years)
Men (15−49 years)
Year

### Cote d'Ivoire (Ivory Coast)*

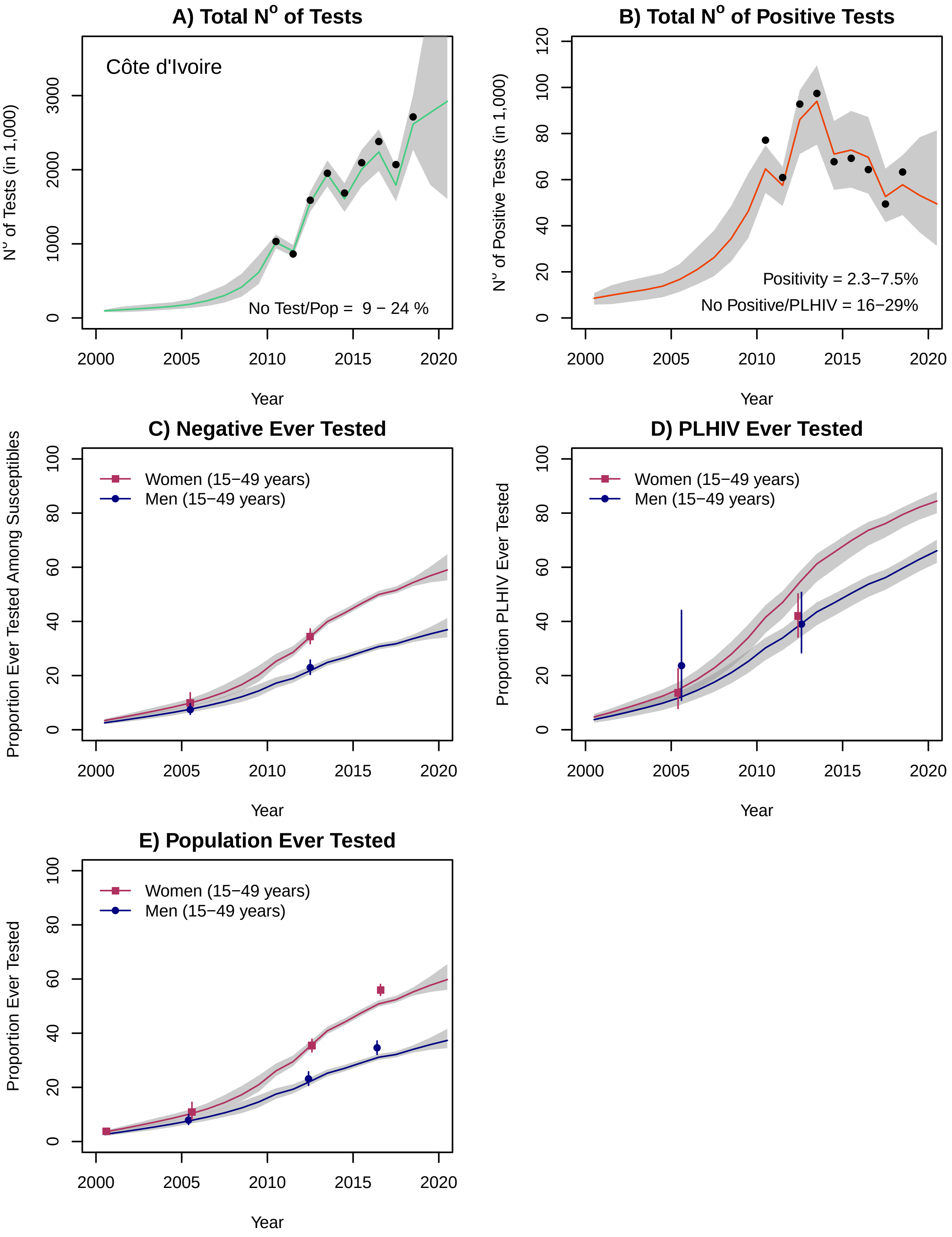

### Democratic Republic of the Congo

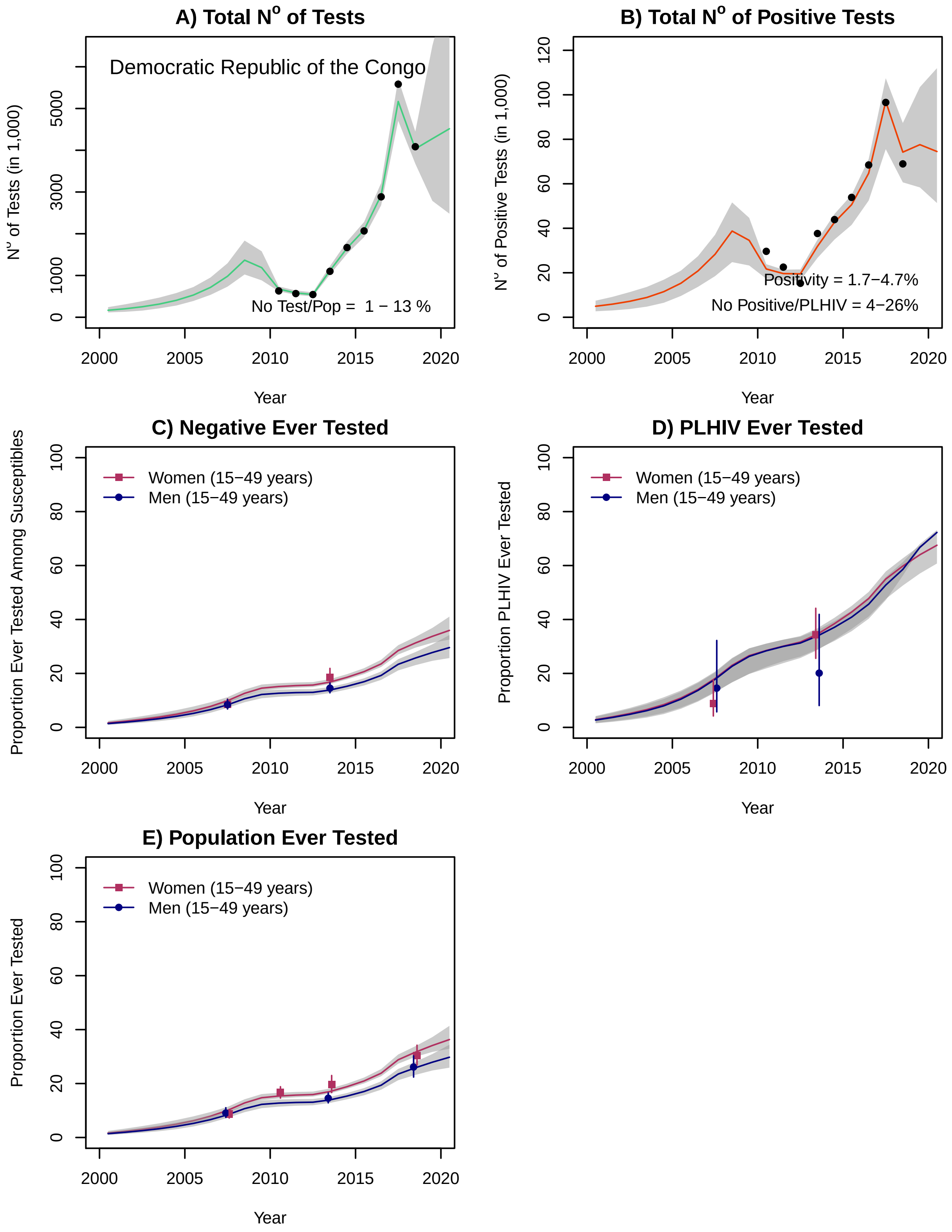

#### Equatorial Guinea

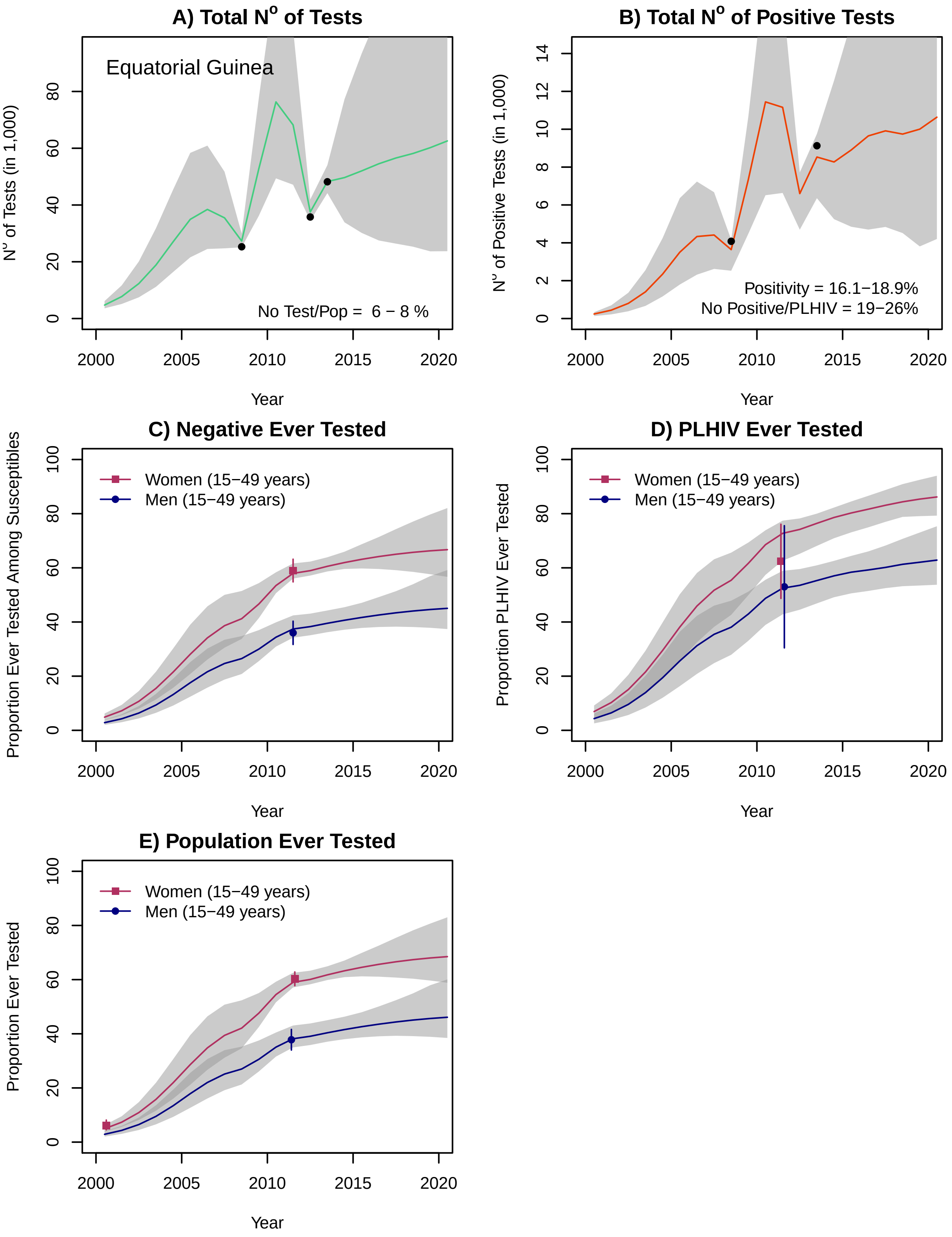

### Eritrea

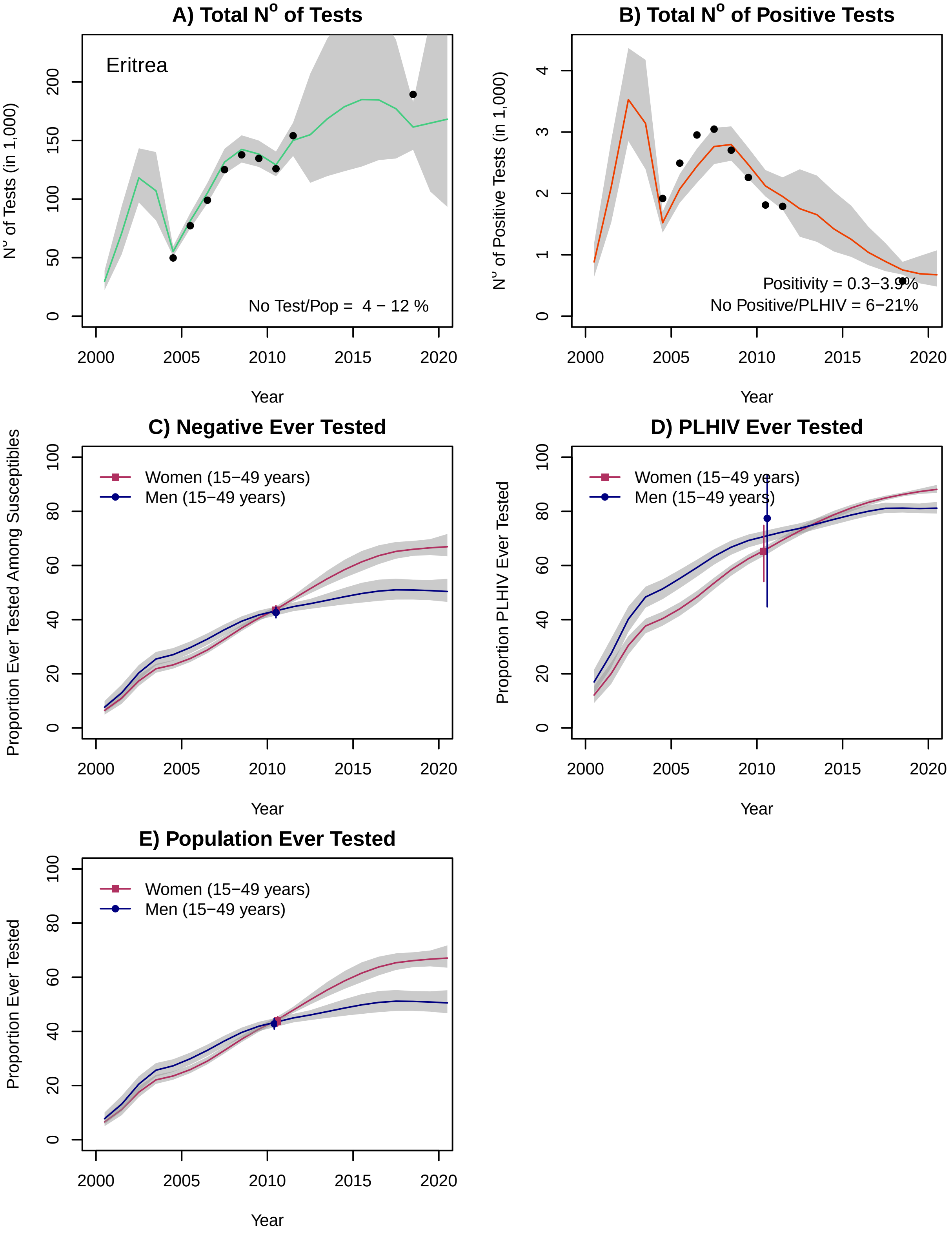
A) Total N° of Tests
Eritrea
No Test/Pop = 4 − 12 %
N° of Tests (in 1,000)
Year
B) Total N° of Positive Tests
Positivity = 0.3−3.9%
No Positive/PLHIV = 6−21%
N° of Positive Tests (in 1,000)
Year
C) Negative Ever Tested
Women (15−49 years)
Men (15−49 years)
Proportion Ever Tested Among Susceptibles
Year
D) PLHIV Ever Tested
Women (15−49 years)
Men (15−49 years)
Proportion PLHIV Ever Tested
Year
E) Population Ever Tested
Women (15−49 years)
Men (15−49 years)
Proportion Ever Tested
Year

#### eSwatini

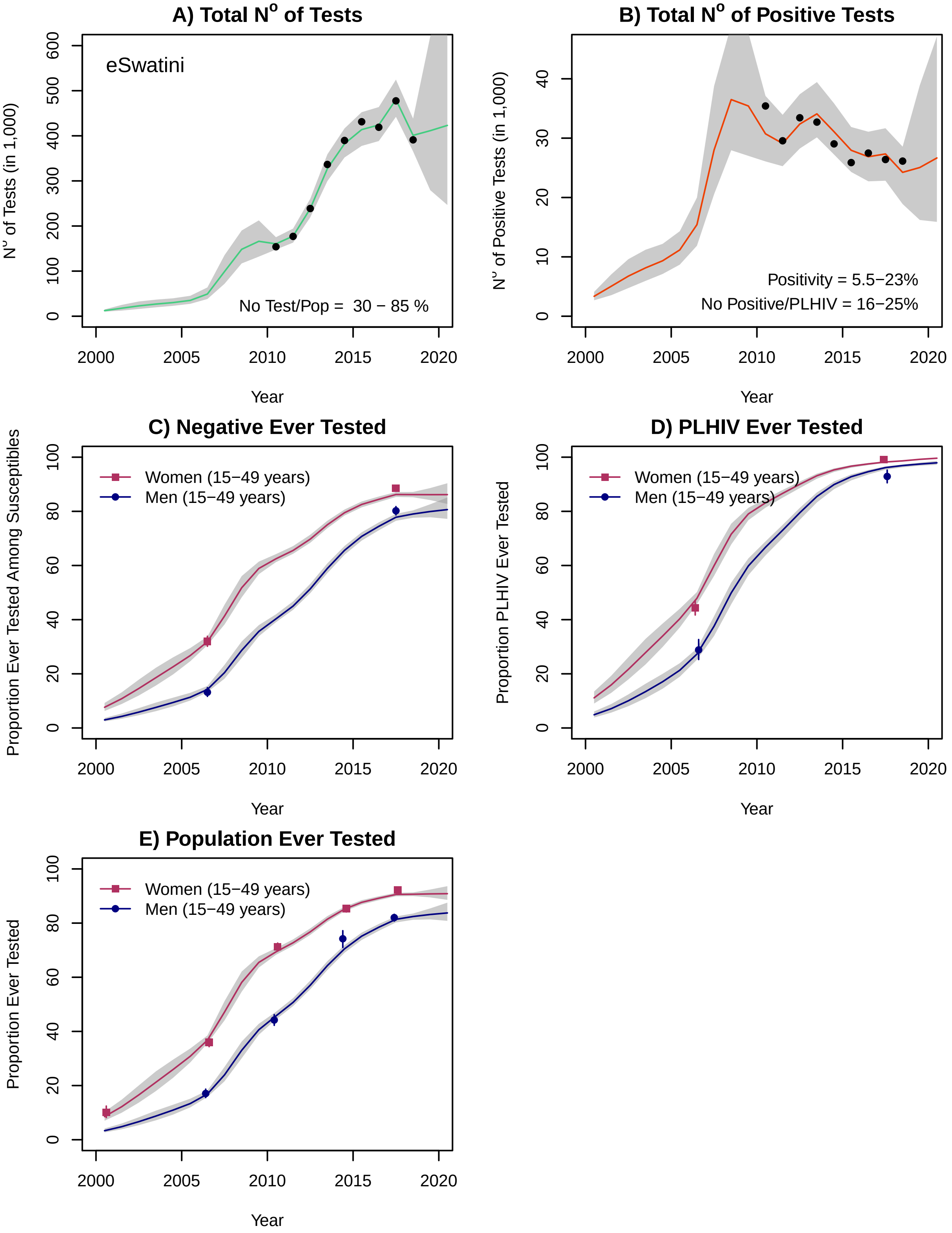

A) Total N° of Tests
eSwatini
No Test/Pop = 30 − 85 %
N° of Tests (in 1,000)
Year
B) Total N° of Positive Tests
Positivity = 5.5−23%
No Positive/PLHIV = 16−25%
N° of Positive Tests (in 1,000)
Year
C) Negative Ever Tested
Women (15−49 years)
Men (15−49 years)
Proportion Ever Tested Among Susceptibles
Year
D) PLHIV Ever Tested
Women (15−49 years)
Men (15−49 years)
Proportion PLHIV Ever Tested
Year
E) Population Ever Tested
Women (15−49 years)
Men (15−49 years)
Proportion Ever Tested
Year

### Ethiopia

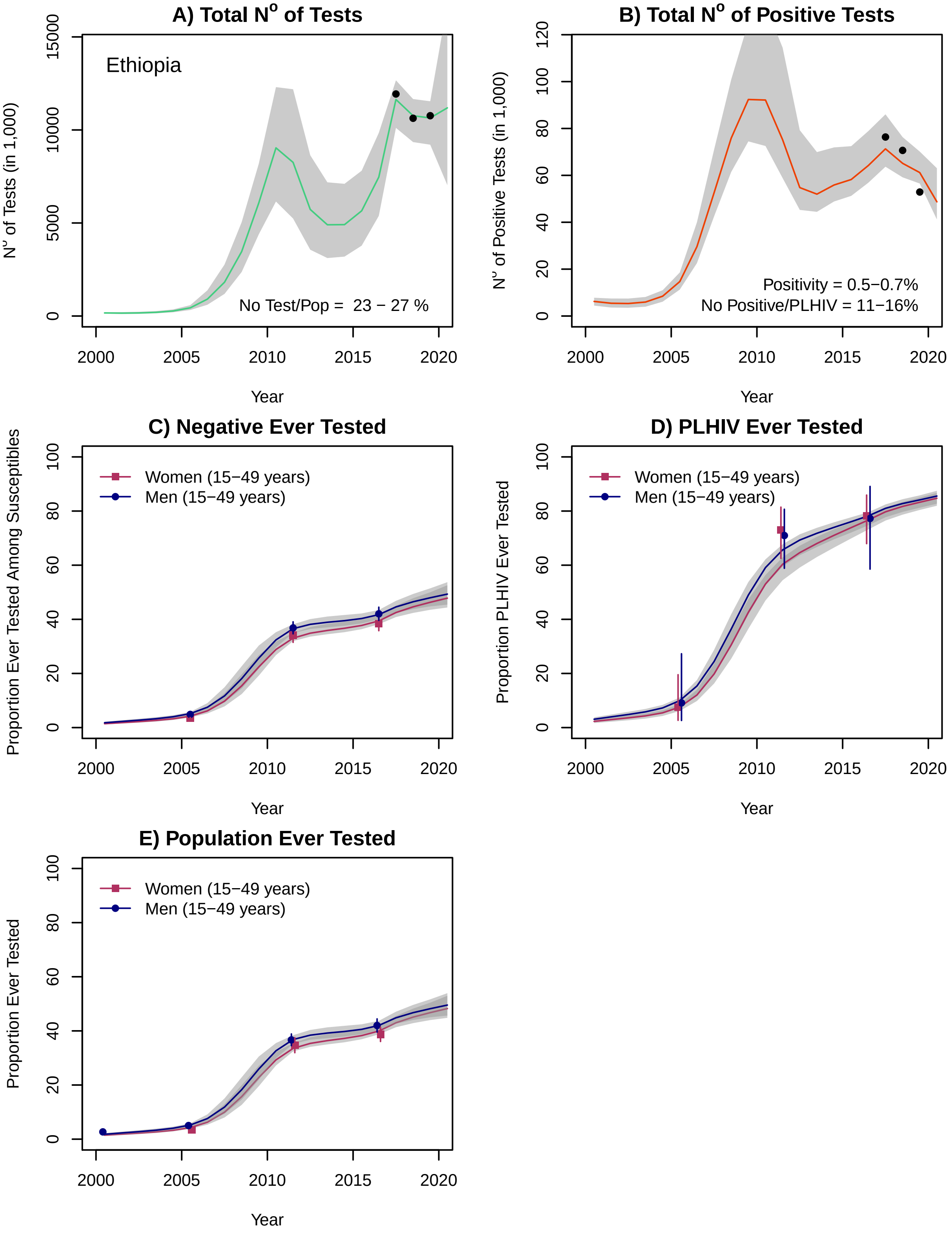
A) Total N$^o$ of Tests
Ethiopia
N$^o$ of Tests (in 1,000)
No Test/Pop = 23 − 27 %
Year
B) Total N$^o$ of Positive Tests
N$^o$ of Positive Tests (in 1,000)
Positivity = 0.5−0.7%
No Positive/PLHIV = 11−16%
Year
C) Negative Ever Tested
Proportion Ever Tested Among Susceptibles
Women (15−49 years)
Men (15−49 years)
Year
D) PLHIV Ever Tested
Proportion PLHIV Ever Tested
Women (15−49 years)
Men (15−49 years)
Year
E) Population Ever Tested
Proportion Ever Tested
Women (15−49 years)
Men (15−49 years)
Year

### Gabon

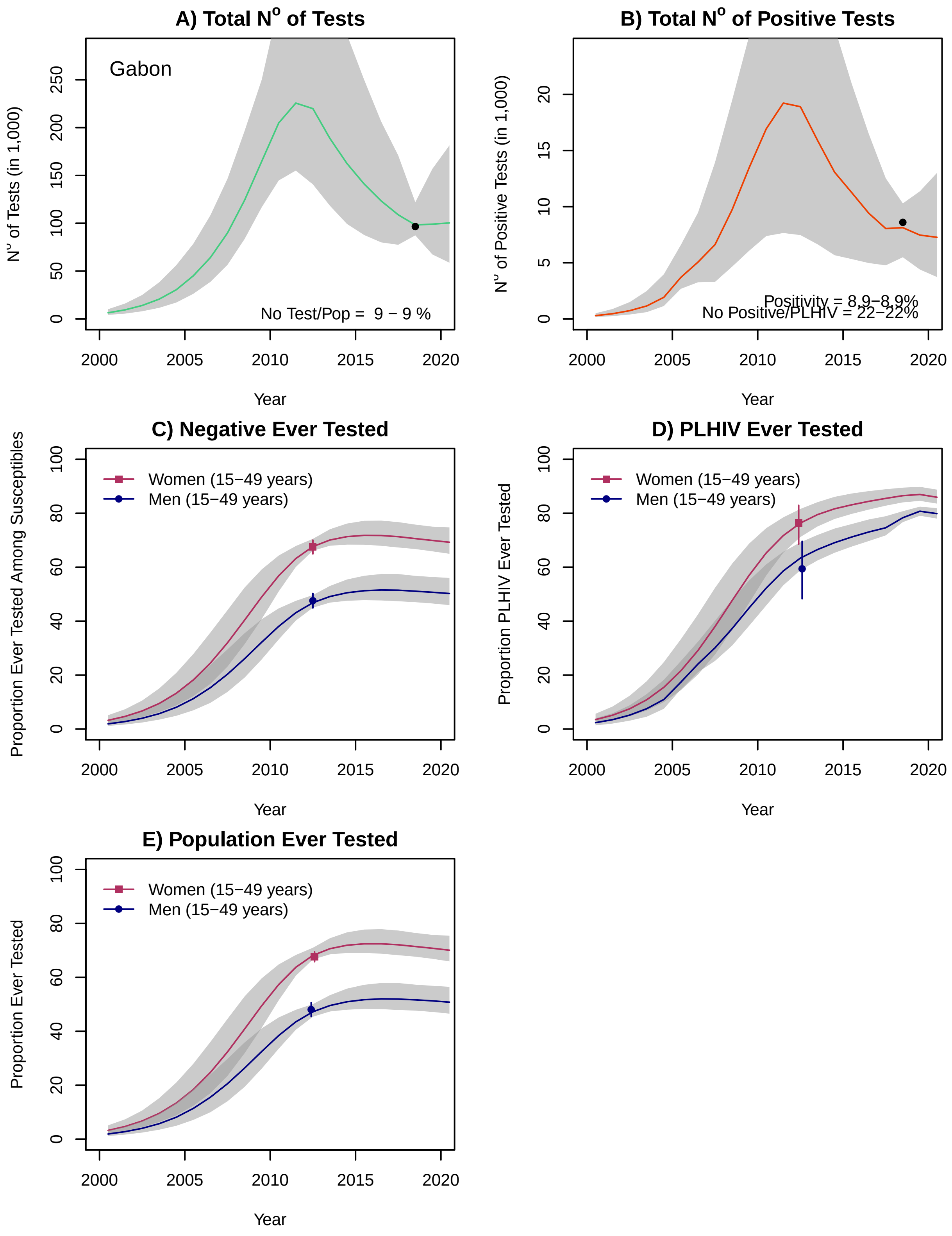

A) Total N° of Tests
Gabon
N° of Tests (in 1,000)
No Test/Pop = 9 − 9 %
B) Total N° of Positive Tests
N° of Positive Tests (in 1,000)
Positivity = 8.9−8.9%
No Positive/PLHIV = 22−22%
C) Negative Ever Tested
Proportion Ever Tested Among Susceptibles
D) PLHIV Ever Tested
Proportion PLHIV Ever Tested
E) Population Ever Tested
Proportion Ever Tested
Women (15−49 years)
Men (15−49 years)
Year

### Gambia

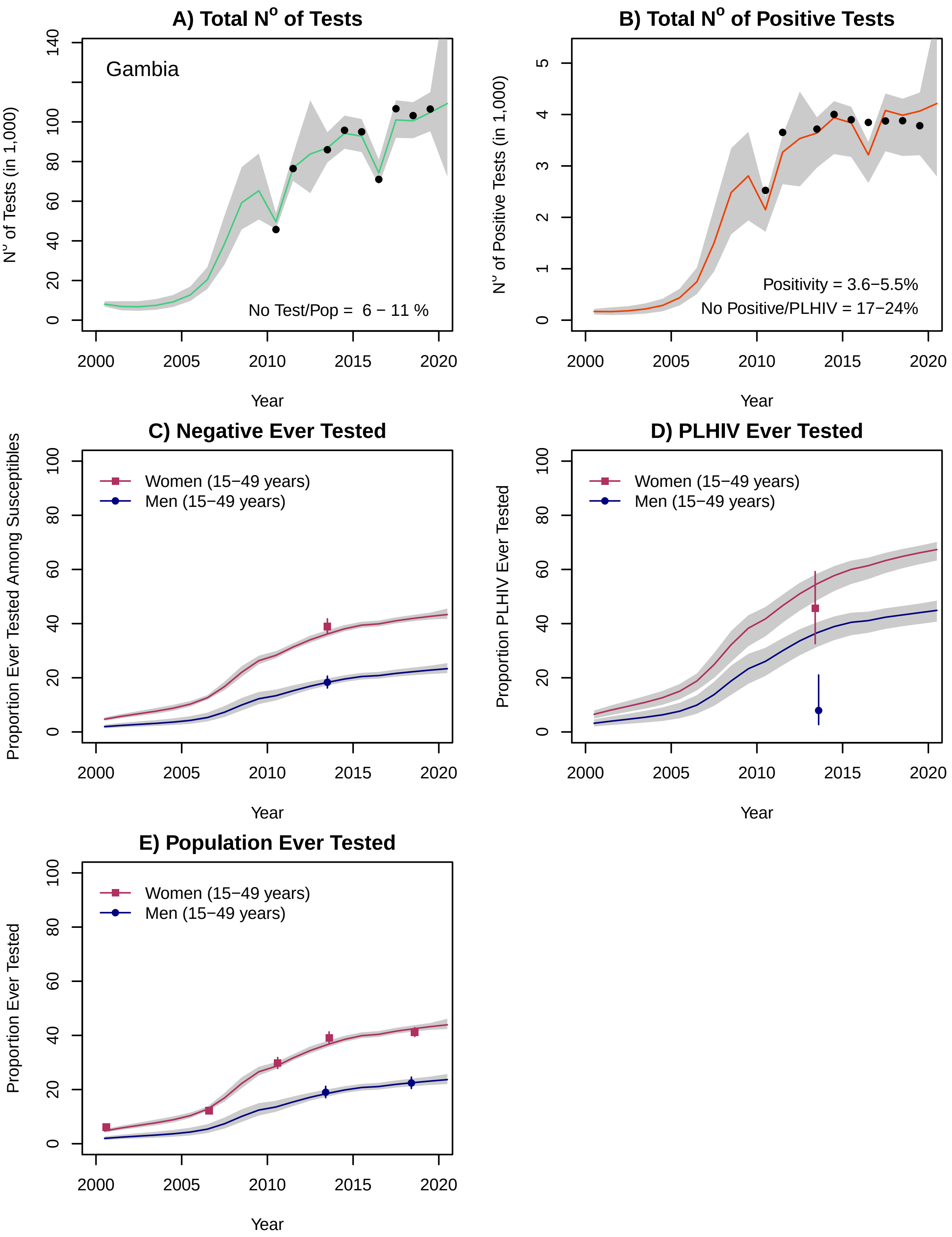
A) Total N° of Tests
Gambia
No Test/Pop = 6 − 11 %
N° of Tests (in 1,000)
Year
B) Total N° of Positive Tests
Positivity = 3.6−5.5%
No Positive/PLHIV = 17−24%
N° of Positive Tests (in 1,000)
Year
C) Negative Ever Tested
Women (15−49 years)
Men (15−49 years)
Proportion Ever Tested Among Susceptibles
Year
D) PLHIV Ever Tested
Women (15−49 years)
Men (15−49 years)
Proportion PLHIV Ever Tested
Year
E) Population Ever Tested
Women (15−49 years)
Men (15−49 years)
Proportion Ever Tested
Year

### Ghana

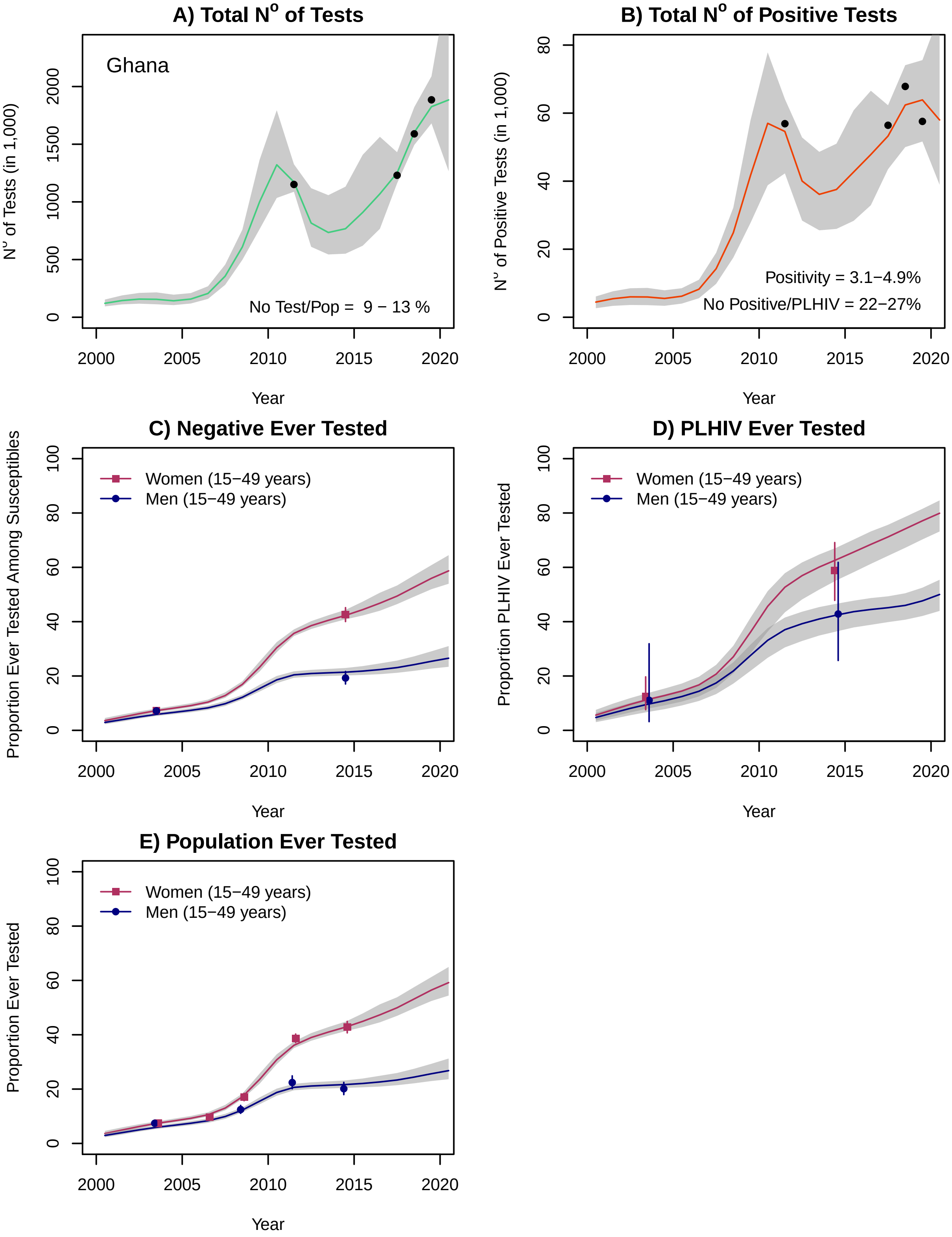
A) Total N° of Tests
Ghana
N° of Tests (in 1,000)
No Test/Pop = 9 − 13 %
B) Total N° of Positive Tests
N° of Positive Tests (in 1,000)
Positivity = 3.1−4.9%
No Positive/PLHIV = 22−27%
C) Negative Ever Tested
Proportion Ever Tested Among Susceptibles
D) PLHIV Ever Tested
Proportion PLHIV Ever Tested
E) Population Ever Tested
Proportion Ever Tested
Women (15−49 years)
Men (15−49 years)
Year

### Guinea

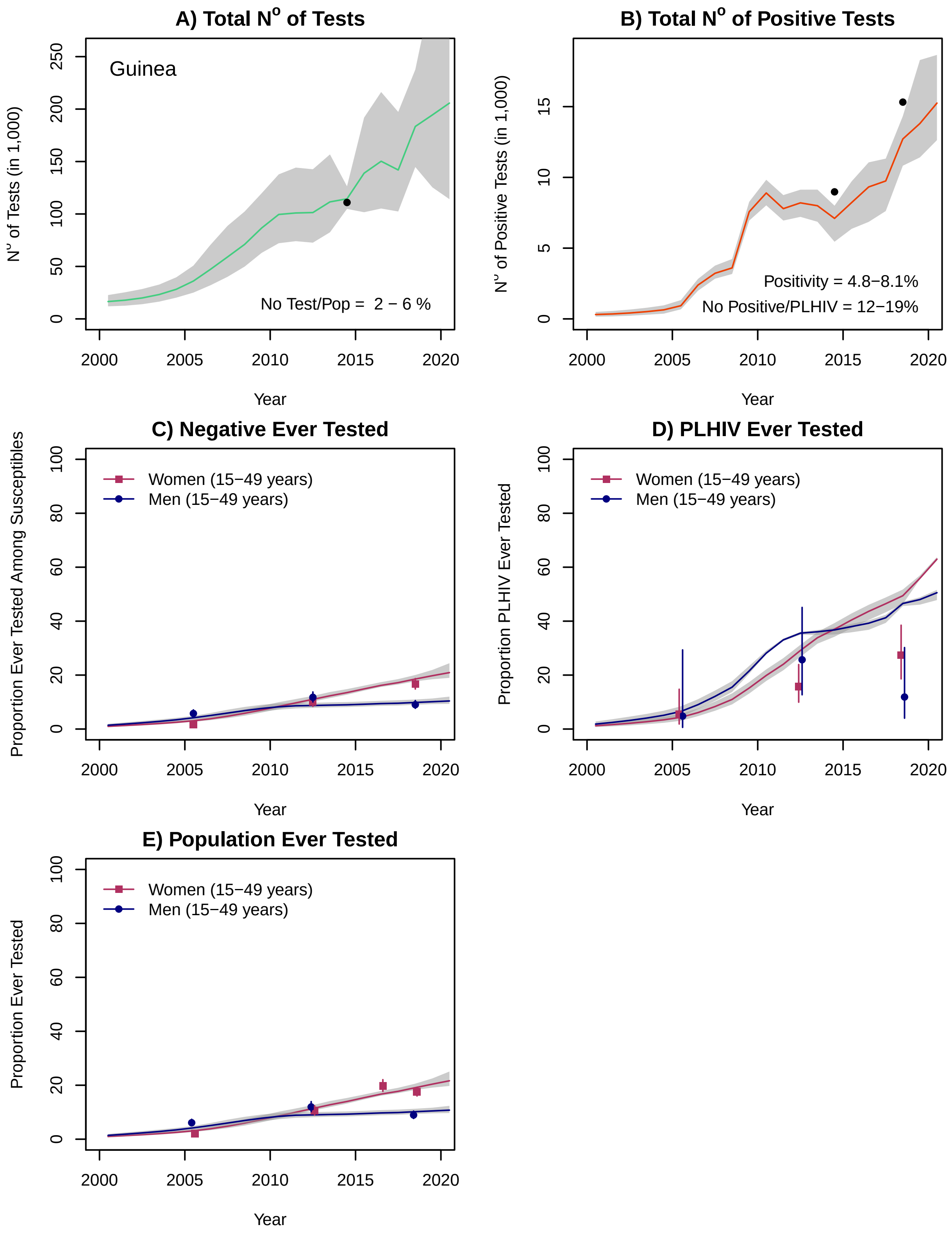

A) Total N° of Tests
Guinea
No Test/Pop = 2 − 6 %
N° of Tests (in 1,000)
Year
B) Total N° of Positive Tests
Positivity = 4.8−8.1%
No Positive/PLHIV = 12−19%
N° of Positive Tests (in 1,000)
Year
C) Negative Ever Tested
Women (15−49 years)
Men (15−49 years)
Proportion Ever Tested Among Susceptibles
Year
D) PLHIV Ever Tested
Women (15−49 years)
Men (15−49 years)
Proportion PLHIV Ever Tested
Year
E) Population Ever Tested
Women (15−49 years)
Men (15−49 years)
Proportion Ever Tested
Year

### Kenya*

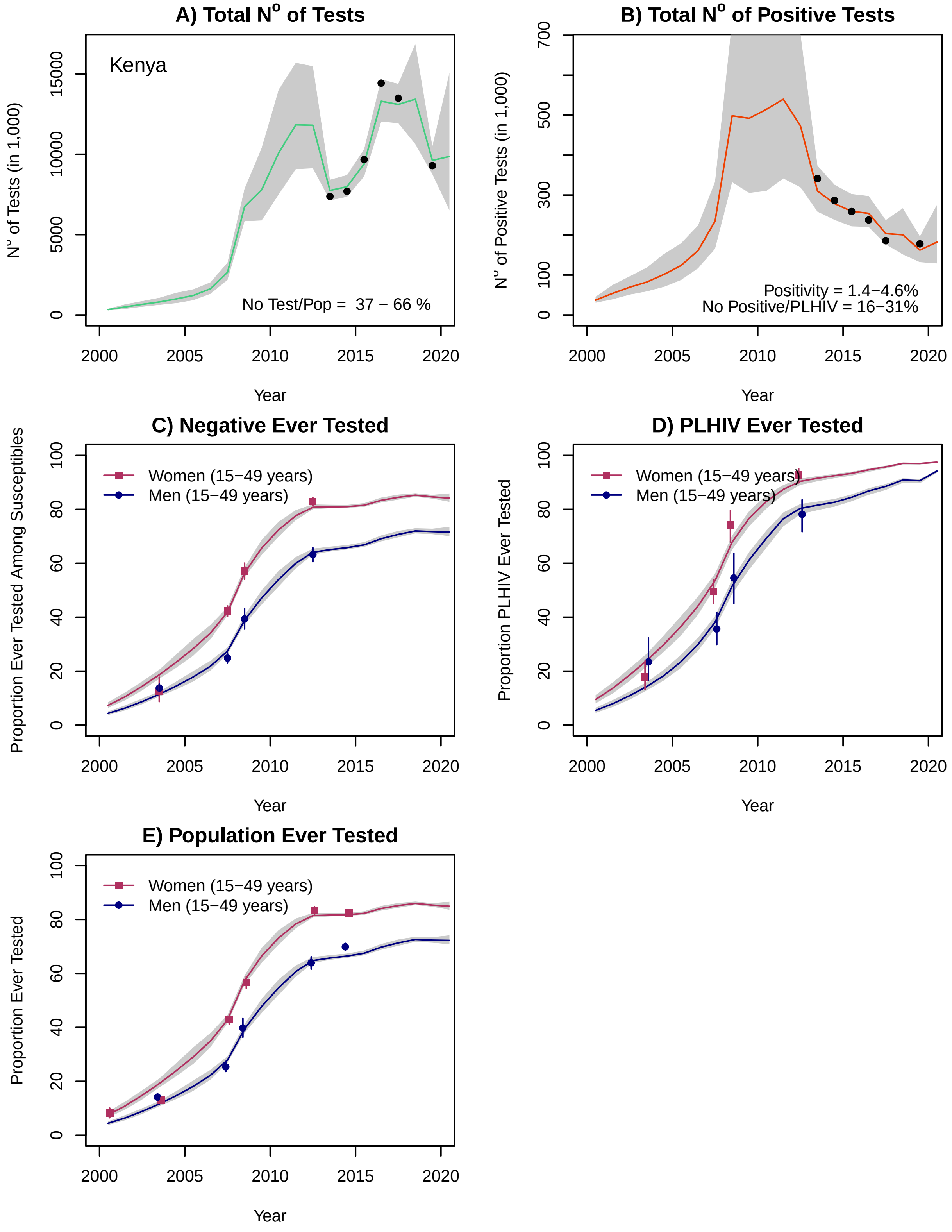
A) Total N$^o$ of Tests
Kenya
No Test/Pop = 37 − 66 %
N$^o$ of Tests (in 1,000)
Year
B) Total N$^o$ of Positive Tests
Positivity = 1.4−4.6%
No Positive/PLHIV = 16−31%
N$^o$ of Positive Tests (in 1,000)
Year
C) Negative Ever Tested
Women (15−49 years)
Men (15−49 years)
Proportion Ever Tested Among Susceptibles
Year
D) PLHIV Ever Tested
Women (15−49 years)
Men (15−49 years)
Proportion PLHIV Ever Tested
Year
E) Population Ever Tested
Women (15−49 years)
Men (15−49 years)
Proportion Ever Tested
Year

### Lesotho*

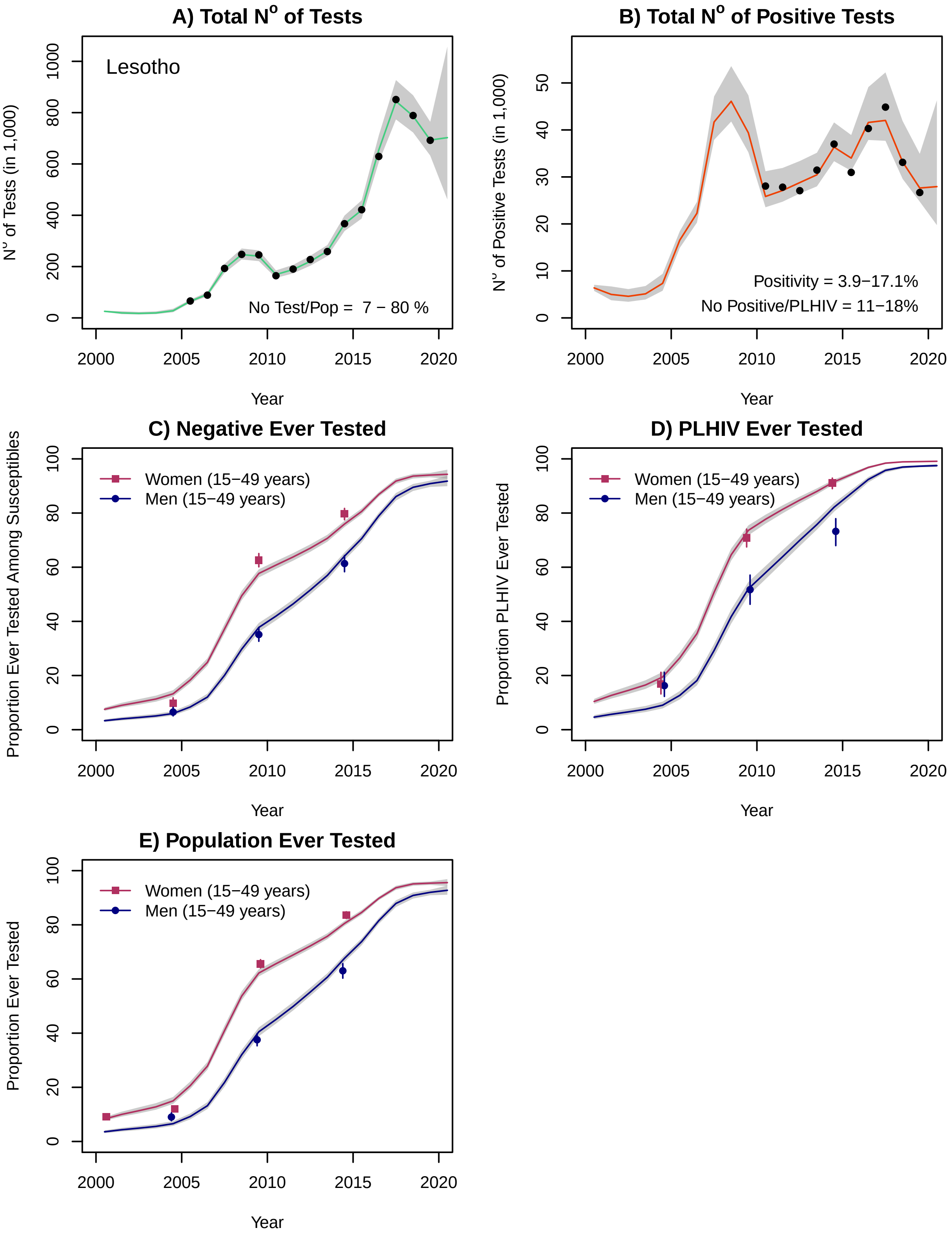
A) Total N° of Tests
Lesotho
No Test/Pop = 7 − 80 %
N° of Tests (in 1,000)
0
200
400
600
800
1000
2000
2005
2010
2015
2020
Year
B) Total N° of Positive Tests
Positivity = 3.9−17.1%
No Positive/PLHIV = 11−18%
N° of Positive Tests (in 1,000)
0
10
20
30
40
50
2000
2005
2010
2015
2020
Year
C) Negative Ever Tested
Women (15−49 years)
Men (15−49 years)
Proportion Ever Tested Among Susceptibles
0
20
40
60
80
100
2000
2005
2010
2015
2020
Year
D) PLHIV Ever Tested
Women (15−49 years)
Men (15−49 years)
Proportion PLHIV Ever Tested
0
20
40
60
80
100
2000
2005
2010
2015
2020
Year
E) Population Ever Tested
Women (15−49 years)
Men (15−49 years)
Proportion Ever Tested
0
20
40
60
80
100
2000
2005
2010
2015
2020
Year

#### Liberia

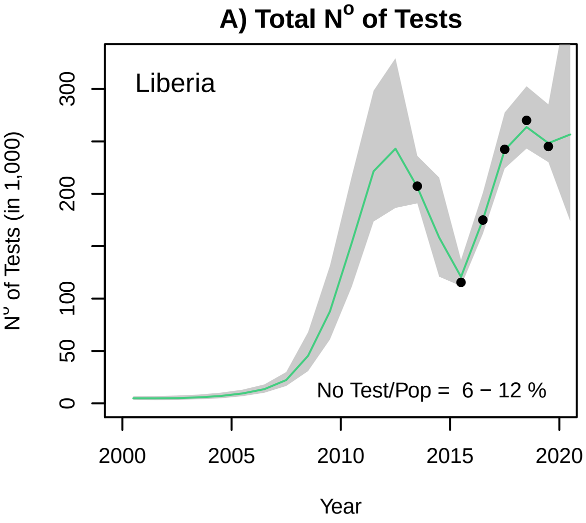
A) Total N° of Tests
Liberia
No Test/Pop = 6 − 12 %
N° of Tests (in 1,000)
Year

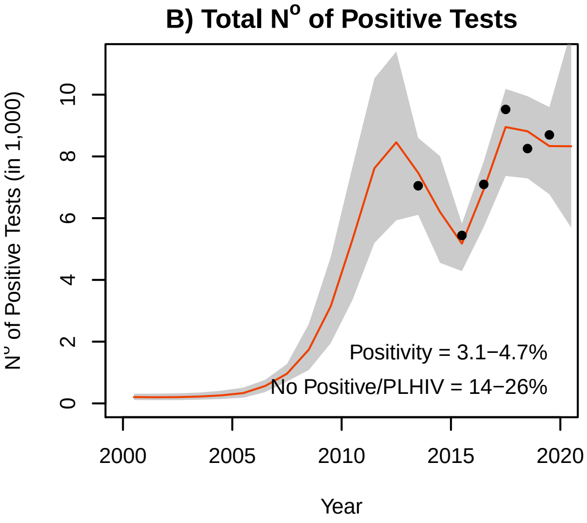
B) Total N° of Positive Tests
Positivity = 3.1−4.7%
No Positive/PLHIV = 14−26%
N° of Positive Tests (in 1,000)
Year

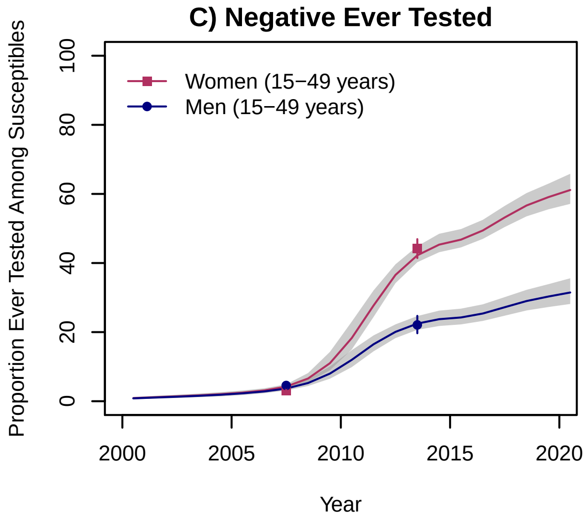
C) Negative Ever Tested
Women (15−49 years)
Men (15−49 years)
Proportion Ever Tested Among Susceptibles
Year

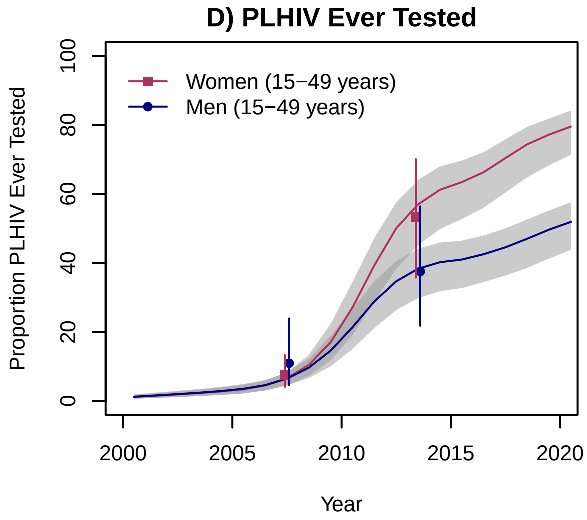
D) PLHIV Ever Tested
Women (15−49 years)
Men (15−49 years)
Proportion PLHIV Ever Tested
Year

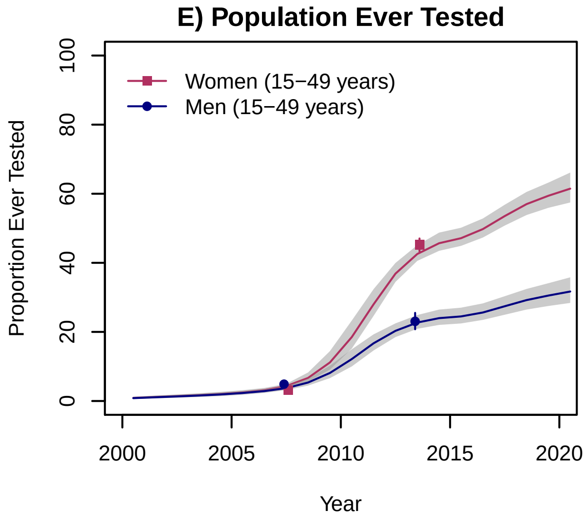
E) Population Ever Tested
Women (15−49 years)
Men (15−49 years)
Proportion Ever Tested
Year

#### Madagascar

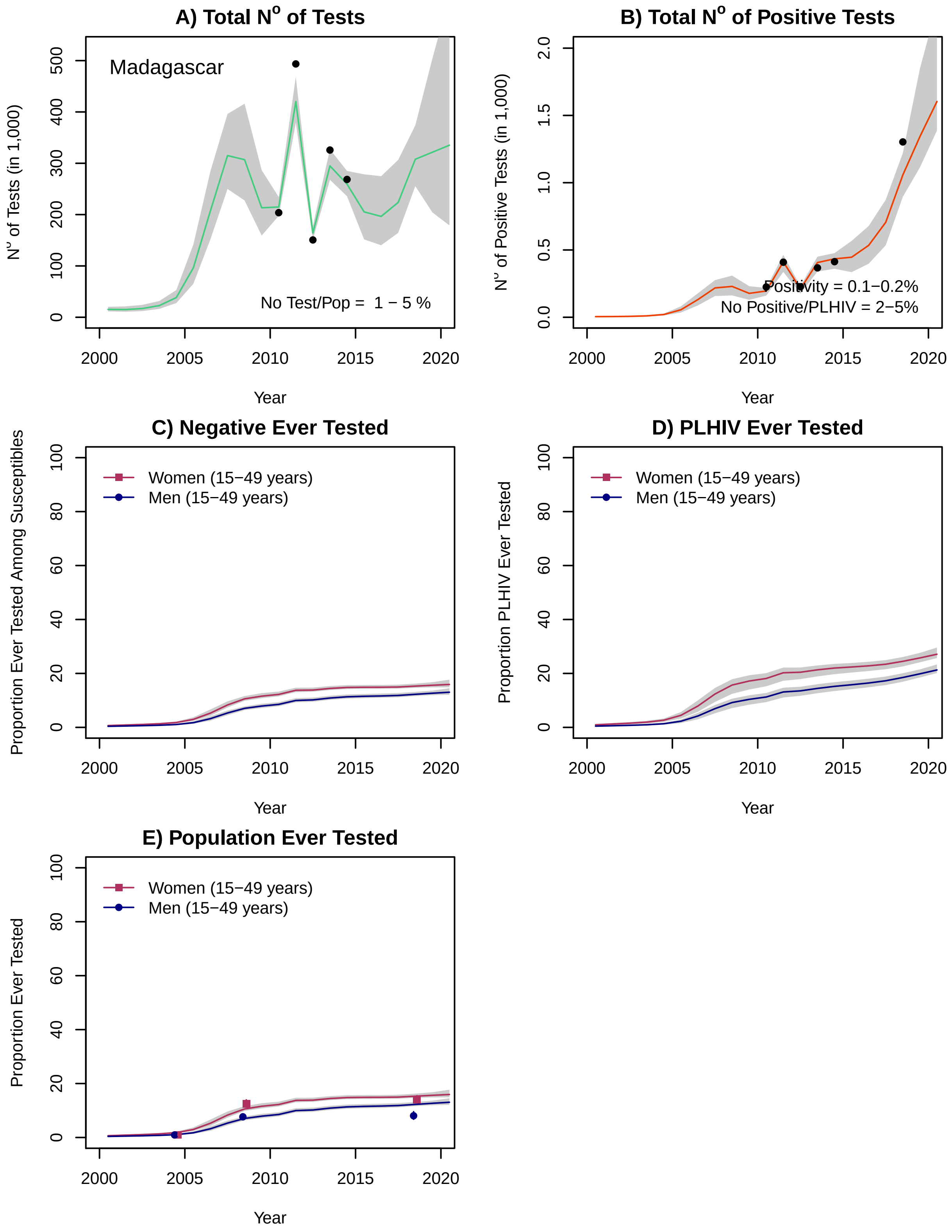

A) Total N° of Tests
N° of Tests (in 1,000)
Madagascar
No Test/Pop = 1 − 5 %
Year
B) Total N° of Positive Tests
N° of Positive Tests (in 1,000)
Positivity = 0.1−0.2%
No Positive/PLHIV = 2−5%
Year
C) Negative Ever Tested
Proportion Ever Tested Among Susceptibles
Women (15−49 years)
Men (15−49 years)
Year
D) PLHIV Ever Tested
Proportion PLHIV Ever Tested
Women (15−49 years)
Men (15−49 years)
Year
E) Population Ever Tested
Proportion Ever Tested
Women (15−49 years)
Men (15−49 years)
Year

### Malawi

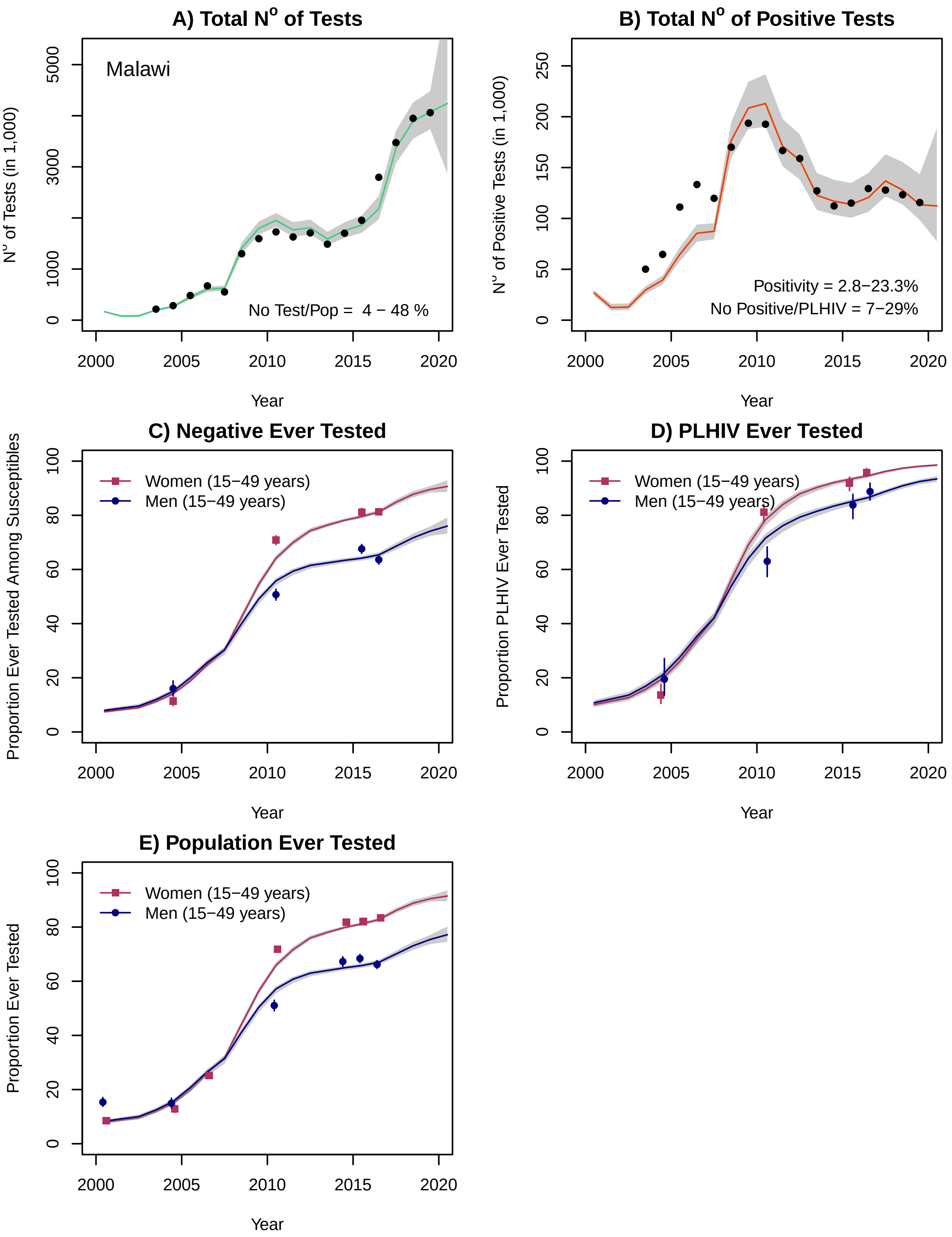
A) Total N° of Tests
Malawi
No Test/Pop = 4 − 48 %
N° of Tests (in 1,000)
Year
B) Total N° of Positive Tests
Positivity = 2.8−23.3%
No Positive/PLHIV = 7−29%
N° of Positive Tests (in 1,000)
Year
C) Negative Ever Tested
Women (15−49 years)
Men (15−49 years)
Proportion Ever Tested Among Susceptibles
Year
D) PLHIV Ever Tested
Women (15−49 years)
Men (15−49 years)
Proportion PLHIV Ever Tested
Year
E) Population Ever Tested
Women (15−49 years)
Men (15−49 years)
Proportion Ever Tested
Year

### Mali

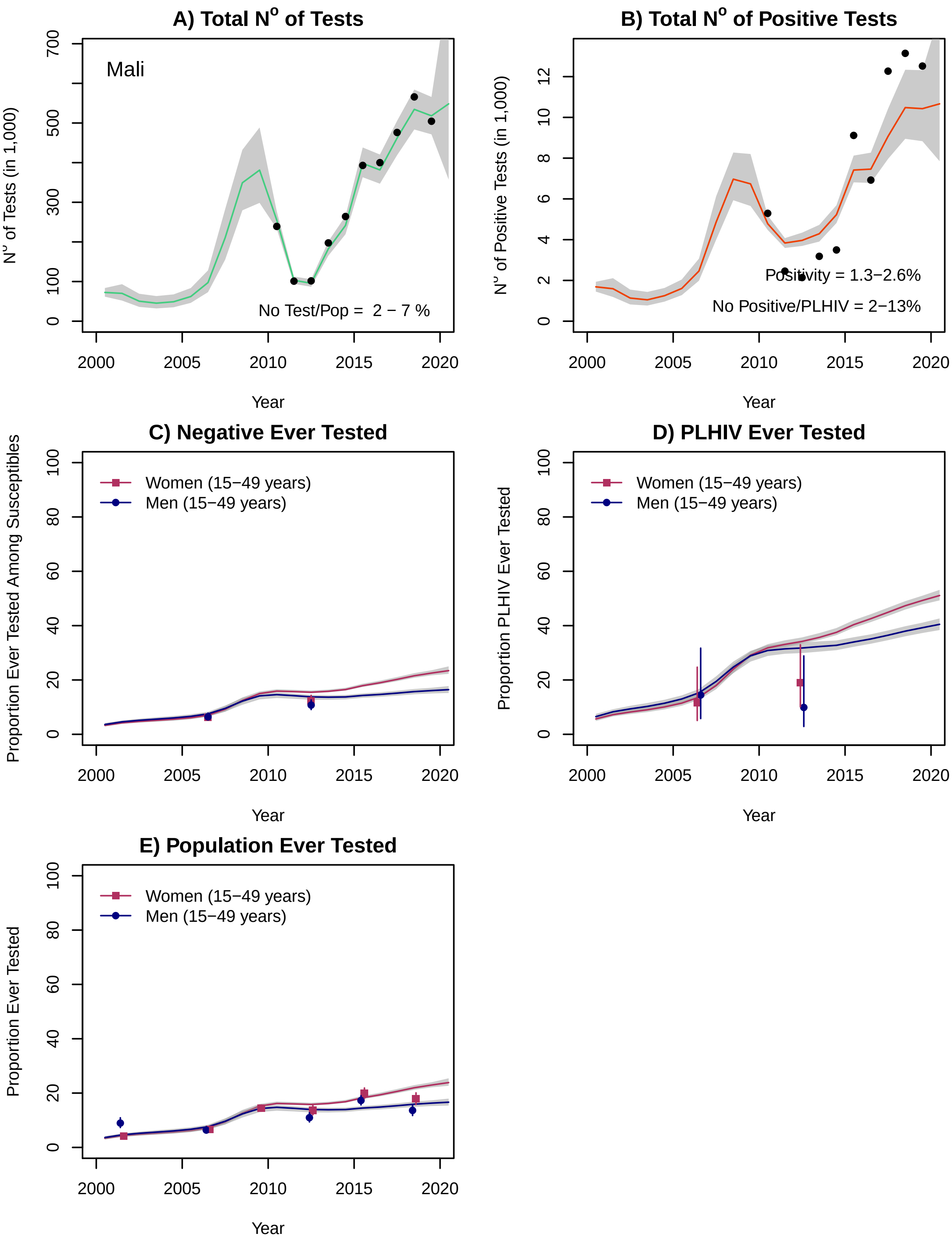

A) Total N° of Tests
Mali
N° of Tests (in 1,000)
No Test/Pop = 2 − 7 %
B) Total N° of Positive Tests
N° of Positive Tests (in 1,000)
Positivity = 1.3−2.6%
No Positive/PLHIV = 2−13%
C) Negative Ever Tested
Proportion Ever Tested Among Susceptibles
Women (15−49 years)
Men (15−49 years)
D) PLHIV Ever Tested
Proportion PLHIV Ever Tested
Women (15−49 years)
Men (15−49 years)
E) Population Ever Tested
Proportion Ever Tested
Women (15−49 years)
Men (15−49 years)
Year

### Mauritania

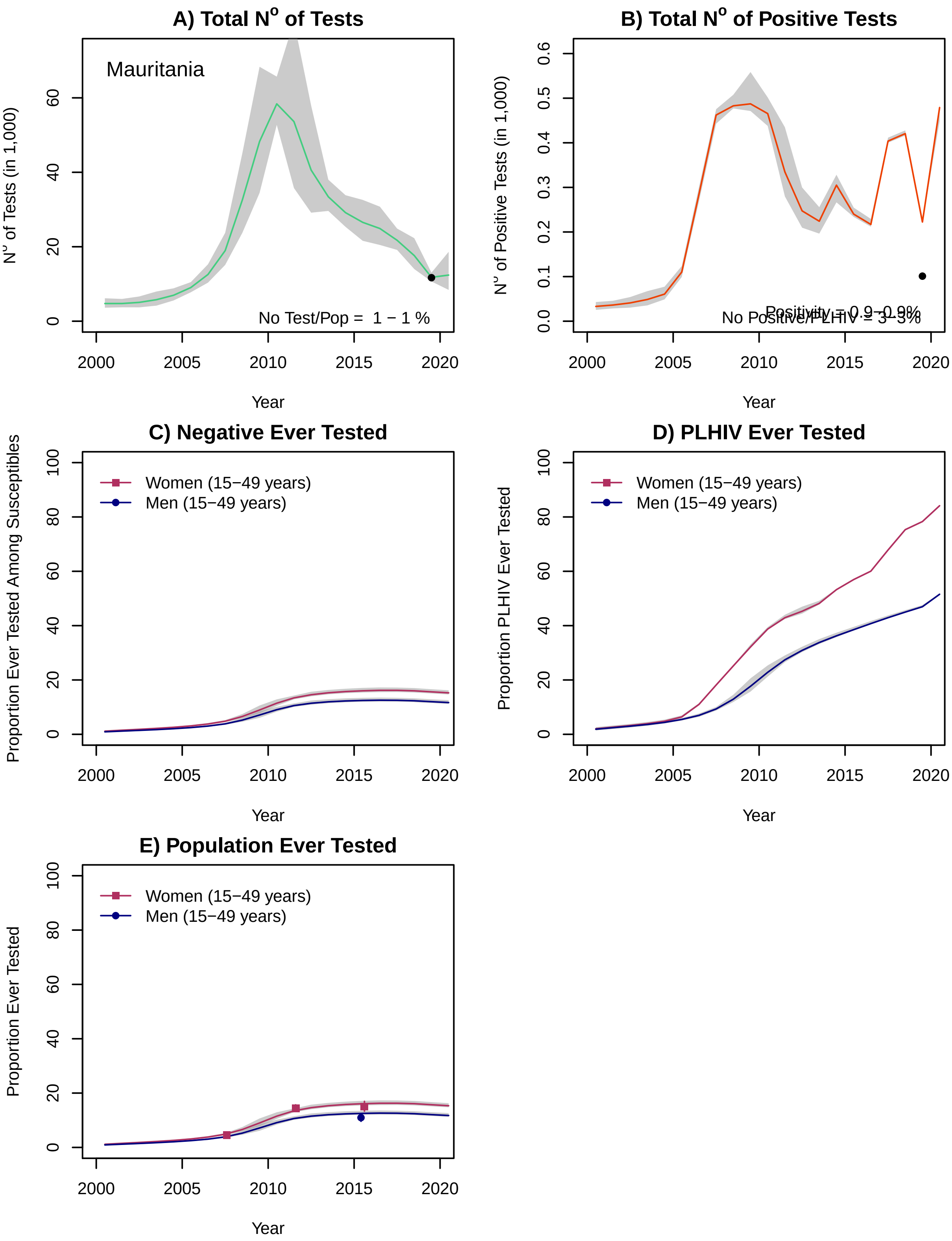
A) Total N° of Tests
Mauritania
N° of Tests (in 1,000)
No Test/Pop = 1 − 1 %
Year
B) Total N° of Positive Tests
N° of Positive Tests (in 1,000)
Positivity = 0.9−0.9%
No Positive/PLHIV = 3−3%
Year
C) Negative Ever Tested
Proportion Ever Tested Among Susceptibles
Women (15−49 years)
Men (15−49 years)
Year
D) PLHIV Ever Tested
Proportion PLHIV Ever Tested
Women (15−49 years)
Men (15−49 years)
Year
E) Population Ever Tested
Proportion Ever Tested
Women (15−49 years)
Men (15−49 years)
Year

### Mozambique

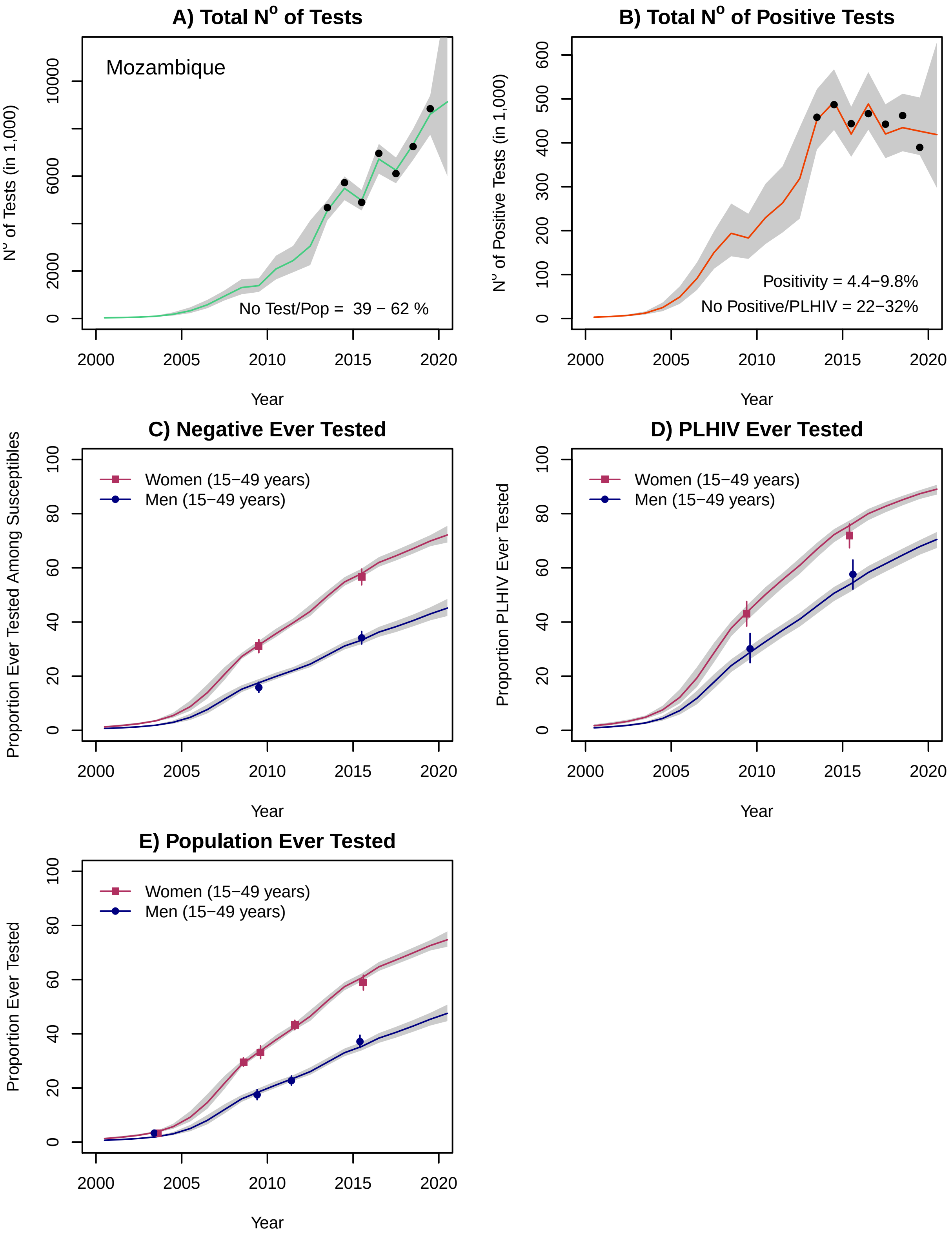

A) Total Nº of Tests
Mozambique
No Test/Pop = 39 − 62 %
Nº of Tests (in 1,000)
Year
B) Total Nº of Positive Tests
Positivity = 4.4−9.8%
No Positive/PLHIV = 22−32%
Nº of Positive Tests (in 1,000)
Year
C) Negative Ever Tested
Women (15−49 years)
Men (15−49 years)
Proportion Ever Tested Among Susceptibles
Year
D) PLHIV Ever Tested
Women (15−49 years)
Men (15−49 years)
Proportion PLHIV Ever Tested
Year
E) Population Ever Tested
Women (15−49 years)
Men (15−49 years)
Proportion Ever Tested
Year

#### Namibia*

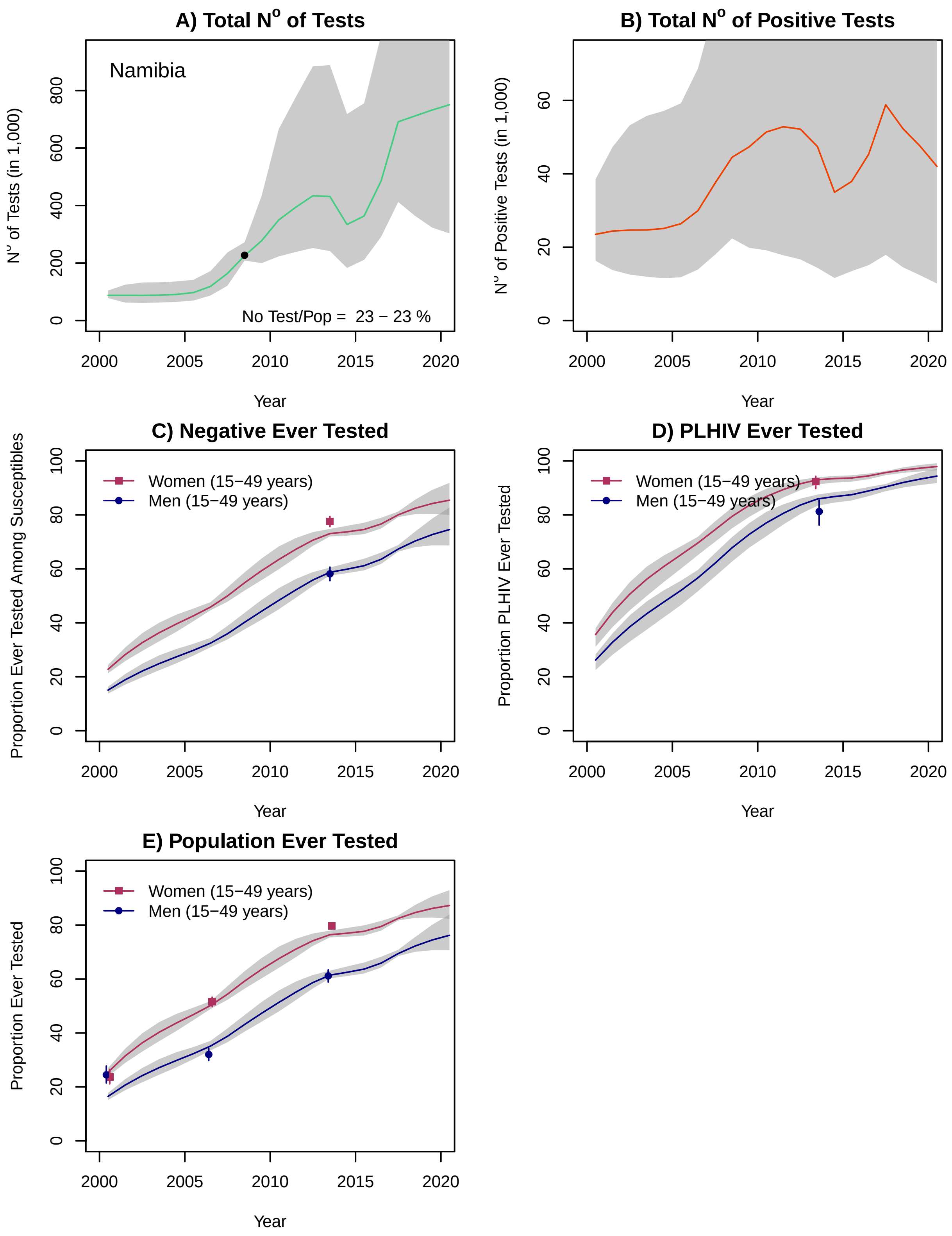
A) Total N° of Tests
Namibia
No Test/Pop = 23 − 23 %
N° of Tests (in 1,000)
Year
B) Total N° of Positive Tests
N° of Positive Tests (in 1,000)
Year
C) Negative Ever Tested
Women (15−49 years)
Men (15−49 years)
Proportion Ever Tested Among Susceptibles
Year
D) PLHIV Ever Tested
Women (15−49 years)
Men (15−49 years)
Proportion PLHIV Ever Tested
Year
E) Population Ever Tested
Women (15−49 years)
Men (15−49 years)
Proportion Ever Tested
Year

### Niger

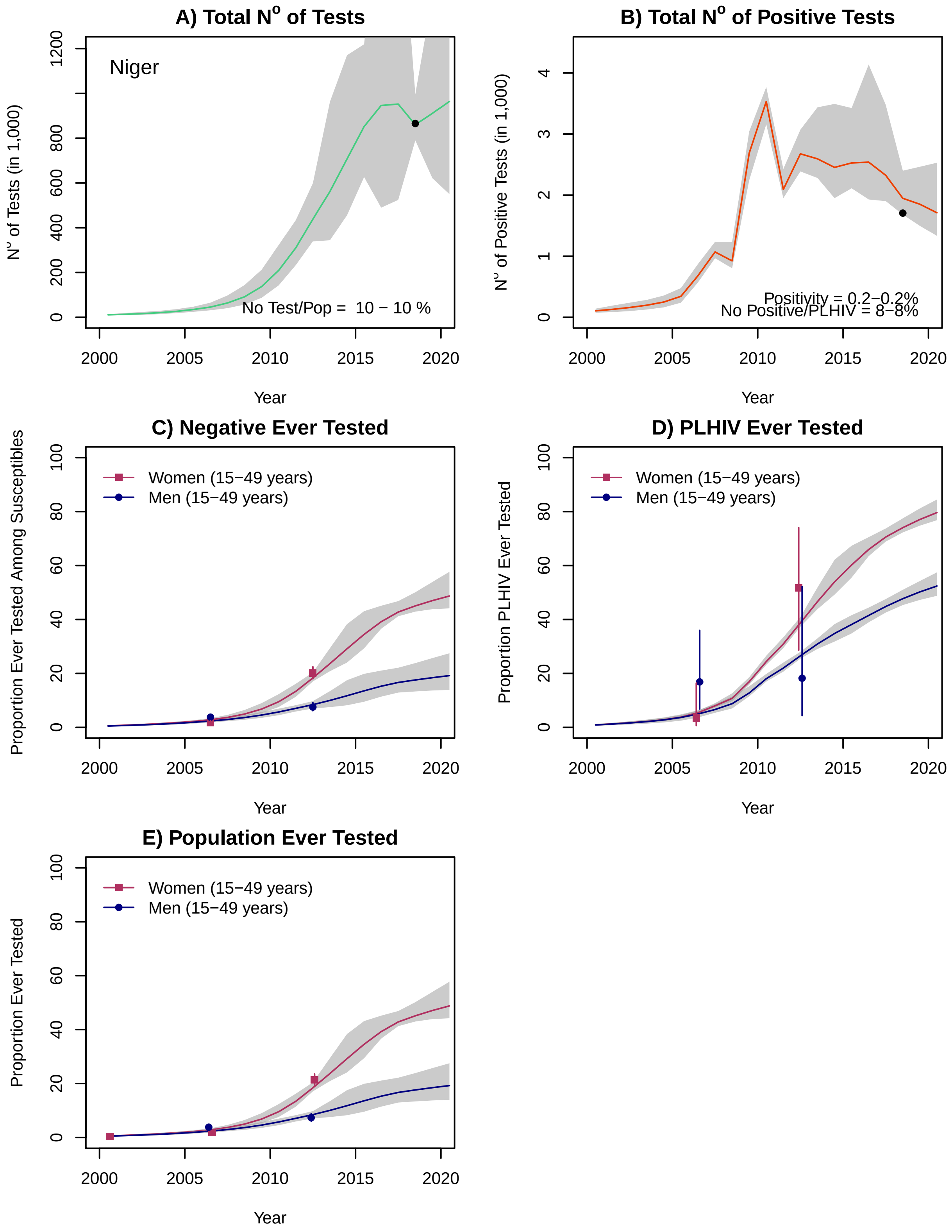
A) Total N° of Tests
Niger
No Test/Pop = 10 − 10 %
N° of Tests (in 1,000)
Year
B) Total N° of Positive Tests
Positivity = 0.2−0.2%
No Positive/PLHIV = 8−8%
N° of Positive Tests (in 1,000)
Year
C) Negative Ever Tested
Women (15−49 years)
Men (15−49 years)
Proportion Ever Tested Among Susceptibles
Year
D) PLHIV Ever Tested
Women (15−49 years)
Men (15−49 years)
Proportion PLHIV Ever Tested
Year
E) Population Ever Tested
Women (15−49 years)
Men (15−49 years)
Proportion Ever Tested
Year

### Nigeria

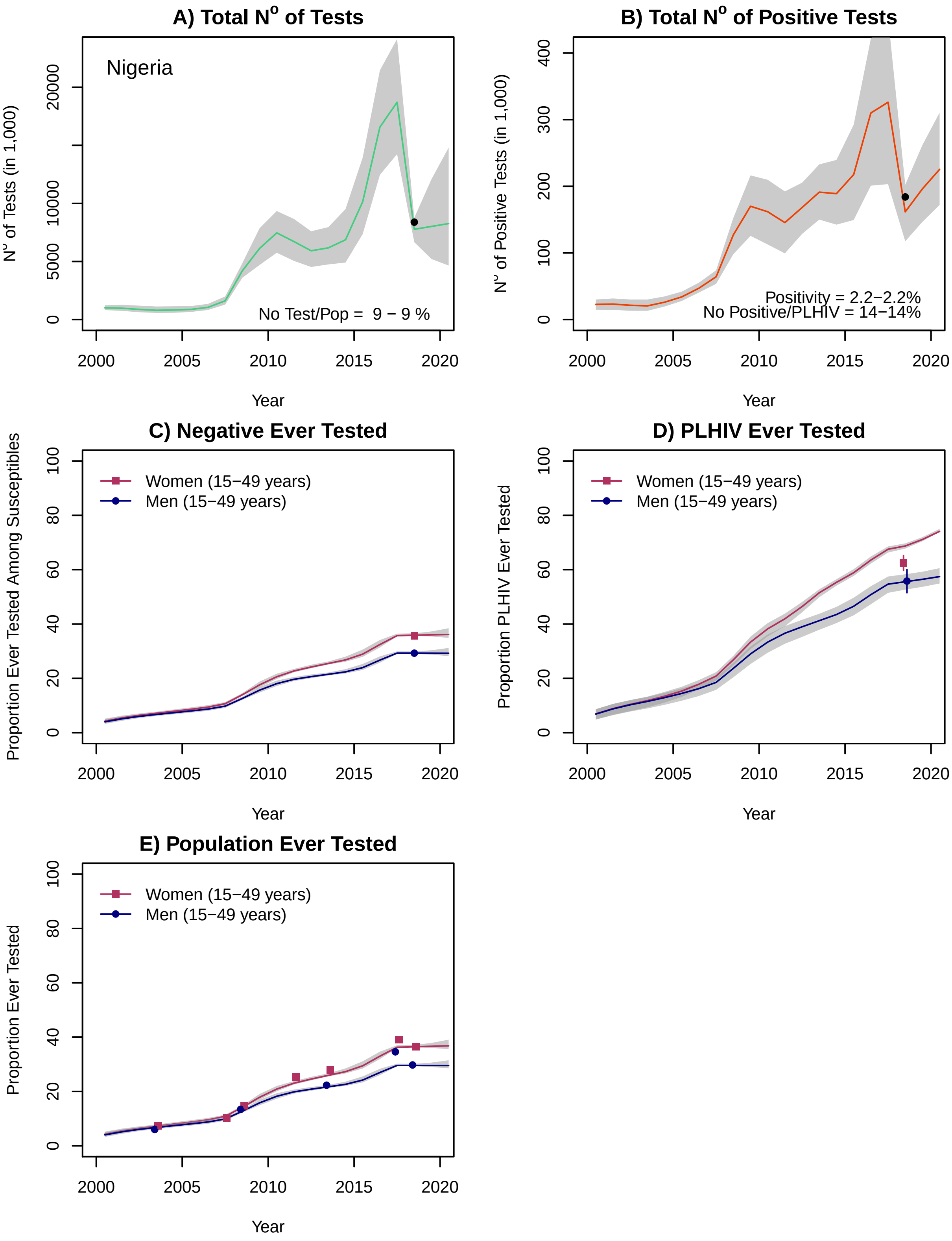
A) Total N° of Tests
Nigeria
N° of Tests (in 1,000)
No Test/Pop = 9 − 9 %
0
5000
10000
20000
2000
2005
2010
2015
2020
Year
B) Total N° of Positive Tests
N° of Positive Tests (in 1,000)
Positivity = 2.2−2.2%
No Positive/PLHIV = 14−14%
0
100
200
300
400
Year
C) Negative Ever Tested
Proportion Ever Tested Among Susceptibles
Women (15−49 years)
Men (15−49 years)
0
20
40
60
80
100
Year
D) PLHIV Ever Tested
Proportion PLHIV Ever Tested
Women (15−49 years)
Men (15−49 years)
Year
E) Population Ever Tested
Proportion Ever Tested
Women (15−49 years)
Men (15−49 years)
Year

### Rwanda*

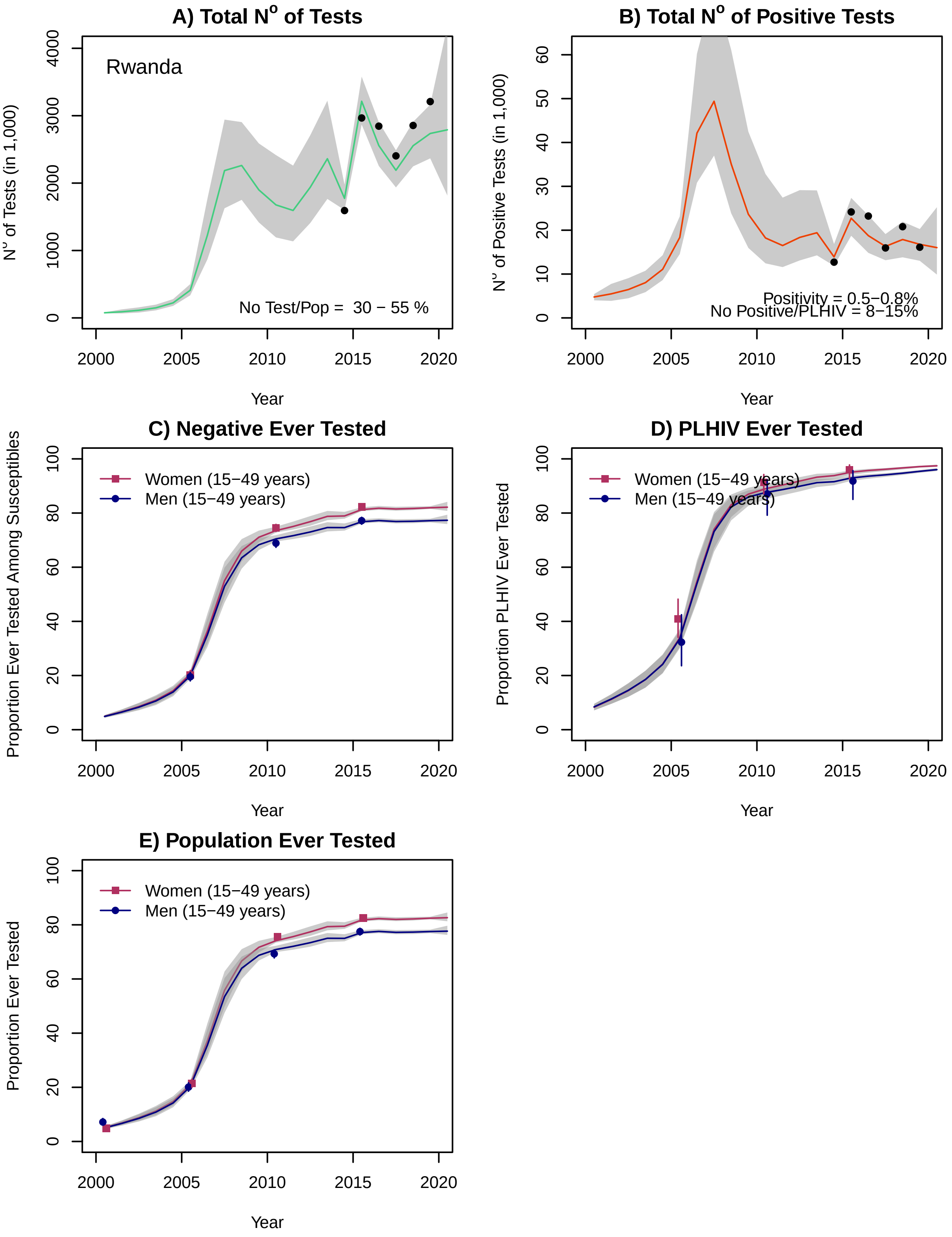
A) Total N° of Tests
Rwanda
N° of Tests (in 1,000)
No Test/Pop =  30 − 55 %
Year
B) Total N° of Positive Tests
N° of Positive Tests (in 1,000)
Positivity = 0.5−0.8%
No Positive/PLHIV = 8−15%
Year
C) Negative Ever Tested
Proportion Ever Tested Among Susceptibles
Women (15−49 years)
Men (15−49 years)
Year
D) PLHIV Ever Tested
Proportion PLHIV Ever Tested
Women (15−49 years)
Men (15−49 years)
Year
E) Population Ever Tested
Proportion Ever Tested
Women (15−49 years)
Men (15−49 years)
Year

### Senegal

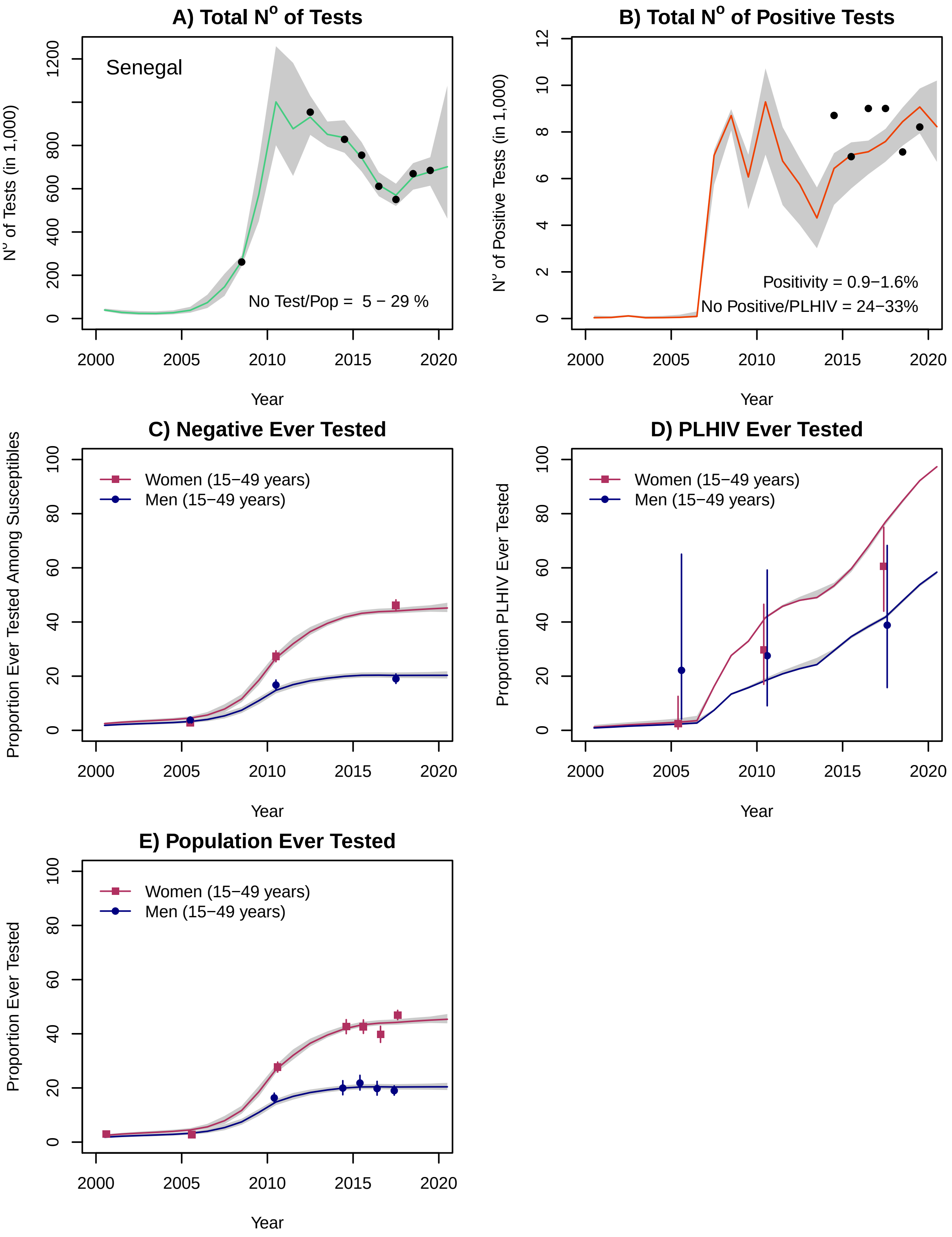

A) Total N° of Tests
Senegal
No Test/Pop = 5 − 29 %
N° of Tests (in 1,000)
Year
B) Total N° of Positive Tests
Positivity = 0.9−1.6%
No Positive/PLHIV = 24−33%
N° of Positive Tests (in 1,000)
Year
C) Negative Ever Tested
Women (15−49 years)
Men (15−49 years)
Proportion Ever Tested Among Susceptibles
Year
D) PLHIV Ever Tested
Women (15−49 years)
Men (15−49 years)
Proportion PLHIV Ever Tested
Year
E) Population Ever Tested
Women (15−49 years)
Men (15−49 years)
Proportion Ever Tested
Year

#### Sierra Leone

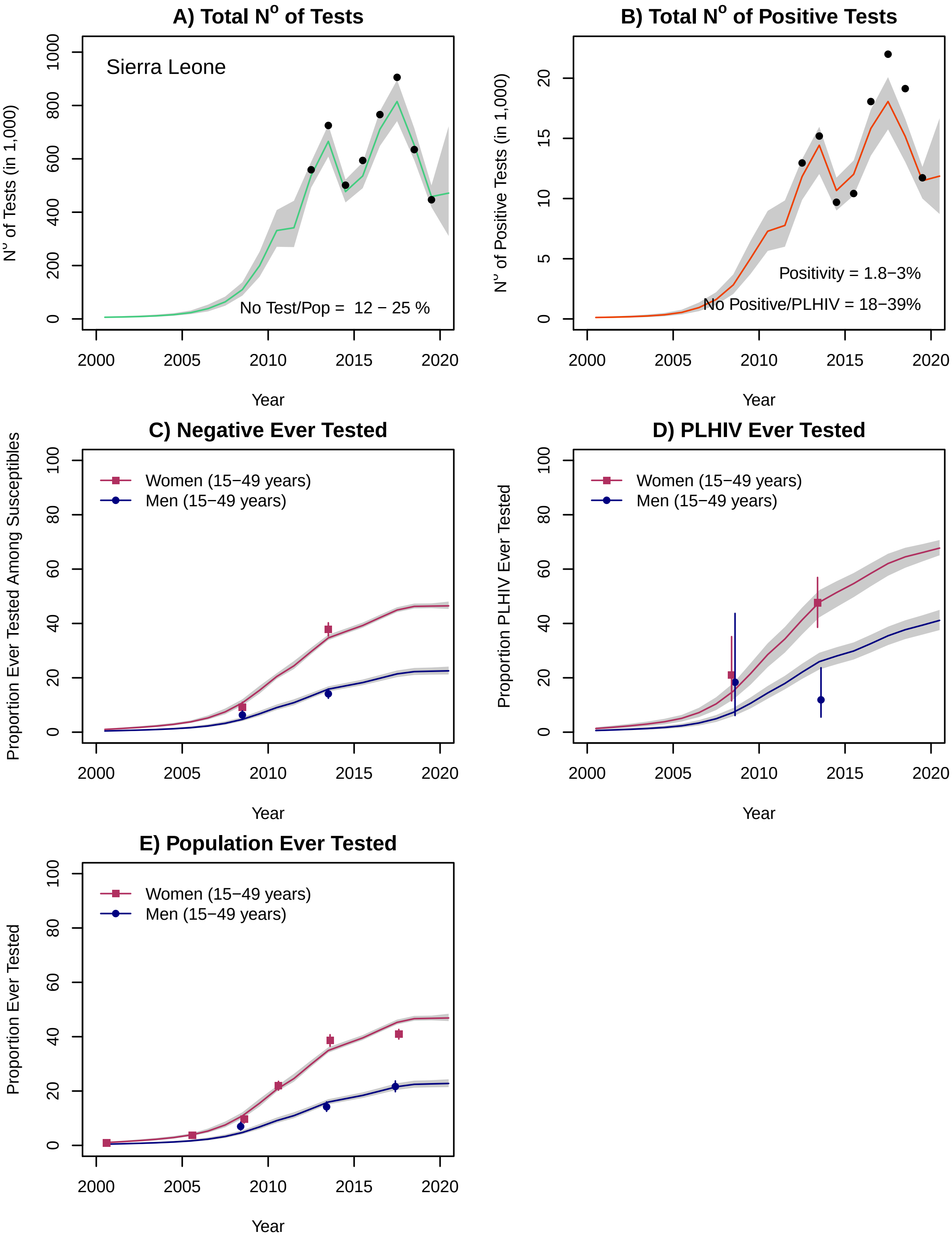

### South Africa

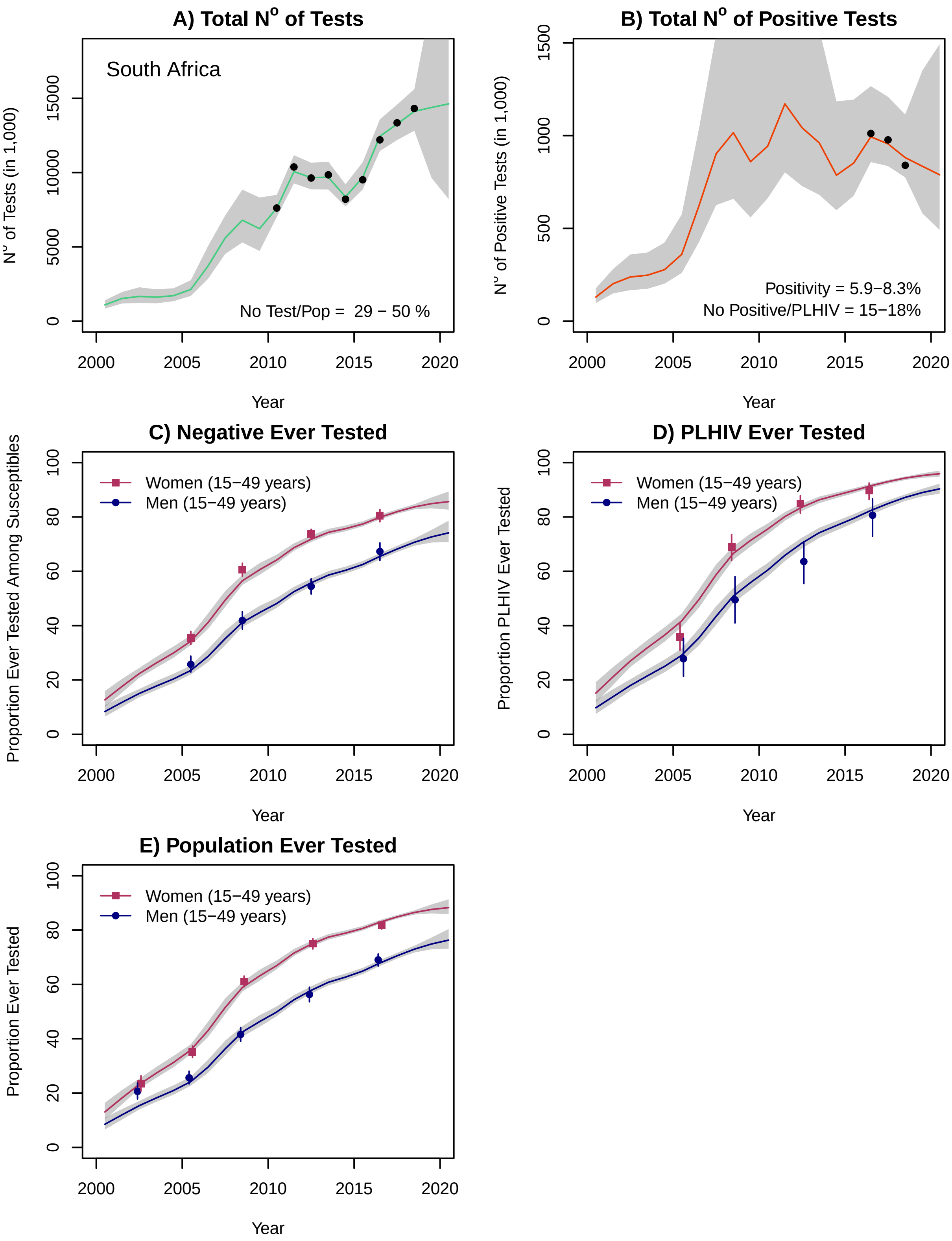

#### South Sudan

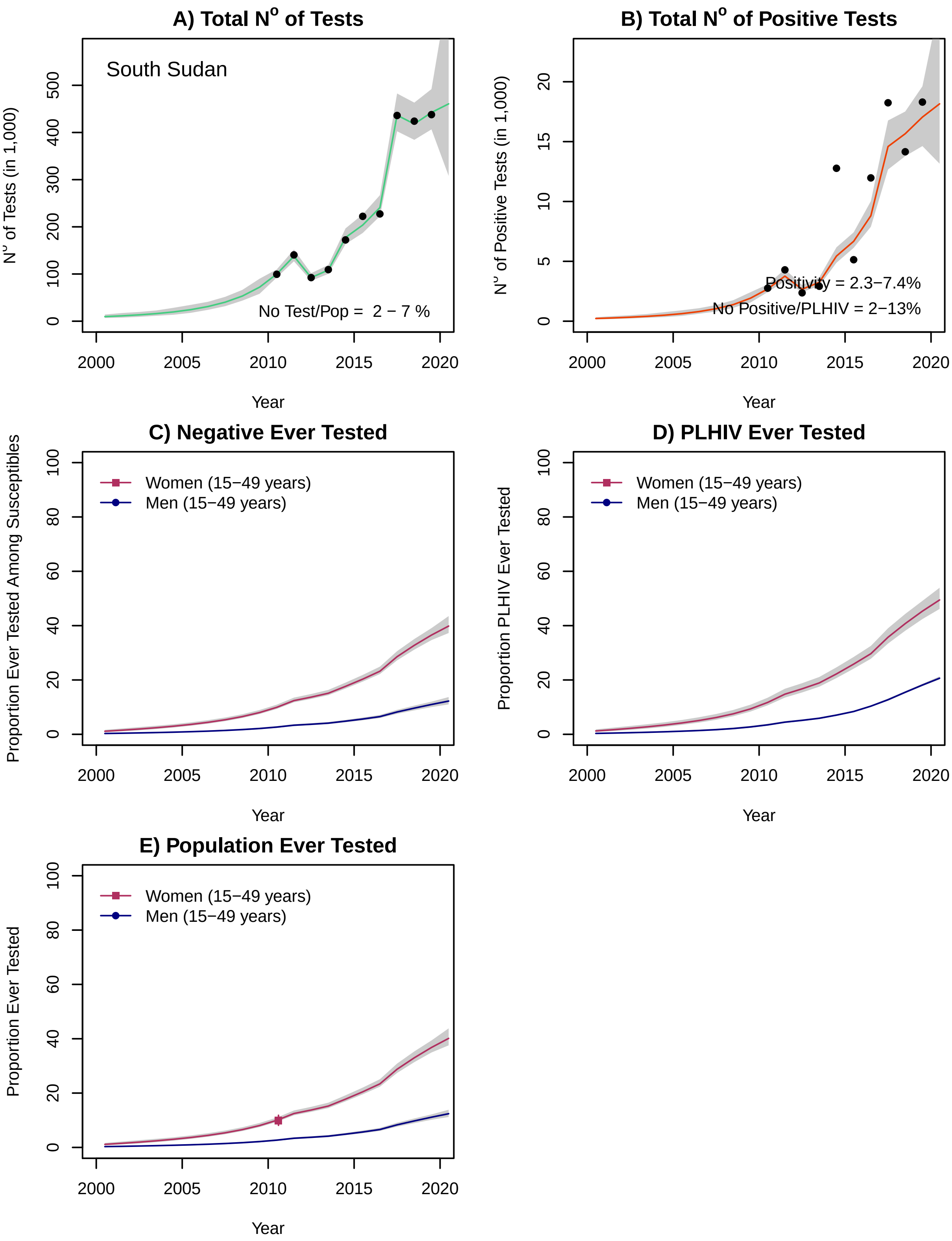

A) Total N° of Tests
South Sudan
N° of Tests (in 1,000)
No Test/Pop = 2 − 7 %
Year
B) Total N° of Positive Tests
N° of Positive Tests (in 1,000)
Positivity = 2.3−7.4%
No Positive/PLHIV = 2−13%
Year
C) Negative Ever Tested
Proportion Ever Tested Among Susceptibles
Women (15−49 years)
Men (15−49 years)
Year
D) PLHIV Ever Tested
Proportion PLHIV Ever Tested
Women (15−49 years)
Men (15−49 years)
Year
E) Population Ever Tested
Proportion Ever Tested
Women (15−49 years)
Men (15−49 years)
Year

### Tanzania

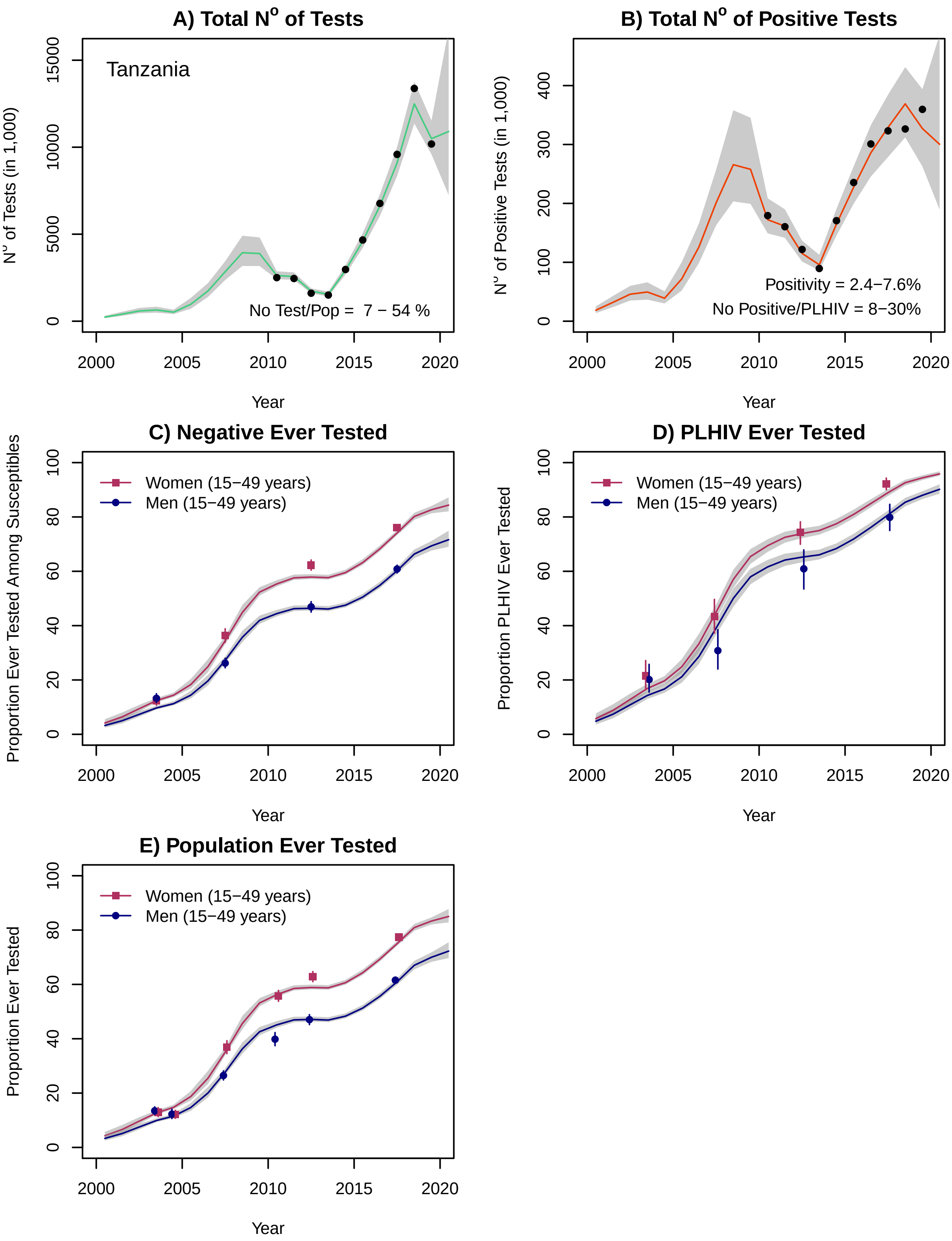

A) Total N° of Tests
Tanzania
N° of Tests (in 1,000)
No Test/Pop = 7 − 54 %
0
5000
10000
15000
2000
2005
2010
2015
2020
Year
B) Total N° of Positive Tests
N° of Positive Tests (in 1,000)
Positivity = 2.4−7.6%
No Positive/PLHIV = 8−30%
0
100
200
300
400
Year
C) Negative Ever Tested
Proportion Ever Tested Among Susceptibles
Women (15−49 years)
Men (15−49 years)
0
20
40
60
80
100
Year
D) PLHIV Ever Tested
Proportion PLHIV Ever Tested
Women (15−49 years)
Men (15−49 years)
Year
E) Population Ever Tested
Proportion Ever Tested
Women (15−49 years)
Men (15−49 years)
Year

### Togo

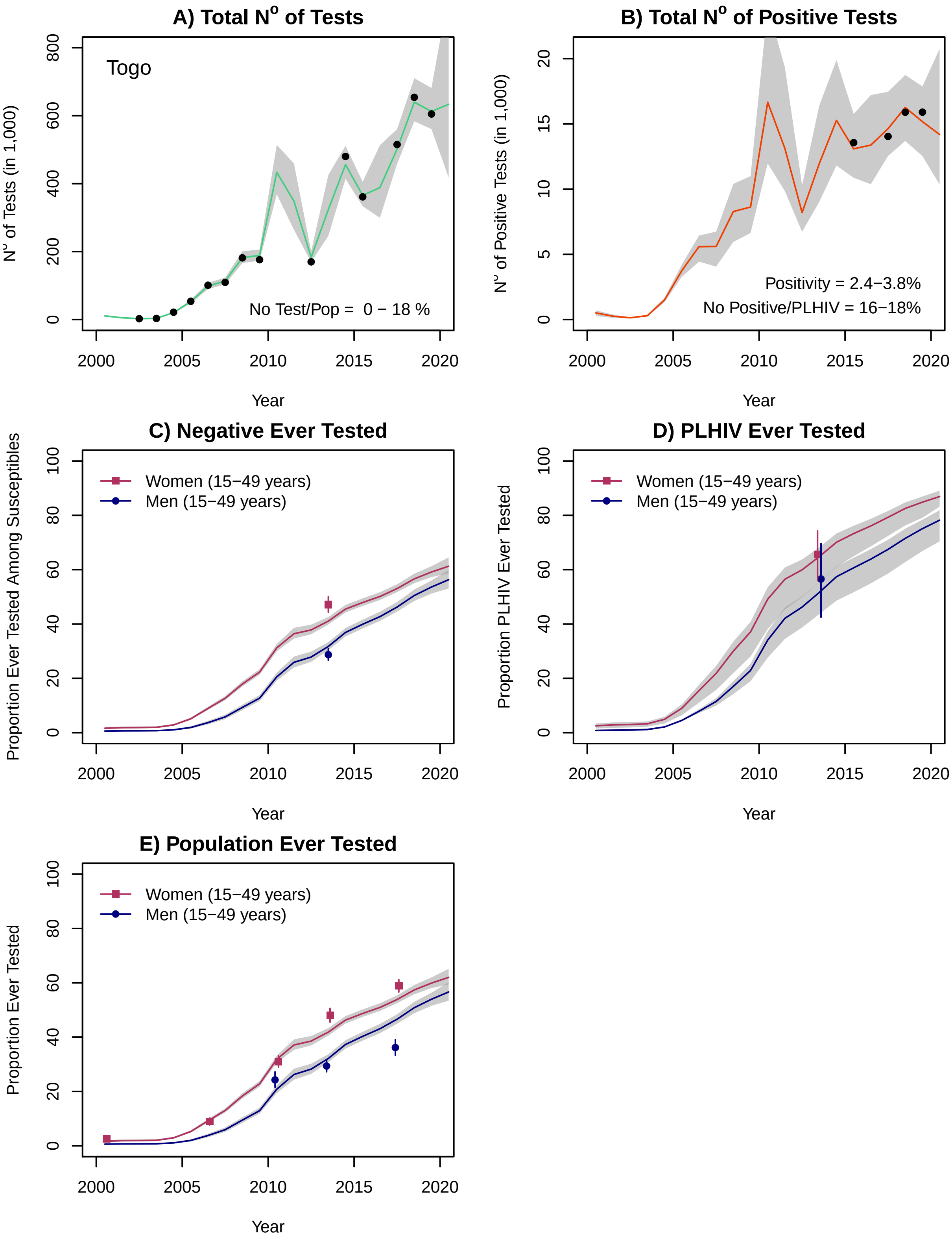
A) Total N° of Tests
Togo
No Test/Pop = 0 − 18 %
N° of Tests (in 1,000)
0
200
400
600
800
2000
2005
2010
2015
2020
Year
B) Total N° of Positive Tests
Positivity = 2.4−3.8%
No Positive/PLHIV = 16−18%
N° of Positive Tests (in 1,000)
0
5
10
15
20
Year
C) Negative Ever Tested
Women (15−49 years)
Men (15−49 years)
Proportion Ever Tested Among Susceptibles
0
20
40
60
80
100
Year
D) PLHIV Ever Tested
Women (15−49 years)
Men (15−49 years)
Proportion PLHIV Ever Tested
Year
E) Population Ever Tested
Women (15−49 years)
Men (15−49 years)
Proportion Ever Tested
Year

### Uganda*

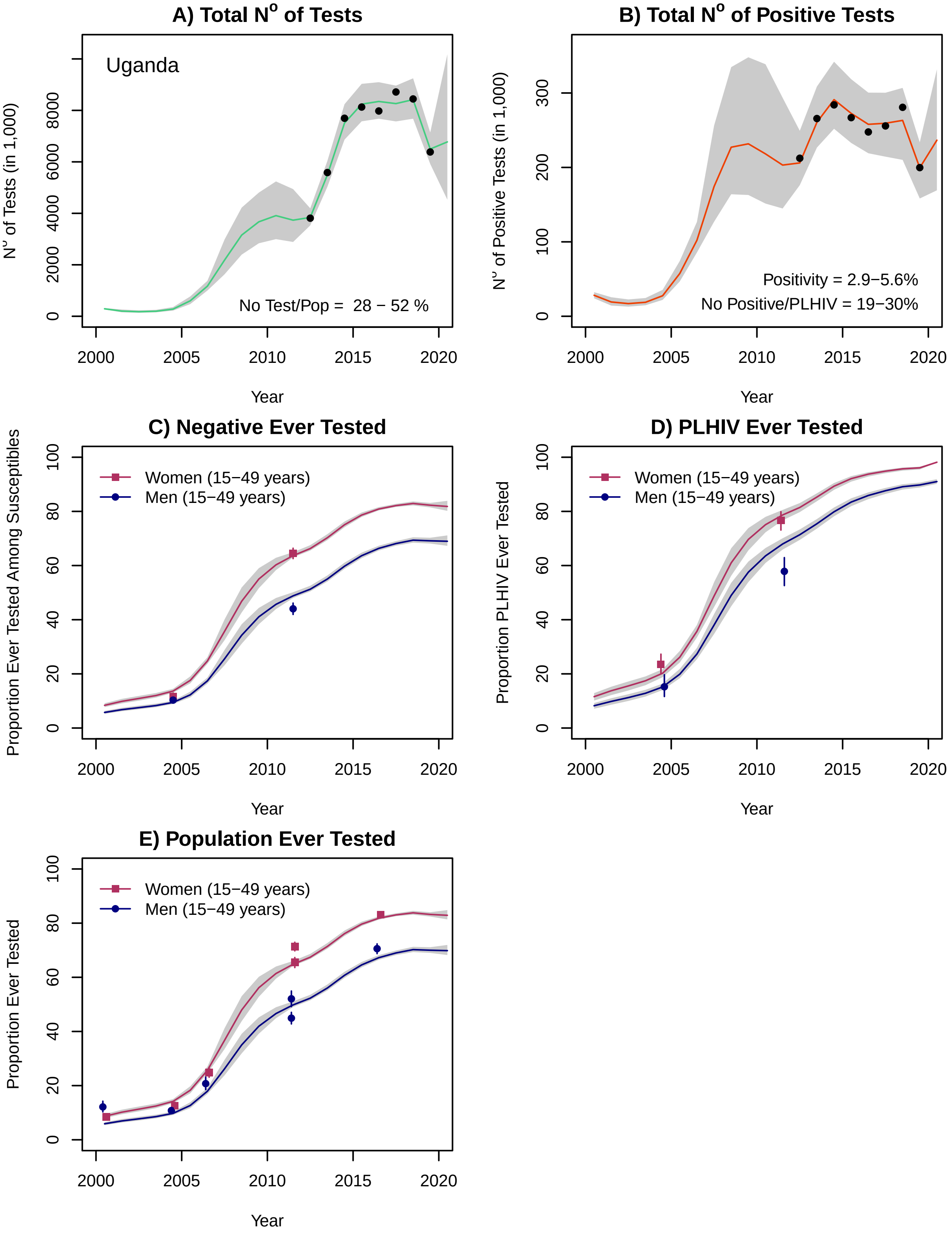
A) Total N° of Tests
Uganda
N° of Tests (in 1,000)
0
2000
4000
6000
8000
No Test/Pop = 28 − 52 %
2000
2005
2010
2015
2020
Year
B) Total N° of Positive Tests
N° of Positive Tests (in 1,000)
0
100
200
300
Positivity = 2.9−5.6%
No Positive/PLHIV = 19−30%
Year
C) Negative Ever Tested
Proportion Ever Tested Among Susceptibles
Women (15−49 years)
Men (15−49 years)
0
20
40
60
80
100
Year
D) PLHIV Ever Tested
Proportion PLHIV Ever Tested
Women (15−49 years)
Men (15−49 years)
Year
E) Population Ever Tested
Proportion Ever Tested
Women (15−49 years)
Men (15−49 years)
Year

### Zambia

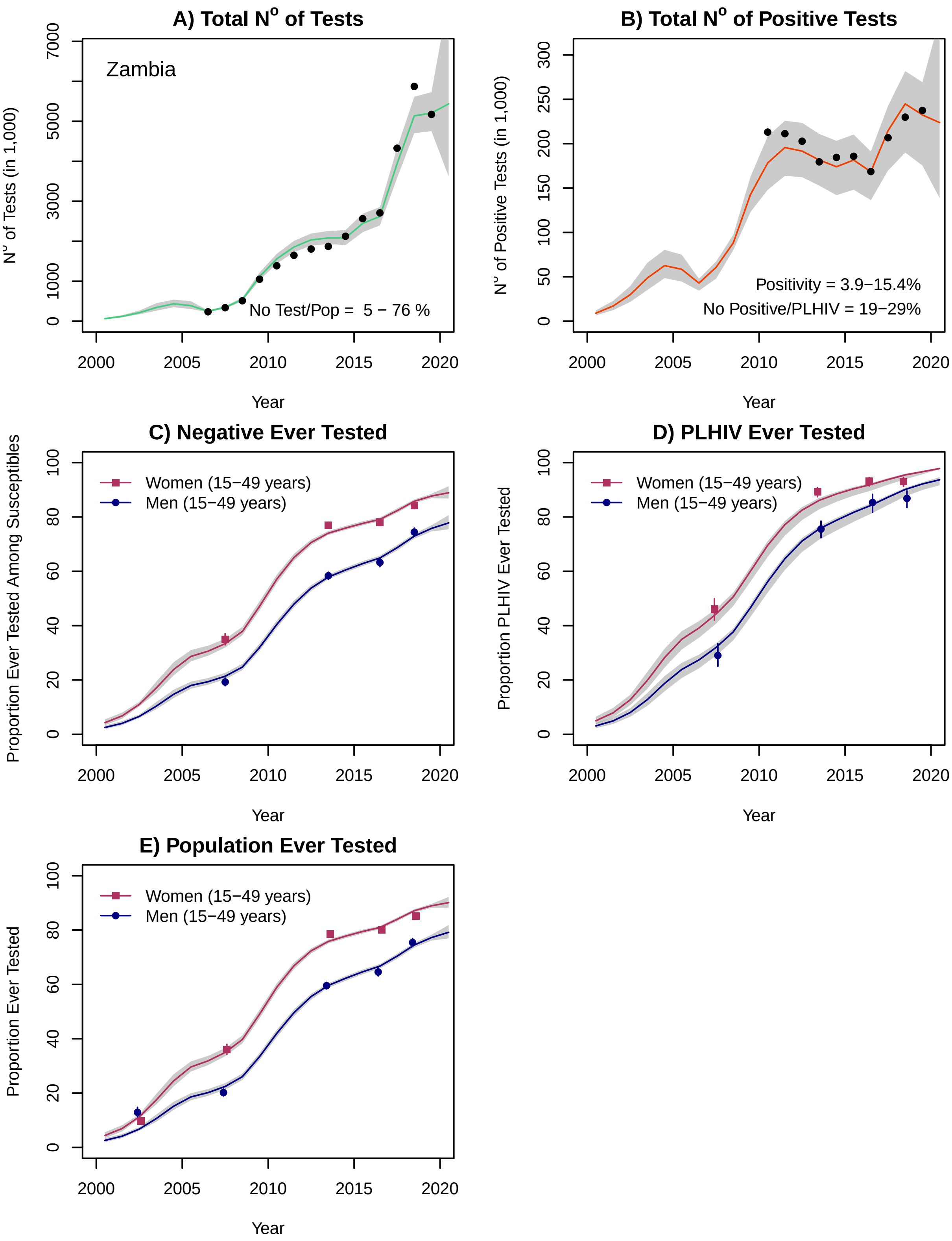

A) Total N° of Tests
Zambia
No Test/Pop = 5 − 76 %
N° of Tests (in 1,000)
Year
B) Total N° of Positive Tests
Positivity = 3.9−15.4%
No Positive/PLHIV = 19−29%
N° of Positive Tests (in 1,000)
Year
C) Negative Ever Tested
Women (15−49 years)
Men (15−49 years)
Proportion Ever Tested Among Susceptibles
Year
D) PLHIV Ever Tested
Women (15−49 years)
Men (15−49 years)
Proportion PLHIV Ever Tested
Year
E) Population Ever Tested
Women (15−49 years)
Men (15−49 years)
Proportion Ever Tested
Year

### Zimbabwe*

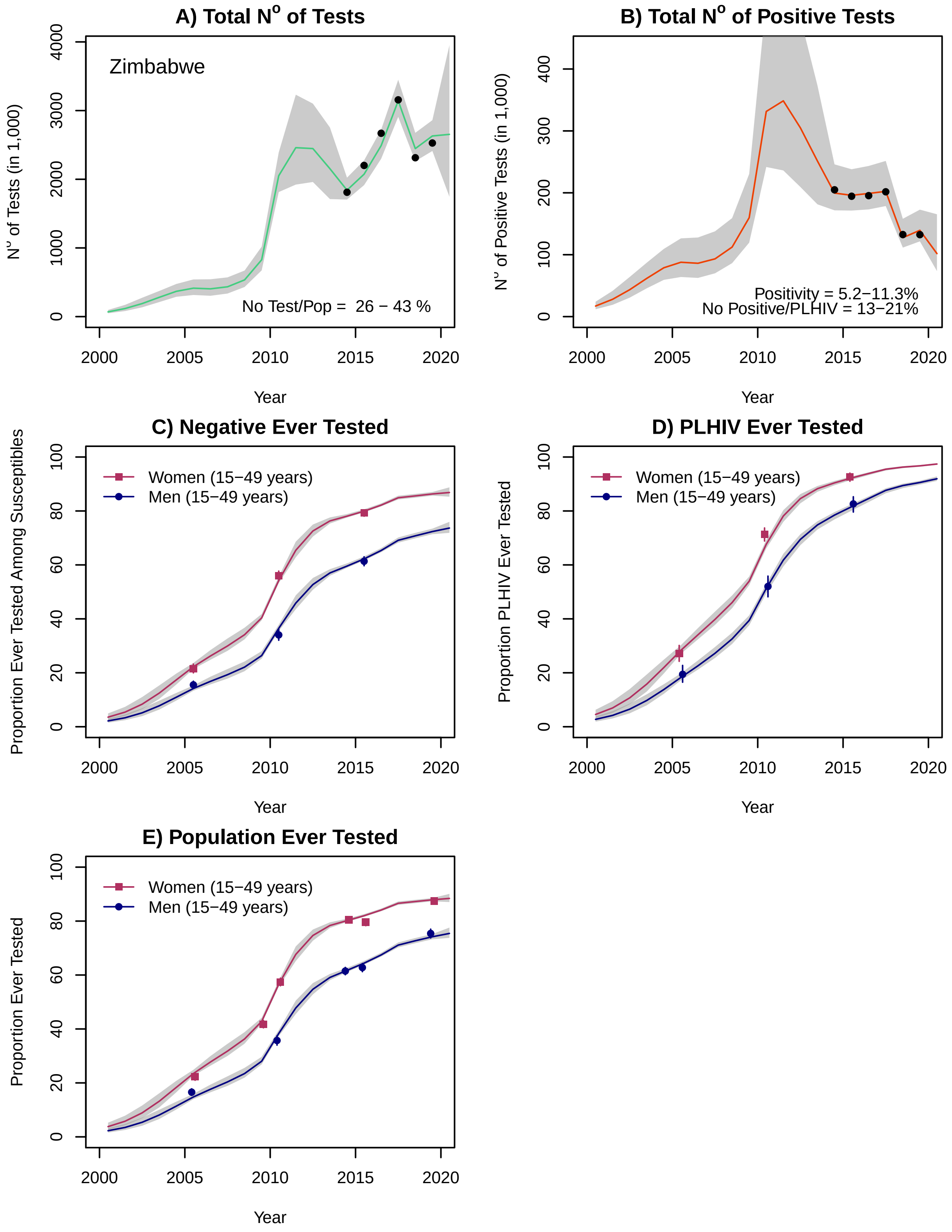

A) Total N° of Tests
Zimbabwe
No Test/Pop = 26 − 43 %
N° of Tests (in 1,000)
Year
B) Total N° of Positive Tests
Positivity = 5.2−11.3%
No Positive/PLHIV = 13−21%
N° of Positive Tests (in 1,000)
Year
C) Negative Ever Tested
Women (15−49 years)
Men (15−49 years)
Proportion Ever Tested Among Susceptibles
Year
D) PLHIV Ever Tested
Women (15−49 years)
Men (15−49 years)
Proportion PLHIV Ever Tested
Year
E) Population Ever Tested
Women (15−49 years)
Men (15−49 years)
Proportion Ever Tested
Year

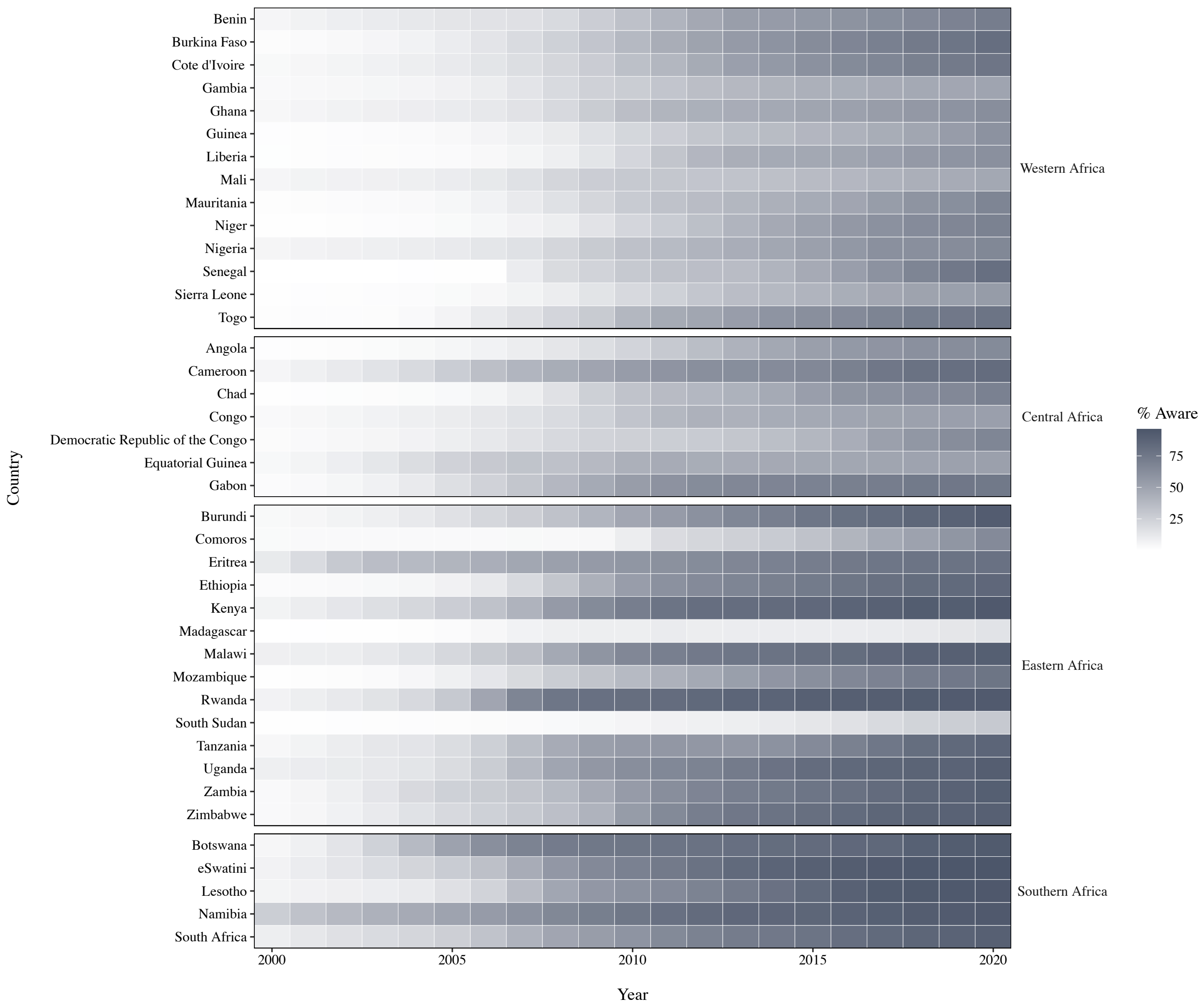


***Figure S1*****: National progress in knowledge of HIV status in sub-Saharan Africa, 2000-2020**

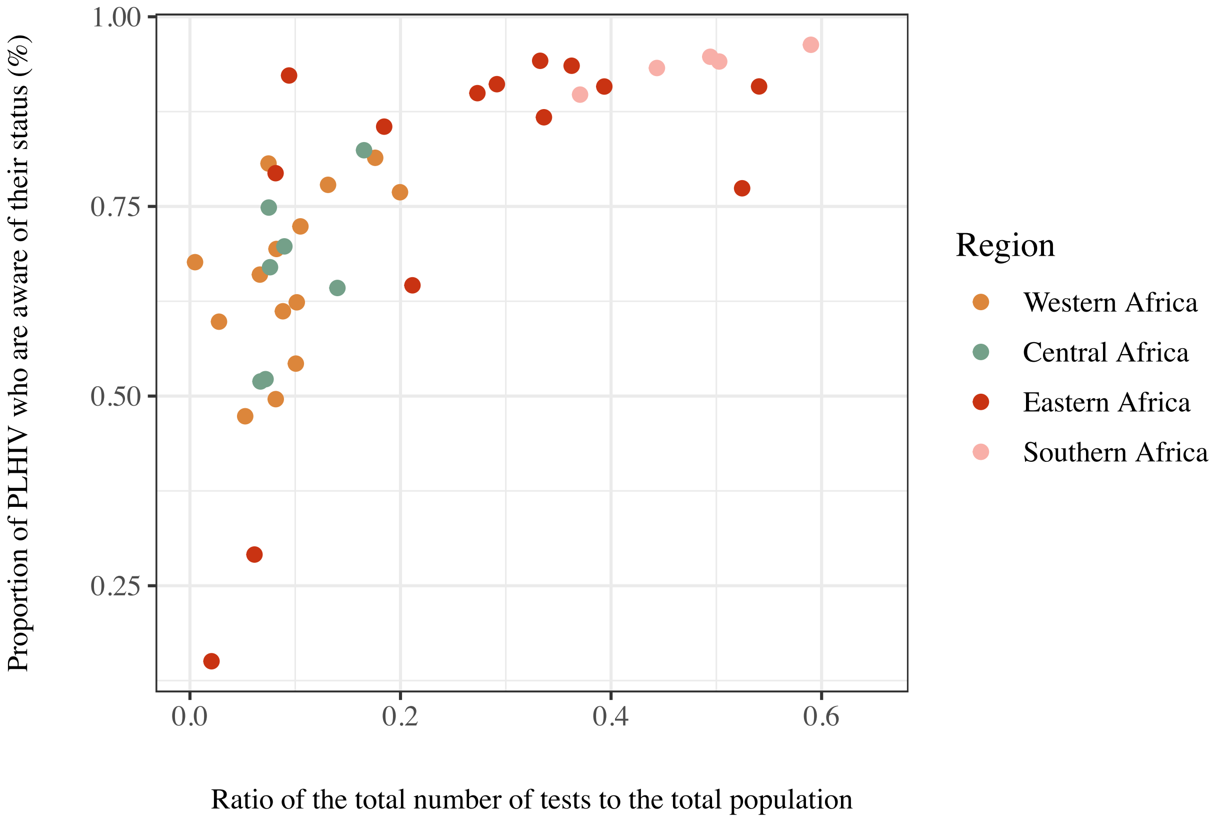


***Figure S2*: Correlation between knowledge of HIV status and ratio of total number of tests relatives to the population aged ≥15 years in sub-Saharan Africa, 2020**
Each point represents a different country (n = 40).

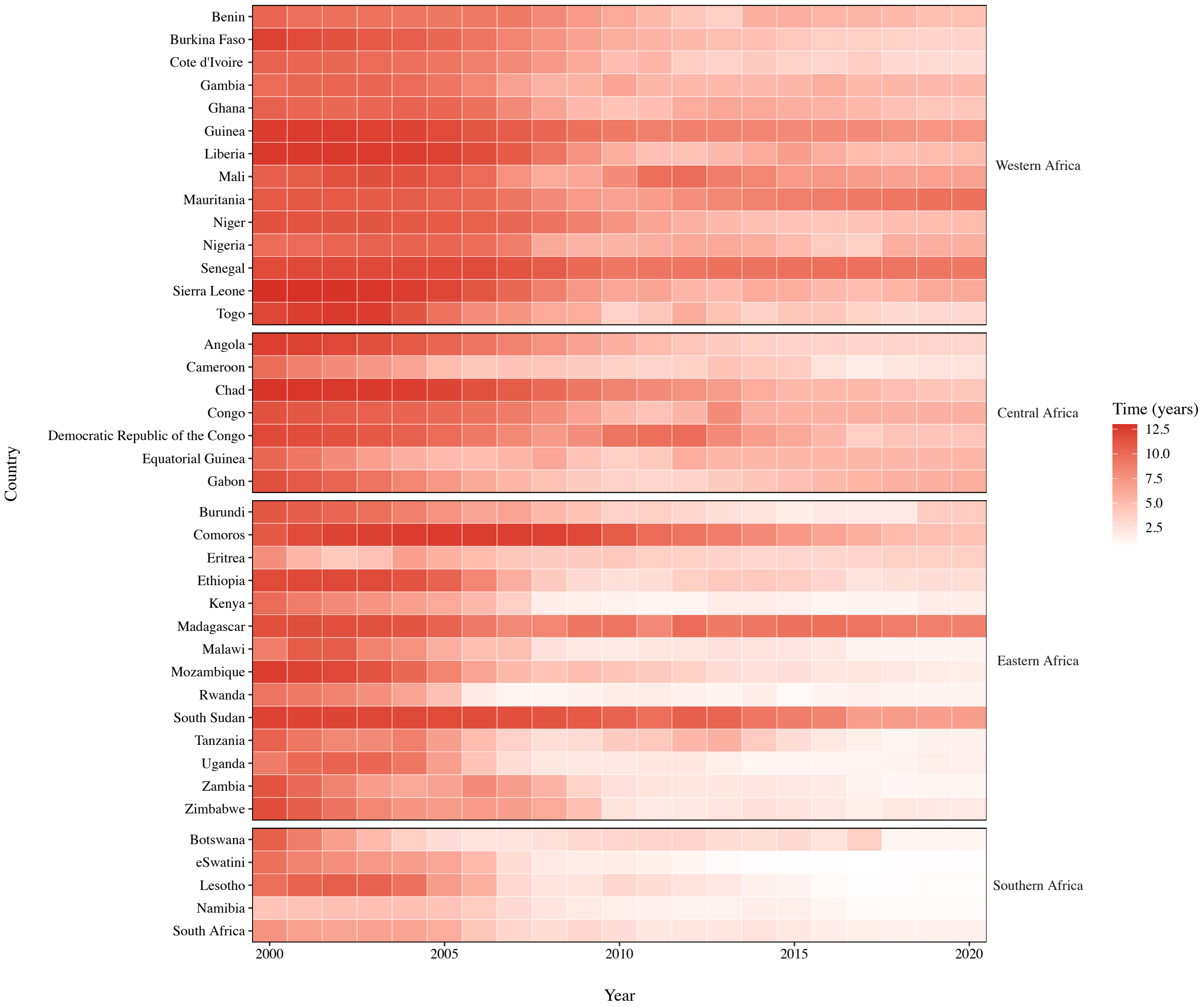


***Figure S3*: National progress in median time to diagnosis or AIDS-related death in sub-Saharan Africa, 2000-2020**

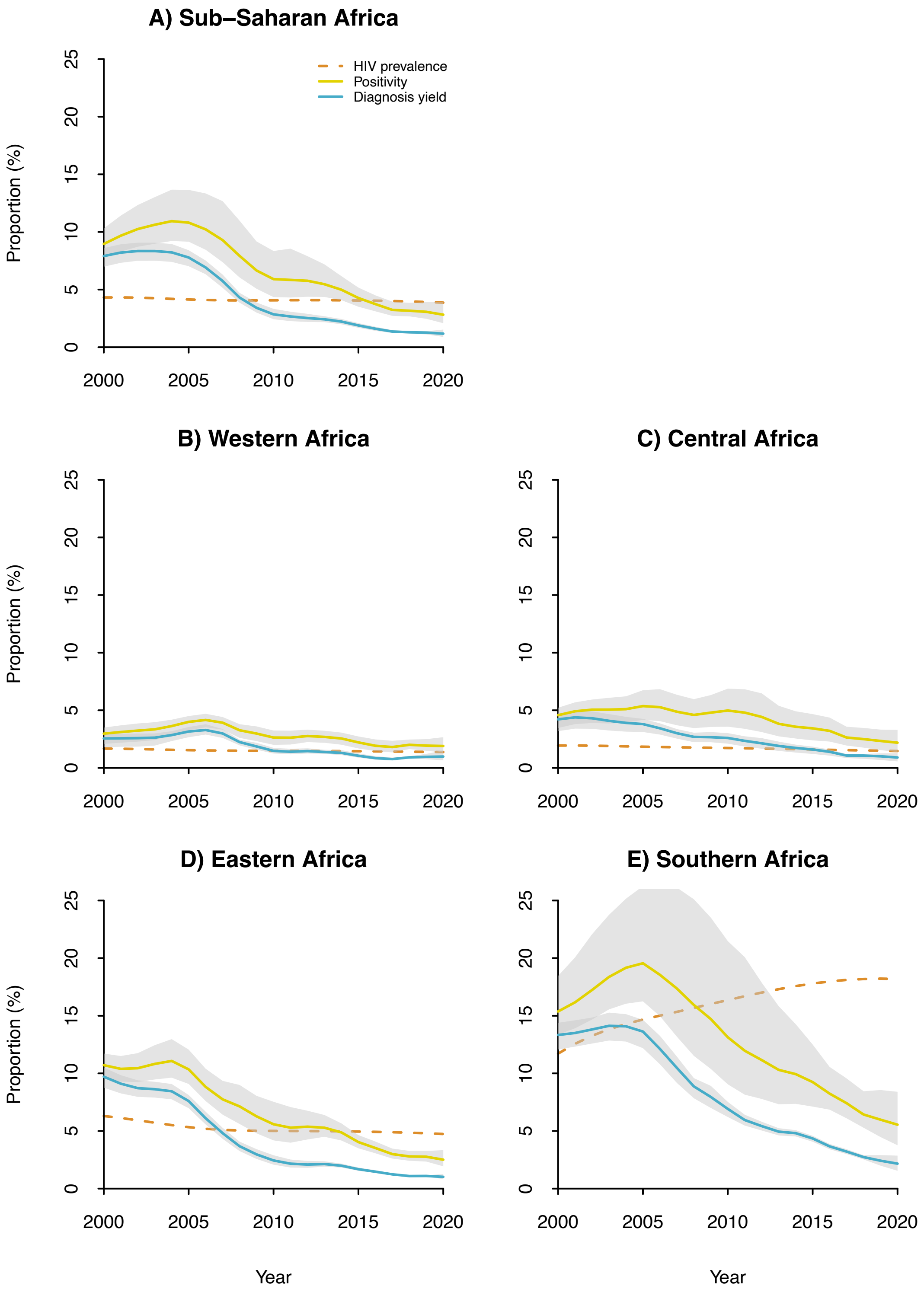


***Figure S4*: Regional trends in positivity and diagnosis yield, 2000-2020**

The positivity corresponds to the proportion of positive tests among all tests, while diagnosis yield corresponds to the proportion of new diagnoses among all tests. HIV prevalence is expressed among the total population aged 15-99 years. The shaded areas correspond to the 95% credible intervals

*Table S1*: **Supplemental HIV testing services programme data sources**

| Country | Year | Source |
|---|---|---|
| Burundi | 2003 | Conseil National de Lutte contre le SIDA, Rapport d'activités sur la lutte contre le SIDA et rapport sur les progrès enregistrés vers un accès universel (Burundi), 2015. Available from: https://www.unaids.org/en/dataanalysis/knowyourresponse/countryprogressreports/2015countries#B |
| | 2004 | Conseil National de Lutte contre le SIDA, Rapport d'activités sur la lutte contre le SIDA et rapport sur les progrès enregistrés vers un accès universel (Burundi), 2015. Available from: https://www.unaids.org/en/dataanalysis/knowyourresponse/countryprogressreports/2015countries#B |
| | 2005 | Conseil National de Lutte contre le SIDA, Rapport d'activités sur la lutte contre le SIDA et rapport sur les progrès enregistrés vers un accès universel (Burundi), 2015. Available from: https://www.unaids.org/en/dataanalysis/knowyourresponse/countryprogressreports/2015countries#B |
| | 2006 | Conseil National de Lutte contre le SIDA, Rapport d'activités sur la lutte contre le SIDA et rapport sur les progrès enregistrés vers un accès universel (Burundi), 2015. Available from: https://www.unaids.org/en/dataanalysis/knowyourresponse/countryprogressreports/2015countries#B |
| | 2007 | Conseil National de Lutte contre le SIDA, Rapport d'activités sur la lutte contre le SIDA et rapport sur les progrès enregistrés vers un accès universel (Burundi), 2015. Available from: https://www.unaids.org/en/dataanalysis/knowyourresponse/countryprogressreports/2015countries#B |
| | 2008 | Conseil National de Lutte contre le SIDA, Rapport d'activités sur la lutte contre le SIDA et rapport sur les progrès enregistrés vers un accès universel (Burundi), 2015. Available from: https://www.unaids.org/en/dataanalysis/knowyourresponse/countryprogressreports/2015countries#B |
| | 2009 | Conseil National de Lutte contre le SIDA, Rapport d'activités sur la lutte contre le SIDA et rapport sur les progrès enregistrés vers un accès universel (Burundi), 2015. Available from: https://www.unaids.org/en/dataanalysis/knowyourresponse/countryprogressreports/2015countries#B |
| Cameroon | 2013 | Rapport national de suivi de la déclaration politique sur le VIH/SIDA Cameroun – Global AIDS Response Progress (GARP), 2014. Available from: https://www.unaids.org/en/dataanalysis/knowyourresponse/countryprogressreports/2014countries#C |
| | 2014 | Ministry of Public Health, Plan d'accélération de la thérapie ARV au Cameroun 2016-2018, 2015. |
| Comoros | 2018 | UNAIDS/WHO/UNICEF. Global AIDS Monitoring, 2020. Accessed at: https://aidsinfo.unaids.org/ |
| Congo | 2013 | Conseil National de Lutte contre le SIDA, Rapport d'activités de la riposte au VIH/SIDA – 2013, 2014. Available from: https://www.unaids.org/en/dataanalysis/knowyourresponse/countryprogressreports/2014countries#C |
| Equatorial Guinea | 2008 | Programa nacional de lucha contra el SIDA, Declaracion de compromiso sobre VIH-SIDA, UNGASS, 2010. Available from: https://www.unaids.org/en/dataanalysis/knowyourresponse/countryprogressreports/2010countries#E |
| | 2012 | Programa nacional de lucha contra el SIDA, informe national sobre los progresos realizados en la lucha contra el VIH/SIDA Guinea Ecuatorial, 2014. Available from: https://www.unaids.org/en/dataanalysis/knowyourresponse/countryprogressreports/2014countries#E |
| | 2013 | Programa nacional de lucha contra el SIDA, informe national sobre los progresos realizados en la lucha contra el VIH/SIDA Guinea Ecuatorial, 2014. Available from: https://www.unaids.org/en/dataanalysis/knowyourresponse/countryprogressreports/2014countries#E |
| Eritrea | 2004 | National AIDS and Tuberculosis Control Division (NATCoD), 2012. Available from: https://www.unaids.org/en/dataanalysis/knowyourresponse/countryprogressreports/2012countries#E |
| | 2005 | National AIDS and Tuberculosis Control Division (NATCoD), 2012. Available from: https://www.unaids.org/en/dataanalysis/knowyourresponse/countryprogressreports/2012countries#E |
| | 2006 | National AIDS and Tuberculosis Control Division (NATCoD), 2012. Available from: https://www.unaids.org/en/dataanalysis/knowyourresponse/countryprogressreports/2012countries#E |
| | 2007 | National AIDS and Tuberculosis Control Division (NATCoD), 2012. Available from: https://www.unaids.org/en/dataanalysis/knowyourresponse/countryprogressreports/2012countries#E |
| | 2008 | National AIDS and Tuberculosis Control Division (NATCoD), 2012. Available from: https://www.unaids.org/en/dataanalysis/knowyourresponse/countryprogressreports/2012countries#E |
| | 2009 | National AIDS and Tuberculosis Control Division (NATCoD), 2012. Available from: https://www.unaids.org/en/dataanalysis/knowyourresponse/countryprogressreports/2012countries#E |
| | 2010 | National AIDS and Tuberculosis Control Division (NATCoD), 2012. Available from: https://www.unaids.org/en/dataanalysis/knowyourresponse/countryprogressreports/2012countries#E |
| | 2011 | National AIDS and Tuberculosis Control Division (NATCoD), 2012. Available from: https://www.unaids.org/en/dataanalysis/knowyourresponse/countryprogressreports/2012countries#E |
| | 2018 | UNAIDS/WHO/UNICEF. Global AIDS Monitoring, 2020. Accessed at: https://aidsinfo.unaids.org/ |
| Gabon | 2018 | UNAIDS/WHO/UNICEF. Global AIDS Monitoring, 2020. Accessed at: https://aidsinfo.unaids.org/ |
| Gambia | 2010 | National AIDS Secretariat Office of the President, Country progress report – The Gambia, 2012. Available from: https://www.unaids.org/en/dataanalysis/knowyourresponse/countryprogressreports/2012countries#G |

| | | |
|---|---|---|
| | 2011 | National AIDS Secretariat Office of the President, Country progress report – The Gambia, 2012. Available from: https://www.unaids.org/en/dataanalysis/knowyourresponse/countryprogressreports/2012countries#G |
| | 2013 | National AIDS Secretariat Office of the President, The Gambia Global AIDS Response Progress Report, 2015. Available from: https://www.unaids.org/en/dataanalysis/knowyourresponse/countryprogressreports/2015countries#G |
| | 2014 | National AIDS Secretariat Office of the President, The Gambia Global AIDS Response Progress Report, 2015. Available from: https://www.unaids.org/en/dataanalysis/knowyourresponse/countryprogressreports/2015countries#G |
| Ghana | 2011 | Ghana AIDS Commission, Country AIDS Response Progress Report – Ghana, 2015. Available from: https://www.unaids.org/en/dataanalysis/knowyourresponse/countryprogressreports/2015countries#G |
| Guinea | 2014 | Comité National de Lutte contre le SIDA, Rapport national de la riposte VIH/SIDA 2014 - Progrès 2010-2014, 2015. Available from: https://www.unaids.org/en/dataanalysis/knowyourresponse/countryprogressreports/2015countries#G |
| | 2018 | UNAIDS/WHO/UNICEF. Global AIDS Monitoring, 2020. Accessed at: https://aidsinfo.unaids.org/ |
| Lesotho | 2005 | Lesotho UNGASS Country Report - Status of the national response to the 2001 declaration of commitment on HIV and AIDS, 2010. Available from: https://www.unaids.org/en/dataanalysis/knowyourresponse/countryprogressreports/2010countries#L |
| | 2006 | Lesotho UNGASS Country Report - Status of the national response to the 2001 declaration of commitment on HIV and AIDS, 2010. Available from: https://www.unaids.org/en/dataanalysis/knowyourresponse/countryprogressreports/2010countries#L |
| | 2007 | Lesotho UNGASS Country Report - Status of the national response to the 2001 declaration of commitment on HIV and AIDS, 2010. Available from: https://www.unaids.org/en/dataanalysis/knowyourresponse/countryprogressreports/2010countries#L |
| | 2008 | Lesotho UNGASS Country Report - Status of the national response to the 2001 declaration of commitment on HIV and AIDS, 2010. Available from: https://www.unaids.org/en/dataanalysis/knowyourresponse/countryprogressreports/2010countries#L |
| | 2009 | Lesotho UNGASS Country Report - Status of the national response to the 2001 declaration of commitment on HIV and AIDS, 2010. Available from: https://www.unaids.org/en/dataanalysis/knowyourresponse/countryprogressreports/2010countries#L |
| Liberia | 2013 | Jacobs GP et al. 2017. Did the 2014 Ebola outbreak in Liberia affect HIV testing, linkage to care and ART initiation?, Public Health Action, 7(S1):S70-S75) |
| Madagascar | 2010 | Comité National de Lutte contre le SIDA, Rapport sur la réponse face au VIH et au SIDA à Madagascar 2014, 2015. Available from: https://www.unaids.org/en/dataanalysis/knowyourresponse/countryprogressreports/2015countries#M |
| | 2011 | Comité National de Lutte contre le SIDA, Rapport sur la réponse face au VIH et au SIDA à Madagascar 2014, 2015. Available from: https://www.unaids.org/en/dataanalysis/knowyourresponse/countryprogressreports/2015countries#M |
| | 2012 | Comité National de Lutte contre le SIDA, Rapport sur la réponse face au VIH et au SIDA à Madagascar 2014, 2015. Available from: https://www.unaids.org/en/dataanalysis/knowyourresponse/countryprogressreports/2015countries#M |
| | 2013 | Comité National de Lutte contre le SIDA, Rapport sur la réponse face au VIH et au SIDA à Madagascar 2014, 2015. Available from: https://www.unaids.org/en/dataanalysis/knowyourresponse/countryprogressreports/2015countries#M |
| | 2014 | Comité National de Lutte contre le SIDA, Rapport sur la réponse face au VIH et au SIDA à Madagascar 2014, 2015. Available from: https://www.unaids.org/en/dataanalysis/knowyourresponse/countryprogressreports/2015countries#M |
| | 2018 | UNAIDS/WHO/UNICEF. Global AIDS Monitoring, 2020. Accessed at: https://aidsinfo.unaids.org/ |
| Mozambique | 2013 | Maheu-Giroux M, Marsh K, Doyle C, et al. National HIV testing and diagnosis coverage in sub-Saharan Africa: a new modeling tool for estimating the “first 90” from program and survey data. *AIDS* 2019; **33**: S255–S69. |
| | 2014 | Maheu-Giroux M, Marsh K, Doyle C, et al. National HIV testing and diagnosis coverage in sub-Saharan Africa: a new modeling tool for estimating the “first 90” from program and survey data. *AIDS* 2019; **33**: S255–S69. |
| | 2016 | Maheu-Giroux M, Marsh K, Doyle C, et al. National HIV testing and diagnosis coverage in sub-Saharan Africa: a new modeling tool for estimating the “first 90” from program and survey data. *AIDS* 2019; **33**: S255–S69. |
| Namibia | 2008 | Republic of Namibia Ministry of Health and Social Services, United Nations General Assembly Special Session (UNGASS) Country Report - Reporting Period 2008-2009, 2010. Available from: https://www.unaids.org/en/dataanalysis/knowyourresponse/countryprogressreports/2010countries#L |
| Niger | 2018 | UNAIDS/WHO/UNICEF. Global AIDS Monitoring, 2020. Accessed at: https://aidsinfo.unaids.org/ |
| Nigeria | 2018 | UNAIDS/WHO/UNICEF. Global AIDS Monitoring, 2020. Accessed at: https://aidsinfo.unaids.org/ |
| Senegal | 2008 | Conseil National de Lutte contre le SIDA, Rapport de situation sur la riposte nationale à l'épidémie de VIH/SIDA – Sénégal: 2008-2009, 2010. Available from: https://www.unaids.org/en/dataanalysis/knowyourresponse/countryprogressreports/2010countries#S |
| | 2012 | Conseil National de Lutte contre le SIDA, Rapport de situation sur la riposte nationale à l'épidémie de VIH/SIDA – Sénégal: 2012-2013, 2014. Availble from: https://www.unaids.org/en/dataanalysis/knowyourresponse/countryprogressreports/2014countries#S |
| | 2013 | Conseil National de Lutte contre le SIDA, Rapport de situation sur la riposte nationale à l’épidémie de VIH/SIDA – Sénégal: 2012-2013, 2014. Available from: https://www.unaids.org/en/dataanalysis/knowyourresponse/countryprogressreports/2014countries#S |

| | | |
|---|---|---|
| Sierra Leone | 2012 | Sierra Leone National AIDS Response Progress Report 2014, 2014. Available from: https://www.unaids.org/en/dataanalysis/knowyourresponse/countryprogressreports/2014countries#S |
| South Africa | 2010 | Johnson L and Dorrington R. 2020. Thembisa version 4.3: A model for evaluating the impact of HIV/AIDS in South Africa. Available at: https://www.google.com/url?sa=t&rct=j&q=&esrc=s&source=web&cd=&ved=2ahUKEwiE9-bQ09frAhXuhXIEHe0vCvcQFjAAegQIBRAB&url=https%3A%2F%2Fwww.thembisa.org%2Fcontent%2Ffiledl%2FThembisa4_3report&usg=AOvVaw1XThc7M7vadKpVmMAXrDKQ |
| | 2011 | Johnson L and Dorrington R. 2020. Thembisa version 4.3: A model for evaluating the impact of HIV/AIDS in South Africa. Available at: https://www.google.com/url?sa=t&rct=j&q=&esrc=s&source=web&cd=&ved=2ahUKEwiE9-bQ09frAhXuhXIEHe0vCvcQFjAAegQIBRAB&url=https%3A%2F%2Fwww.thembisa.org%2Fcontent%2Ffiledl%2FThembisa4_3report&usg=AOvVaw1XThc7M7vadKpVmMAXrDKQ |
| | 2012 | Johnson L and Dorrington R. 2020. Thembisa version 4.3: A model for evaluating the impact of HIV/AIDS in South Africa. Available at: https://www.google.com/url?sa=t&rct=j&q=&esrc=s&source=web&cd=&ved=2ahUKEwiE9-bQ09frAhXuhXIEHe0vCvcQFjAAegQIBRAB&url=https%3A%2F%2Fwww.thembisa.org%2Fcontent%2Ffiledl%2FThembisa4_3report&usg=AOvVaw1XThc7M7vadKpVmMAXrDKQ |
| | 2013 | Johnson L and Dorrington R. 2020. Thembisa version 4.3: A model for evaluating the impact of HIV/AIDS in South Africa. Available at: https://www.google.com/url?sa=t&rct=j&q=&esrc=s&source=web&cd=&ved=2ahUKEwiE9-bQ09frAhXuhXIEHe0vCvcQFjAAegQIBRAB&url=https%3A%2F%2Fwww.thembisa.org%2Fcontent%2Ffiledl%2FThembisa4_3report&usg=AOvVaw1XThc7M7vadKpVmMAXrDKQ |
| | 2014 | Johnson L and Dorrington R. 2020. Thembisa version 4.3: A model for evaluating the impact of HIV/AIDS in South Africa. Available at: https://www.google.com/url?sa=t&rct=j&q=&esrc=s&source=web&cd=&ved=2ahUKEwiE9-bQ09frAhXuhXIEHe0vCvcQFjAAegQIBRAB&url=https%3A%2F%2Fwww.thembisa.org%2Fcontent%2Ffiledl%2FThembisa4_3report&usg=AOvVaw1XThc7M7vadKpVmMAXrDKQ |
| | 2015 | Johnson L and Dorrington R. 2020. Thembisa version 4.3: A model for evaluating the impact of HIV/AIDS in South Africa. Available at: https://www.google.com/url?sa=t&rct=j&q=&esrc=s&source=web&cd=&ved=2ahUKEwiE9-bQ09frAhXuhXIEHe0vCvcQFjAAegQIBRAB&url=https%3A%2F%2Fwww.thembisa.org%2Fcontent%2Ffiledl%2FThembisa4_3report&usg=AOvVaw1XThc7M7vadKpVmMAXrDKQ |
| | 2016 | Johnson L and Dorrington R. 2020. Thembisa version 4.3: A model for evaluating the impact of HIV/AIDS in South Africa. Available at: https://www.google.com/url?sa=t&rct=j&q=&esrc=s&source=web&cd=&ved=2ahUKEwiE9-bQ09frAhXuhXIEHe0vCvcQFjAAegQIBRAB&url=https%3A%2F%2Fwww.thembisa.org%2Fcontent%2Ffiledl%2FThembisa4_3report&usg=AOvVaw1XThc7M7vadKpVmMAXrDKQ |
| | 2017 | Johnson L and Dorrington R. 2020. Thembisa version 4.3: A model for evaluating the impact of HIV/AIDS in South Africa. Available at: https://www.google.com/url?sa=t&rct=j&q=&esrc=s&source=web&cd=&ved=2ahUKEwiE9-bQ09frAhXuhXIEHe0vCvcQFjAAegQIBRAB&url=https%3A%2F%2Fwww.thembisa.org%2Fcontent%2Ffiledl%2FThembisa4_3report&usg=AOvVaw1XThc7M7vadKpVmMAXrDKQ |
| | 2018 | UNAIDS/WHO/UNICEF. Global AIDS Monitoring, 2020. Accessed at: https://aidsinfo.unaids.org/ |
| Togo | 2002 | Conseil National de Lutte contre le SIDA et les Infections Sexuellement Transmissibles, Suivi de la déclaration d'engagement sur le VIH et le SIDA - Rapport 2010, 2010. Available from: https://www.unaids.org/en/dataanalysis/knowyourresponse/countryprogressreports/2010countries#T |
| | 2003 | Conseil National de Lutte contre le SIDA et les Infections Sexuellement Transmissibles, Suivi de la déclaration d'engagement sur le VIH et le SIDA - Rapport 2010, 2010. Available from: https://www.unaids.org/en/dataanalysis/knowyourresponse/countryprogressreports/2010countries#T |
| | 2004 | Conseil National de Lutte contre le SIDA et les Infections Sexuellement Transmissibles, Suivi de la déclaration d'engagement sur le VIH et le SIDA - Rapport 2010, 2010. Available from: https://www.unaids.org/en/dataanalysis/knowyourresponse/countryprogressreports/2010countries#T |
| | 2005 | Conseil National de Lutte contre le SIDA et les Infections Sexuellement Transmissibles, Suivi de la déclaration d'engagement sur le VIH et le SIDA - Rapport 2010, 2010. Available from: https://www.unaids.org/en/dataanalysis/knowyourresponse/countryprogressreports/2010countries#T |
| | 2006 | Conseil National de Lutte contre le SIDA et les Infections Sexuellement Transmissibles, Suivi de la déclaration d'engagement sur le VIH et le SIDA - Rapport 2010, 2010. Available from: https://www.unaids.org/en/dataanalysis/knowyourresponse/countryprogressreports/2010countries#T |
| | 2007 | Conseil National de Lutte contre le SIDA et les Infections Sexuellement Transmissibles, Suivi de la déclaration d'engagement sur le VIH et le SIDA - Rapport 2010, 2010. Available from: https://www.unaids.org/en/dataanalysis/knowyourresponse/countryprogressreports/2010countries#T |
| | 2008 | Conseil National de Lutte contre le SIDA et les Infections Sexuellement Transmissibles, Suivi de la déclaration d'engagement sur le VIH et le SIDA - Rapport 2010, 2010. Available from: https://www.unaids.org/en/dataanalysis/knowyourresponse/countryprogressreports/2010countries#T |

| | | |
|---|---|---|
| | 2009 | Conseil National de Lutte contre le SIDA et les Infections Sexuellement Transmissibles, Suivi de la déclaration d'engagement sur le VIH et le SIDA - Rapport 2010, 2010. Available from: https://www.unaids.org/en/dataanalysis/knowyourresponse/countryprogressreports/2010countries#T |
| | 2012 | Conseil National de Lutte contre le SIDA et les Infections Sexuellement Transmissibles, Rapport annuel des activités de lutte contre le VIH/SIDA au Togo en 2012, 2013. |
| | 2014 | Conseil National de Lutte contre le SIDA et les Infections Sexuellement Transmissibles, Rapport d'activité sur la riposte au VIH/SIDA au Togo, 2015. Available from: https://www.unaids.org/en/dataanalysis/knowyourresponse/countryprogressreports/2015countries#T |
| | 2015 | Programme national de lutte contre le SIDA et les IST, Rapport annuel 2015 des activités du PNLS-IST, 2015. Available from: https://www.google.com/url?sa=t&rct=j&q=&esrc=s&source=web&cd=&ved=2ahUKEwiY3LSw2dfrAhWNmlkKHdy_AgQQFjAAegQIAxAB&url=http%3A%2F%2Fwww.pnls.tg%2Frapports%2FRAPPORT%2520ANNUEL%2520PNLS%25202015.pdf&usg=AOvVaw2rlrRl38FNKwcgBhBHxI_u |
| Zambia | 2006 | National AIDS Council, Zambia country report - Monitoring the declaration of commitment on HIV and AIDS and the universal access, 2010. Available from: https://www.unaids.org/en/dataanalysis/knowyourresponse/countryprogressreports/2010countries#T |
| | 2007 | National AIDS Council, Zambia country report - Monitoring the declaration of commitment on HIV and AIDS and the universal access, 2010. Available from: https://www.unaids.org/en/dataanalysis/knowyourresponse/countryprogressreports/2010countries#T |
| | 2008 | National AIDS Council, Zambia country report - Monitoring the declaration of commitment on HIV and AIDS and the universal access, 2010. Available from: https://www.unaids.org/en/dataanalysis/knowyourresponse/countryprogressreports/2010countries#T |
| | 2009 | National AIDS Council, Zambia country report - Monitoring the declaration of commitment on HIV and AIDS and the universal access, 2010. Available from: https://www.unaids.org/en/dataanalysis/knowyourresponse/countryprogressreports/2010countries#T |

***Table S2*: Checklist of information that should be included in new reports of global health estimates**

| Item # | Checklist item | Reported on page # |
|---|---|---|
| **Objectives and funding** | | |
| **1** | Define the indicator(s), populations (including age, sex, and geographic entities), and time period(s) for which estimates were made. | 7, 8 |
| **2** | List the funding sources for the work. | 2 |
| **Data Inputs** | | |
| *For all data inputs from multiple sources that are synthesized as part of the study:* | | |
| **3** | Describe how the data were identified and how the data were accessed. | 6 |
| **4** | Specify the inclusion and exclusion criteria. Identify all ad-hoc exclusions. | 6, 7 |
| **5** | Provide information on all included data sources and their main characteristics. For each data source used, report reference information or contact name/institution, population represented, data collection method, year(s) of data collection, sex and age range, diagnostic criteria or measurement method, and sample size, as relevant. | 6, Figure 1, appendix pp 47-50 |
| **6** | Identify and describe any categories of input data that have potentially important biases (e.g., based on characteristics listed in item 5). | 6, 12, 13 |
| *For data inputs that contribute to the analysis but were not synthesized as part of the study:* | | |
| **7** | Describe and give sources for any other data inputs. | 6, Figure 1, appendix pp 47-50 |
| *For all data inputs:* | | |
| **8** | Provide all data inputs in a file format from which data can be efficiently extracted (e.g., a spreadsheet rather than a PDF), including all relevant meta-data listed in item 5. For any data inputs that cannot be shared because of ethical or legal reasons, such as third-party ownership, provide a contact name or the name of the institution that retains the right to the data. | 6 (links provided) |
| **Data analysis** | | |
| **9** | Provide a conceptual overview of the data analysis method. A diagram may be helpful. | 5 |
| **10** | Provide a detailed description of all steps of the analysis, including mathematical formulae. This description should cover, as relevant, data cleaning, data pre-processing, data adjustments and weighting of data sources, and mathematical or statistical model(s). | 5-8, appendix p 1 |
| **11** | Describe how candidate models were evaluated and how the final model(s) were selected. | 5 |
| **12** | Provide the results of an evaluation of model performance, if done, as well as the results of any relevant sensitivity analysis. | appendix pp 2-42 |
| **13** | Describe methods for calculating uncertainty of the estimates. State which sources of uncertainty were, and were not, accounted for in the uncertainty analysis. | 8, 12 |
| **14** | State how analytic or statistical source code used to generate estimates can be accessed. | 8 |
| **Results and Discussion** | | |
| **15** | Provide published estimates in a file format from which data can be efficiently extracted. | Table 1, appendix pp 52-57 |
| **16** | Report a quantitative measure of the uncertainty of the estimates (e.g. uncertainty intervals). | 2, 9-11, Tables |
| **17** | Interpret results in light of existing evidence. If updating a previous set of estimates, describe the reasons for changes in estimates. | 10-13 |
| **18** | Discuss limitations of the estimates. Include a discussion of any modelling assumptions or data limitations that affect interpretation of the estimates. | 12-13 |

***Table S3*: Number of undiagnosed people living with HIV by sex and age stratification and by sub-Saharan African region in 2020 (x 1,000)**

| Sex | Age group (years) | Sub-Saharan Africa | | Western Africa | | Central Africa | | Eastern Africa | | Southern Africa | |
|---|---|---|---|---|---|---|---|---|---|---|---|
| Men | 15-24 | 346 | (318 to 366) | 89 | (85 to 91) | 33 | (30 to 35) | 163 | (150 to 172) | 61 | (51 to 69) |
| | 25-34 | 597 | (514 to 673) | 156 | (143 to 169) | 53 | (40 to 62) | 258 | (227 to 287) | 130 | (100 to 156) |
| | 35-49 | 701 | (611 to 788) | 211 | (191 to 230) | 57 | (45 to 68) | 296 | (265 to 327) | 136 | (104 to 169) |
| | 50-99 | 264 | (231 to 299) | 91 | (81 to 97) | 18 | (14 to 21) | 109 | (96 to 125) | 46 | (36 to 58) |
| Women | 15-24 | 563 | (492 to 621) | 124 | (119 to 129) | 79 | (63 to 91) | 222 | (200 to 240) | 138 | (108 to 164) |
| | 25-34 | 557 | (459 to 652) | 173 | (154 to 188) | 92 | (66 to 123) | 196 | (170 to 224) | 94 | (65 to 125) |
| | 35-49 | 581 | (507 to 664) | 166 | (156 to 179) | 100 | (78 to 123) | 199 | (179 to 221) | 115 | (87 to 145) |
| | 50-99 | 234 | (202 to 272) | 54 | (51 to 59) | 39 | (31 to 46) | 77 | (71 to 87) | 64 | (48 to 83) |

Numbers in parentheses correspond to 95% credible intervals.
Some small discrepancies are observed between sub-Saharan Africa estimate and the sum of Western, Central, Eastern, and Southern regions estimates because each region-specific estimate is based on independent draws of posterior estimates.

***Table S4*: Progress in HIV testing related outcomes in sub-Saharan Africa stratified by sex and age group, 2000-2020**

| Outcome | Year | Overall | Men | Women | 15-24 years-old | 25-34 years-old | 35-49 years-old | 50-99 years-old |
|---|---|---|---|---|---|---|---|---|
| Proportion of individuals having ever been tested for HIV among overall population (%) | 2000 | 3·6 (3·0 to 4·4) | 3·3 (2·6 to 4·0) | 4·0 (3·3 to 4·9) | 3·2 (2·6 to 3·9) | 5·1 (4·2 to 6·2) | 3·5 (2·9 to 4·3) | 2·6 (2·2 to 3·3) |
| | 2005 | 11 (10 to 12) | 9·9 (9·0 to 11) | 13 (12 to 14) | 8·8 (7·9 to 9·9) | 15 (14 to 17) | 12 (11 to 13) | 9·1 (8·4 to 10) |
| | 2010 | 30 (29 to 32) | 27 (26 to 29) | 33 (32 to 35) | 25 (23 to 26) | 39 (37 to 41) | 33 (31 to 34) | 26 (25 to 28) |
| | 2015 | 41 (40 to 42) | 37 (36 to 38) | 46 (45 to 47) | 29 (28 to 31) | 52 (51 to 53) | 49 (48 to 50) | 40 (38 to 42) |
| | 2020 | 51 (49 to 54) | 46 (43 to 49) | 56 (54 to 60) | 35 (32 to 39) | 61 (59 to 64) | 62 (61 to 65) | 54 (51 to 57) |
| Proportion of PLHIV who know their HIV status (%) | 2000 | 5·7 (4·6 to 7·0) | 4·6 (3·7 to 5·8) | 6·4 (5·1 to 7·9) | 4·9 (3·8 to 6·1) | 7·2 (5·7 to 8·8) | 4·9 (4·0 to 6·1) | 3·5 (2·8 to 4·3) |
| | 2005 | 20 (18 to 22) | 16 (14 to 18) | 22 (20 to 25) | 14 (12 to 16) | 24 (21 to 27) | 19 (17 to 22) | 15 (14 to 17) |
| | 2010 | 53 (50 to 55) | 46 (43 to 48) | 57 (54 to 59) | 38 (35 to 41) | 58 (54 to 60) | 55 (52 to 57) | 50 (48 to 53) |
| | 2015 | 71 (69 to 73) | 64 (62 to 66) | 75 (73 to 77) | 51 (49 to 53) | 72 (69 to 74) | 76 (74 to 78) | 75 (73 to 77) |
| | 2020 | 84 (82 to 86) | 79 (76 to 81) | 87 (85 to 89) | 65 (62 to 69) | 81 (79 to 84) | 88 (86 to 89) | 90 (88 to 91) |
| Time to diagnosis or AIDS-related death, median (year) | 2000 | 9·6 (9·1 to 10) | 9·7 (9·2 to 10) | 9·5 (8·9 to 10) | 11 (10 to 12) | 9·1 (8·5 to 9·5) | 8·2 (7·9 to 8·5) | 7·6 (7·5 to 7·8) |
| | 2005 | 7·2 (6·3 to 8·0) | 7·7 (6·8 to 8·4) | 6·9 (5·9 to 7·8) | 7·7 (6·6 to 8·7) | 6·8 (5·9 to 7·6) | 6·8 (6·3 to 7·3) | 6·6 (6·2 to 6·9) |
| | 2010 | 3·6 (3·2 to 4·1) | 4·3 (3·8 to 4·7) | 3·2 (2·8 to 3·7) | 3·7 (3·2 to 4·3) | 3·1 (2·7 to 3·6) | 4·0 (3·6 to 4·4) | 4·2 (3·9 to 4·6) |
| | 2015 | 3·0 (2·7 to 3·4) | 3·7 (3·4 to 4·2) | 2·6 (2·2 to 2·9) | 3·2 (2·8 to 3·6) | 2·5 (2·2 to 2·9) | 3·3 (3·0 to 3·7) | 3·5 (3·3 to 3·8) |
| | 2020 | 2·6 (1·8 to 3·5) | 3·3 (2·4 to 4·2) | 2·2 (1·5 to 3·0) | 2·8 (2·0 to 3·8) | 2·2 (1·5 to 3·0) | 2·8 (2·0 to 3·6) | 2·9 (2·2 to 3·6) |
| Probability of getting tested within 1 year following infection (%) | 2000 | 2·6 (2·1 to 3·3) | 2·1 (1·6 to 2·6) | 3·0 (2·3 to 3·8) | 2·6 (2·0 to 3·3) | 3·2 (2·5 to 4·0) | 2·1 (1·6 to 2·6) | 1·4 (1·1 to 1·8) |
| | 2005 | 5·9 (4·5 to 7·7) | 4·6 (3·5 to 6·1) | 6·7 (5·0 to 8·8) | 5·9 (4·5 to 7·8) | 6·9 (5·3 to 9·1) | 4·5 (3·4 to 5·9) | 3·5 (2·7 to 4·6) |
| | 2010 | 21 (18 to 25) | 16 (13 to 19) | 25 (21 to 29) | 21 (18 to 25) | 26 (22 to 31) | 16 (13 to 19) | 12 (9·9 to 15) |
| | 2015 | 26 (23 to 30) | 19 (16 to 22) | 31 (27 to 35) | 26 (23 to 29) | 32 (28 to 37) | 20 (17 to 23) | 16 (14 to 19) |
| | 2020 | 33 (23 to 46) | 25 (17 to 36) | 39 (27 to 53) | 32 (23 to 45) | 40 (28 to 54) | 27 (17 to 39) | 23 (15 to 33) |
| Probability of getting tested before reaching a CD4 count lower than 350 cells per µL (%) | 2000 | 19 (16 to 23) | 16 (13 to 19) | 22 (18 to 26) | 24 (20 to 28) | 20 (16 to 24) | 12 (10 to 15) | 7·2 (5·8 to 8·9) |
| | 2005 | 35 (29 to 42) | 30 (24 to 36) | 38 (32 to 45) | 42 (35 to 49) | 36 (30 to 43) | 24 (19 to 29) | 16 (13 to 20) |
| | 2010 | 63 (58 to 66) | 56 (51 to 60) | 67 (63 to 71) | 68 (64 to 72) | 67 (63 to 71) | 50 (45 to 55) | 38 (33 to 42) |
| | 2015 | 67 (63 to 70) | 60 (55 to 63) | 72 (68 to 75) | 71 (67 to 75) | 72 (68 to 76) | 56 (51 to 60) | 44 (40 to 48) |
| | 2020 | 71 (62 to 79) | 64 (54 to 73) | 76 (67 to 83) | 74 (66 to 82) | 76 (67 to 83) | 62 (51 to 71) | 52 (41 to 62) |
| Positivity (% of positive test among all tests) | 2000 | 9·0 (7·7 to 10) | 6·6 (5·7 to 7·5) | 11 (9·3 to 13) | 5·8 (5·1 to 6·6) | 13 (11 to 15) | 10 (9·0 to 12) | 3·4 (2·9 to 3·9) |
| | 2005 | 11 (9·2 to 14) | 7·9 (6·9 to 9·3) | 13 (11 to 17) | 5·6 (4·8 to 6·8) | 16 (13 to 21) | 15 (13 to 18) | 5·4 (4·8 to 6·2) |
| | 2010 | 5·9 (4·3 to 8·3) | 4·3 (3·3 to 5·8) | 6·9 (5·0 to 10) | 3·1 (2·4 to 4·2) | 7·5 (5·2 to 11) | 9·5 (7·3 to 13) | 3·8 (3·0 to 4·8) |
| | 2015 | 4·3 (3·5 to 5·2) | 3·6 (3·0 to 4·3) | 4·6 (3·8 to 5·8) | 2·5 (2·1 to 2·9) | 4·8 (3·7 to 6·1) | 6·9 (5·7 to 8·5) | 3·4 (2·7 to 4·2) |
| | 2020 | 2·8 (2·1 to 3·9) | 2·6 (1·9 to 3·4) | 3·0 (2·2 to 4·3) | 1·6 (1·3 to 2·0) | 3·0 (2·2 to 4·1) | 4·6 (3·3 to 6·7) | 2·8 (1·9 to 4·3) |
| Diagnosis yield (% of new diagnoses among all tests) | 2000 | 7·9 (7·0 to 8·6) | 6·0 (5·2 to 6·6) | 9·4 (8·3 to 10) | 5·3 (4·7 to 5·8) | 11 (10 to 12) | 9·4 (8·2 to 10) | 3·1 (2·7 to 3·5) |
| | 2005 | 7·8 (7·0 to 8·4) | 6·2 (5·6 to 6·7) | 8·9 (8·0 to 9·7) | 4·4 (4·0 to 4·8) | 11 (9·4 to 12) | 11 (10 to 12) | 4·5 (4·0 to 5·0) |
| | 2010 | 2·8 (2·4 to 3·3) | 2·5 (2·2 to 3·0) | 3·0 (2·6 to 3·6) | 1·8 (1·6 to 2·1) | 3·0 (2·5 to 3·5) | 4·9 (4·2 to 5·7) | 2·5 (2·1 to 2·9) |
| | 2015 | 1·9 (1·7 to 2·1) | 1·9 (1·6 to 2·1) | 1·9 (1·7 to 2·1) | 1·5 (1·3 to 1·6) | 1·8 (1·6 to 2·0) | 2·9 (2·5 to 3·3) | 1·8 (1·6 to 2·1) |
| | 2020 | 1·2 (0·9 to 1·5) | 1·1 (0·9 to 1·5) | 1·2 (0·9 to 1·6) | 1·0 (0·8 to 1·2) | 1·1 (0·8 to 1·4) | 1·7 (1·2 to 2·3) | 1·2 (0·8 to 1·7) |
| Proportion of new HIV diagnoses among all positive tests (%) | 2000 | 89 (77 to 96) | 91 (83 to 97) | 88 (74 to 96) | 92 (82 to 97) | 87 (73 to 95) | 91 (80 to 97) | 92 (83 to 97) |
| | 2005 | 72 (55 to 86) | 79 (65 to 89) | 69 (51 to 84) | 80 (65 to 91) | 66 (48 to 82) | 76 (60 to 88) | 84 (72 to 92) |
| | 2010 | 48 (33 to 65) | 59 (44 to 73) | 44 (29 to 62) | 59 (43 to 75) | 39 (25 to 58) | 52 (38 to 67) | 66 (51 to 79) |
| | 2015 | 44 (37 to 53) | 52 (44 to 60) | 41 (33 to 50) | 60 (52 to 68) | 37 (29 to 47) | 43 (34 to 51) | 55 (43 to 65) |
| | 2020 | 42 (30 to 55) | 45 (34 to 56) | 40 (28 to 55) | 61 (50 to 73) | 36 (25 to 49) | 37 (25 to 50) | 42 (28 to 59) |

PLHIV: people living with HIV, HTS: HIV testing services. Numbers in parentheses correspond to 95% credible intervals.

*Table S5*: **Progress in HIV testing related outcomes in Western Africa stratified by sex and age group, 2000-2020**

| Outcome | Year | Overall | Men | Women | 15-24 years-old | 25-34 years-old | 35-49 years-old | 50-99 years-old |
|---|---|---|---|---|---|---|---|---|
| Proportion of individuals having ever been tested for HIV among overall population (%) | 2000 | 3·1 (2·6 to 3·8) | 3·1 (2·5 to 3·7) | 3·2 (2·6 to 3·8) | 2·7 (2·2 to 3·2) | 4·7 (3·9 to 5·8) | 2·8 (2·3 to 3·4) | 2·1 (1·7 to 2·6) |
| | 2005 | 7·2 (6·6 to 7·8) | 6·9 (6·2 to 7·6) | 7·4 (6·8 to 8·1) | 5·0 (4·4 to 5·6) | 11 (9·7 to 12) | 8·1 (7·5 to 8·7) | 5·5 (5·0 to 6·0) |
| | 2010 | 19 (18 to 19) | 17 (16 to 18) | 20 (19 to 21) | 13 (12 to 14) | 26 (25 to 27) | 21 (20 to 22) | 15 (14 to 16) |
| | 2015 | 28 (27 to 29) | 24 (23 to 25) | 33 (31 to 34) | 18 (17 to 20) | 37 (36 to 39) | 35 (34 to 36) | 26 (24 to 27) |
| | 2020 | 36 (34 to 39) | 30 (28 to 32) | 42 (40 to 45) | 21 (18 to 24) | 45 (43 to 48) | 47 (46 to 49) | 37 (35 to 40) |
| Proportion of PLHIV who know their HIV status (%) | 2000 | 4·0 (2·8 to 5·2) | 3·9 (2·7 to 5·0) | 4·2 (2·9 to 5·4) | 3·0 (2·0 to 3·8) | 5·4 (3·7 to 6·9) | 3·5 (2·5 to 4·5) | 2·7 (1·9 to 3·5) |
| | 2005 | 10 (7·7 to 12) | 9·2 (6·8 to 11) | 11 (8·3 to 13) | 5·4 (4·0 to 6·7) | 12 (8·5 to 14) | 11 (8·2 to 12) | 9·7 (8·1 to 11) |
| | 2010 | 33 (28 to 35) | 28 (24 to 31) | 36 (31 to 39) | 19 (16 to 22) | 35 (28 to 38) | 34 (31 to 37) | 37 (34 to 38) |
| | 2015 | 52 (48 to 55) | 43 (39 to 46) | 58 (54 to 60) | 32 (29 to 34) | 48 (42 to 52) | 57 (54 to 59) | 61 (59 to 63) |
| | 2020 | 67 (65 to 69) | 55 (52 to 59) | 74 (72 to 76) | 42 (40 to 44) | 58 (55 to 62) | 72 (70 to 74) | 79 (77 to 81) |
| Time to diagnosis or AIDS-related death, median (year) | 2000 | 11 (10 to 11) | 10 (9·7 to 11) | 11 (10 to 11) | 12 (12 to 13) | 10 (9·5 to 11) | 9·0 (8·8 to 9·3) | 7·9 (7·8 to 8·1) |
| | 2005 | 10 (9·4 to 11) | 10 (9·4 to 11) | 10 (9·5 to 11) | 12 (11 to 13) | 9·9 (9·1 to 11) | 8·8 (8·5 to 9·1) | 7·8 (7·5 to 7·9) |
| | 2010 | 5·5 (4·9 to 6·5) | 6·1 (5·4 to 7·0) | 5·1 (4·5 to 6·1) | 6·2 (5·4 to 7·4) | 4·6 (3·9 to 5·7) | 5·8 (5·3 to 6·4) | 5·7 (5·3 to 6·1) |
| | 2015 | 5·2 (4·5 to 6·0) | 6·0 (5·2 to 6·8) | 4·6 (3·9 to 5·4) | 6·0 (5·2 to 7·0) | 4·3 (3·6 to 5·3) | 5·3 (4·7 to 5·9) | 5·2 (4·8 to 5·6) |
| | 2020 | 5·4 (4·1 to 6·5) | 6·2 (4·8 to 7·4) | 4·8 (3·6 to 6·0) | 6·4 (4·9 to 7·8) | 4·5 (3·3 to 5·8) | 5·1 (4·1 to 6·1) | 5·1 (4·3 to 5·7) |
| Probability of getting tested within 1 year following infection (%) | 2000 | 1·6 (1·1 to 2·1) | 1·5 (1·0 to 2·0) | 1·7 (1·2 to 2·2) | 1·5 (1·0 to 2·0) | 2·2 (1·5 to 2·9) | 1·2 (0·8 to 1·5) | 1·0 (0·7 to 1·3) |
| | 2005 | 1·8 (1·1 to 2·6) | 1·6 (1·0 to 2·3) | 1·9 (1·2 to 2·8) | 1·7 (1·1 to 2·4) | 2·3 (1·5 to 3·3) | 1·3 (0·8 to 1·9) | 1·1 (0·7 to 1·6) |
| | 2010 | 11 (7·7 to 14) | 9·0 (6·2 to 12) | 13 (8·8 to 17) | 9·4 (6·3 to 12) | 16 (11 to 21) | 8·1 (5·6 to 11) | 6·2 (4·4 to 8·0) |
| | 2015 | 13 (9·3 to 17) | 9·8 (6·9 to 13) | 16 (11 to 20) | 10 (7·3 to 14) | 19 (13 to 25) | 10 (7·3 to 13) | 7·8 (5·8 to 10) |
| | 2020 | 12 (7·0 to 19) | 8·4 (4·8 to 14) | 15 (8·7 to 23) | 9·1 (5·3 to 15) | 17 (10 to 28) | 9·8 (5·7 to 16) | 7·8 (4·8 to 13) |
| Probability of getting tested before reaching a CD4 count lower than 350 cells per µL (%) | 2000 | 13 (9·3 to 16) | 12 (8·7 to 15) | 14 (9·6 to 17) | 17 (12 to 21) | 15 (10 to 18) | 7·6 (5·4 to 9·5) | 5·4 (3·8 to 6·8) |
| | 2005 | 14 (9·2 to 19) | 13 (8·3 to 17) | 15 (9·9 to 20) | 18 (12 to 24) | 15 (10 to 21) | 8·3 (5·5 to 12) | 6·0 (3·9 to 8·5) |
| | 2010 | 45 (36 to 51) | 40 (31 to 46) | 49 (40 to 55) | 50 (41 to 56) | 52 (43 to 59) | 33 (25 to 38) | 23 (18 to 28) |
| | 2015 | 47 (38 to 53) | 40 (32 to 47) | 52 (43 to 59) | 50 (41 to 58) | 55 (45 to 62) | 36 (28 to 42) | 26 (20 to 31) |
| | 2020 | 44 (32 to 56) | 37 (25 to 49) | 49 (37 to 61) | 45 (33 to 58) | 52 (39 to 65) | 35 (24 to 47) | 25 (16 to 35) |
| Positivity (% of positive test among all tests) | 2000 | 3·0 (2·1 to 3·5) | 2·5 (1·7 to 2·9) | 3·5 (2·4 to 4·1) | 1·4 (1·0 to 1·6) | 4·2 (2·9 to 5·0) | 4·1 (2·9 to 4·8) | 1·8 (1·3 to 2·1) |
| | 2005 | 4·0 (3·3 to 4·5) | 2·9 (2·4 to 3·4) | 4·9 (4·1 to 5·5) | 1·3 (1·0 to 1·6) | 4·8 (3·7 to 5·6) | 7·0 (6·1 to 7·9) | 4·5 (3·8 to 5·4) |
| | 2010 | 2·6 (2·0 to 3·2) | 2·0 (1·6 to 2·5) | 3·0 (2·3 to 3·8) | 1·1 (0·9 to 1·3) | 2·8 (2·0 to 3·7) | 4·6 (3·7 to 5·6) | 2·7 (2·2 to 3·3) |
| | 2015 | 2·2 (1·7 to 2·7) | 1·9 (1·4 to 2·3) | 2·4 (1·8 to 3·0) | 1·0 (0·8 to 1·2) | 2·1 (1·5 to 2·6) | 4·0 (3·1 to 4·9) | 2·7 (2·1 to 3·4) |
| | 2020 | 1·9 (1·3 to 2·7) | 1·7 (1·3 to 2·4) | 2·0 (1·4 to 2·9) | 0·9 (0·7 to 1·3) | 1·6 (1·2 to 2·2) | 3·2 (2·3 to 4·6) | 2·7 (1·8 to 4·0) |
| Diagnosis yield (% of new diagnoses among all tests) | 2000 | 2·6 (1·8 to 2·9) | 2·1 (1·5 to 2·5) | 2·9 (2·1 to 3·4) | 1·2 (0·9 to 1·4) | 3·5 (2·4 to 4·0) | 3·6 (2·6 to 4·2) | 1·7 (1·2 to 1·9) |
| | 2005 | 3·2 (2·7 to 3·6) | 2·3 (1·9 to 2·6) | 3·9 (3·3 to 4·4) | 1·1 (0·9 to 1·3) | 3·5 (2·7 to 4·0) | 5·6 (4·9 to 6·6) | 4·0 (3·4 to 5·0) |
| | 2010 | 1·5 (1·2 to 1·7) | 1·3 (1·1 to 1·5) | 1·6 (1·3 to 1·9) | 0·7 (0·6 to 0·8) | 1·3 (1·1 to 1·6) | 2·8 (2·4 to 3·3) | 1·9 (1·5 to 2·3) |
| | 2015 | 1·1 (0·9 to 1·3) | 1·0 (0·9 to 1·2) | 1·1 (0·9 to 1·3) | 0·6 (0·5 to 0·7) | 0·9 (0·7 to 1·0) | 1·9 (1·6 to 2·4) | 1·4 (1·1 to 1·8) |
| | 2020 | 1·0 (0·7 to 1·5) | 1·0 (0·7 to 1·4) | 1·0 (0·6 to 1·6) | 0·6 (0·4 to 1·0) | 0·7 (0·5 to 1·0) | 1·7 (1·1 to 2·6) | 1·5 (0·9 to 2·4) |
| Proportion of new HIV diagnoses among all positive tests (%) | 2000 | 86 (79 to 94) | 87 (80 to 95) | 85 (78 to 94) | 91 (86 to 96) | 83 (74 to 93) | 88 (83 to 95) | 91 (87 to 96) |
| | 2005 | 79 (71 to 90) | 80 (72 to 91) | 79 (71 to 89) | 86 (79 to 93) | 74 (65 to 87) | 81 (73 to 90) | 89 (84 to 94) |
| | 2010 | 56 (46 to 73) | 65 (53 to 81) | 53 (43 to 69) | 67 (59 to 81) | 47 (38 to 67) | 61 (49 to 75) | 70 (57 to 82) |
| | 2015 | 48 (38 to 61) | 54 (44 to 68) | 45 (35 to 58) | 60 (51 to 71) | 42 (32 to 57) | 49 (38 to 62) | 53 (41 to 65) |
| | 2020 | 52 (38 to 68) | 56 (44 to 70) | 50 (35 to 68) | 68 (55 to 81) | 44 (31 to 60) | 53 (38 to 69) | 55 (39 to 72) |

PLHIV: people living with HIV, HTS: HIV testing services. Numbers in parentheses correspond to 95% credible intervals.

***Table S6*: Progress in HIV testing related outcomes in Central Africa stratified by sex and age group, 2000-2020**

| Outcome | Year | Overall | Men | Women | 15-24 years-old | 25-34 years-old | 35-49 years-old | 50-99 years-old |
|---|---|---|---|---|---|---|---|---|
| Proportion of individuals having ever been tested for HIV among overall population (%) | 2000 | 1·9 (1·3 to 2·6) | 1·7 (1·2 to 2·3) | 2·0 (1·4 to 2·8) | 1·6 (1·1 to 2·2) | 2·5 (1·8 to 3·5) | 1·9 (1·3 to 2·5) | 1·5 (1·0 to 2·0) |
| | 2005 | 7·8 (6·6 to 9·2) | 7·0 (5·8 to 8·4) | 8·6 (7·3 to 10) | 6·0 (5·0 to 7·3) | 11 (9·0 to 13) | 8·5 (7·3 to 10) | 6·6 (5·6 to 7·8) |
| | 2010 | 19 (17 to 22) | 17 (15 to 20) | 21 (19 to 23) | 13 (12 to 16) | 26 (23 to 29) | 23 (21 to 25) | 18 (16 to 20) |
| | 2015 | 29 (28 to 30) | 26 (24 to 27) | 32 (31 to 33) | 18 (17 to 19) | 37 (35 to 39) | 37 (35 to 38) | 29 (26 to 32) |
| | 2020 | 42 (39 to 48) | 39 (34 to 44) | 47 (42 to 53) | 26 (22 to 33) | 52 (47 to 59) | 54 (50 to 59) | 46 (41 to 51) |
| Proportion of PLHIV who know their HIV status (%) | 2000 | 3·2 (2·0 to 4·6) | 3·1 (2·0 to 4·5) | 3·3 (2·1 to 4·8) | 2·7 (1·7 to 4·0) | 3·9 (2·5 to 5·6) | 2·9 (1·8 to 4·2) | 2·4 (1·5 to 3·4) |
| | 2005 | 15 (12 to 18) | 15 (12 to 18) | 16 (12 to 18) | 11 (8·8 to 14) | 18 (14 to 22) | 15 (12 to 17) | 13 (10 to 15) |
| | 2010 | 37 (32 to 41) | 36 (32 to 40) | 38 (32 to 42) | 21 (17 to 25) | 40 (34 to 45) | 41 (36 to 44) | 38 (35 to 42) |
| | 2015 | 53 (47 to 56) | 50 (45 to 54) | 54 (48 to 58) | 30 (25 to 33) | 53 (45 to 57) | 59 (54 to 63) | 59 (55 to 63) |
| | 2020 | 70 (64 to 76) | 68 (63 to 74) | 71 (65 to 78) | 45 (38 to 54) | 67 (58 to 76) | 76 (71 to 81) | 79 (76 to 83) |
| Time to diagnosis or AIDS-related death, median (year) | 2000 | 11 (10 to 12) | 11 (10 to 11) | 11 (10 to 12) | 13 (12 to 14) | 10 (9·7 to 11) | 9·0 (8·7 to 9·3) | 7·9 (7·7 to 8·1) |
| | 2005 | 8·2 (7·1 to 9·3) | 8·2 (7·0 to 9·2) | 8·3 (7·1 to 9·4) | 9·3 (7·9 to 11) | 7·6 (6·5 to 8·7) | 7·4 (6·7 to 8·0) | 6·7 (6·2 to 7·2) |
| | 2010 | 6·1 (5·2 to 7·3) | 6·1 (5·2 to 7·2) | 6·0 (5·1 to 7·3) | 6·8 (5·9 to 8·3) | 5·3 (4·4 to 6·6) | 5·8 (5·0 to 6·6) | 5·6 (5·0 to 6·2) |
| | 2015 | 4·7 (3·9 to 6·0) | 5·2 (4·4 to 6·5) | 4·5 (3·7 to 5·8) | 5·2 (4·3 to 6·6) | 4·0 (3·1 to 5·4) | 4·8 (4·0 to 5·9) | 4·9 (4·2 to 5·6) |
| | 2020 | 3·9 (2·2 to 6·0) | 4·6 (2·8 to 6·5) | 3·6 (1·9 to 5·7) | 4·3 (2·4 to 6·6) | 3·2 (1·7 to 5·5) | 4·0 (2·4 to 5·8) | 4·1 (2·6 to 5·4) |
| Probability of getting tested within 1 year following infection (%) | 2000 | 1·5 (1·0 to 2·1) | 1·3 (0·9 to 1·9) | 1·5 (1·0 to 2·2) | 1·5 (0·9 to 2·1) | 1·8 (1·2 to 2·6) | 1·2 (0·8 to 1·7) | 1·0 (0·7 to 1·5) |
| | 2005 | 5·1 (3·6 to 7·1) | 4·6 (3·2 to 6·6) | 5·3 (3·7 to 7·4) | 4·9 (3·5 to 6·8) | 6·2 (4·3 to 8·8) | 4·1 (2·8 to 5·7) | 3·6 (2·5 to 4·9) |
| | 2010 | 9·3 (6·5 to 13) | 8·1 (5·7 to 12) | 9·9 (6·8 to 14) | 8·5 (5·9 to 12) | 12 (8·1 to 18) | 7·5 (5·1 to 11) | 6·2 (4·3 to 8·8) |
| | 2015 | 12 (7·8 to 18) | 9·7 (6·3 to 14) | 13 (8·4 to 20) | 11 (6·9 to 16) | 16 (9·9 to 24) | 9·7 (6·2 to 15) | 8·0 (5·3 to 13) |
| | 2020 | 16 (7·6 to 34) | 13 (6·4 to 26) | 17 (8·3 to 37) | 14 (7·0 to 30) | 20 (9·7 to 42) | 13 (6·3 to 30) | 11 (5·6 to 25) |
| Probability of getting tested before reaching a CD4 count lower than 350 cells per µL (%) | 2000 | 12 (8·3 to 17) | 11 (7·8 to 16) | 12 (8·5 to 17) | 15 (10 to 20) | 13 (8·6 to 17) | 7·7 (5·3 to 11) | 5·6 (3·8 to 7·7) |
| | 2005 | 30 (22 to 38) | 29 (22 to 37) | 30 (23 to 38) | 34 (26 to 42) | 32 (24 to 41) | 21 (15 to 27) | 16 (12 to 21) |
| | 2010 | 43 (34 to 51) | 41 (32 to 49) | 44 (34 to 52) | 47 (37 to 54) | 48 (37 to 56) | 33 (25 to 41) | 24 (18 to 31) |
| | 2015 | 52 (41 to 60) | 47 (36 to 56) | 54 (43 to 62) | 56 (45 to 63) | 57 (45 to 66) | 40 (29 to 49) | 30 (21 to 38) |
| | 2020 | 59 (39 to 75) | 52 (34 to 70) | 62 (41 to 78) | 63 (43 to 78) | 64 (42 to 81) | 48 (28 to 67) | 37 (21 to 56) |
| Positivity (% of positive test among all tests) | 2000 | 4·6 (3·5 to 5·2) | 3·1 (2·3 to 3·6) | 5·8 (4·4 to 6·5) | 3·1 (2·4 to 3·6) | 6·6 (5·0 to 7·6) | 5·1 (3·8 to 5·9) | 2·1 (1·6 to 2·4) |
| | 2005 | 5·4 (4·2 to 6·7) | 3·7 (2·9 to 4·5) | 6·7 (5·2 to 8·5) | 2·9 (2·3 to 3·7) | 7·8 (6·1 to 10) | 6·8 (5·4 to 8·2) | 2·9 (2·4 to 3·4) |
| | 2010 | 5·0 (3·6 to 6·9) | 3·4 (2·6 to 4·4) | 6·1 (4·3 to 8·6) | 2·3 (1·8 to 3·1) | 6·5 (4·3 to 9·6) | 7·3 (5·4 to 9·6) | 3·2 (2·5 to 4·1) |
| | 2015 | 3·4 (2·4 to 4·7) | 2·3 (1·7 to 3·1) | 4·2 (2·8 to 5·9) | 1·6 (1·2 to 2·1) | 3·9 (2·5 to 5·9) | 5·6 (3·8 to 7·4) | 2·8 (2·0 to 3·7) |
| | 2020 | 2·2 (1·4 to 3·3) | 1·5 (1·0 to 2·2) | 2·7 (1·7 to 4·1) | 1·1 (0·8 to 1·4) | 2·3 (1·4 to 3·7) | 3·5 (2·1 to 5·3) | 2·1 (1·3 to 3·3) |
| Diagnosis yield (% of new diagnoses among all tests) | 2000 | 4·2 (3·2 to 4·7) | 2·8 (2·1 to 3·3) | 5·3 (4·1 to 5·9) | 2·9 (2·3 to 3·3) | 6·0 (4·6 to 6·8) | 4·8 (3·6 to 5·4) | 2·0 (1·5 to 2·3) |
| | 2005 | 3·8 (3·1 to 4·3) | 2·7 (2·2 to 3·1) | 4·6 (3·8 to 5·2) | 2·2 (1·8 to 2·5) | 5·2 (4·2 to 5·8) | 5·1 (4·2 to 5·7) | 2·4 (2·0 to 2·7) |
| | 2010 | 2·6 (2·1 to 3·0) | 2·0 (1·7 to 2·3) | 3·0 (2·4 to 3·5) | 1·5 (1·2 to 1·7) | 2·9 (2·2 to 3·5) | 3·9 (3·2 to 4·7) | 2·1 (1·7 to 2·6) |
| | 2015 | 1·6 (1·2 to 1·8) | 1·1 (0·9 to 1·3) | 1·9 (1·4 to 2·3) | 1·0 (0·8 to 1·1) | 1·6 (1·1 to 1·9) | 2·4 (1·8 to 3·1) | 1·5 (1·1 to 1·9) |
| | 2020 | 0·9 (0·6 to 1·3) | 0·7 (0·4 to 1·0) | 1·1 (0·7 to 1·5) | 0·7 (0·5 to 0·9) | 0·9 (0·5 to 1·2) | 1·3 (0·8 to 2·1) | 0·9 (0·5 to 1·5) |
| Proportion of new HIV diagnoses among all positive tests (%) | 2000 | 93 (85 to 97) | 93 (86 to 97) | 92 (85 to 97) | 94 (87 to 98) | 91 (83 to 96) | 93 (87 to 97) | 95 (90 to 98) |
| | 2005 | 71 (55 to 86) | 75 (59 to 88) | 70 (53 to 85) | 76 (60 to 89) | 66 (49 to 83) | 75 (61 to 88) | 82 (70 to 92) |
| | 2010 | 52 (36 to 71) | 59 (43 to 77) | 49 (34 to 70) | 66 (49 to 83) | 44 (29 to 66) | 54 (39 to 72) | 66 (49 to 80) |
| | 2015 | 46 (33 to 59) | 49 (36 to 60) | 45 (32 to 59) | 65 (50 to 76) | 41 (27 to 57) | 44 (31 to 55) | 53 (39 to 62) |
| | 2020 | 42 (24 to 57) | 47 (30 to 61) | 40 (23 to 55) | 62 (42 to 75) | 38 (20 to 55) | 38 (23 to 52) | 43 (27 to 58) |

PLHIV: people living with HIV, HTS: HIV testing services. Numbers in parentheses correspond to 95% credible intervals.

*Table S7*: **Progress in HIV testing related outcomes in Eastern Africa stratified by sex and age group, 2000-2020**

| Outcome | Year | Overall | Men | Women | 15-24 years-old | 25-34 years-old | 35-49 years-old | 50-99 years-old |
|---|---|---|---|---|---|---|---|---|
| Proportion of individuals having ever been tested for HIV among overall population (%) | 2000 | 3·3 (2·8 to 3·9) | 2·9 (2·4 to 3·4) | 3·7 (3·1 to 4·4) | 3·3 (2·8 to 3·9) | 4·6 (3·9 to 5·4) | 2·8 (2·4 to 3·4) | 2·1 (1·7 to 2·5) |
| | 2005 | 13 (12 to 14) | 11 (9·9 to 12) | 14 (13 to 15) | 11 (10 to 12) | 17 (16 to 19) | 12 (11 to 13) | 8·6 (7·9 to 9·4) |
| | 2010 | 41 (39 to 42) | 37 (35 to 39) | 44 (43 to 46) | 37 (35 to 39) | 51 (49 to 53) | 41 (40 to 43) | 32 (30 to 34) |
| | 2015 | 53 (52 to 54) | 49 (47 to 50) | 58 (57 to 59) | 41 (40 to 42) | 66 (65 to 67) | 61 (60 to 62) | 49 (48 to 51) |
| | 2020 | 64 (62 to 66) | 59 (57 to 61) | 68 (66 to 71) | 48 (45 to 52) | 74 (73 to 76) | 74 (73 to 76) | 65 (63 to 67) |
| Proportion of PLHIV who know their HIV status (%) | 2000 | 4·8 (4·0 to 5·8) | 4·0 (3·3 to 4·8) | 5·4 (4·4 to 6·5) | 4·1 (3·3 to 4·9) | 6·1 (5·1 to 7·4) | 4·1 (3·4 to 4·9) | 3·0 (2·4 to 3·6) |
| | 2005 | 19 (17 to 21) | 16 (14 to 17) | 21 (19 to 23) | 14 (12 to 16) | 23 (20 to 25) | 18 (16 to 20) | 14 (12 to 15) |
| | 2010 | 56 (53 to 58) | 49 (47 to 51) | 60 (57 to 62) | 44 (41 to 47) | 60 (57 to 63) | 57 (55 to 60) | 53 (50 to 55) |
| | 2015 | 74 (72 to 75) | 67 (65 to 69) | 78 (77 to 80) | 57 (55 to 59) | 74 (72 to 76) | 78 (76 to 79) | 77 (75 to 79) |
| | 2020 | 86 (85 to 88) | 81 (79 to 83) | 90 (89 to 91) | 71 (69 to 74) | 84 (83 to 86) | 89 (88 to 90) | 92 (91 to 93) |
| Time to diagnosis or AIDS-related death, median (year) | 2000 | 11 (10 to 11) | 11 (10 to 11) | 11 (10 to 11) | 12 (12 to 13) | 10 (9·8 to 11) | 9·0 (8·8 to 9·2) | 8·0 (7·8 to 8·0) |
| | 2005 | 7·3 (6·3 to 8·2) | 7·8 (6·9 to 8·5) | 6·9 (5·8 to 7·9) | 7·6 (6·4 to 8·8) | 6·9 (5·9 to 7·8) | 7·3 (6·7 to 7·8) | 6·8 (6·4 to 7·1) |
| | 2010 | 3·2 (2·8 to 3·6) | 4·0 (3·6 to 4·4) | 2·6 (2·3 to 3·0) | 3·1 (2·7 to 3·5) | 2·8 (2·4 to 3·2) | 4·0 (3·6 to 4·4) | 4·2 (3·9 to 4·6) |
| | 2015 | 2·6 (2·4 to 2·9) | 3·4 (3·1 to 3·7) | 2·0 (1·8 to 2·3) | 2·6 (2·3 to 2·9) | 2·2 (2·0 to 2·4) | 3·2 (2·9 to 3·4) | 3·4 (3·2 to 3·7) |
| | 2020 | 2·0 (1·5 to 2·7) | 2·7 (2·0 to 3·5) | 1·5 (1·1 to 2·1) | 2·0 (1·4 to 2·7) | 1·7 (1·3 to 2·3) | 2·4 (1·8 to 3·1) | 2·6 (2·0 to 3·3) |
| Probability of getting tested within 1 year following infection (%) | 2000 | 1·7 (1·4 to 2·1) | 1·5 (1·2 to 1·8) | 1·9 (1·6 to 2·3) | 1·9 (1·5 to 2·3) | 2·1 (1·7 to 2·5) | 1·2 (1·0 to 1·4) | 0·9 (0·8 to 1·1) |
| | 2005 | 5·8 (4·4 to 7·6) | 4·7 (3·6 to 6·1) | 6·5 (4·9 to 8·6) | 6·2 (4·7 to 8·2) | 6·8 (5·2 to 8·9) | 3·8 (2·9 to 5·0) | 3·1 (2·4 to 4·1) |
| | 2010 | 25 (21 to 29) | 18 (15 to 22) | 30 (25 to 35) | 27 (22 to 32) | 29 (24 to 35) | 16 (13 to 19) | 12 (10 to 15) |
| | 2015 | 31 (28 to 35) | 22 (19 to 24) | 38 (34 to 42) | 33 (29 to 36) | 37 (33 to 41) | 21 (19 to 24) | 17 (15 to 19) |
| | 2020 | 40 (29 to 52) | 28 (20 to 39) | 48 (36 to 61) | 41 (30 to 53) | 47 (35 to 60) | 29 (20 to 40) | 24 (17 to 34) |
| Probability of getting tested before reaching a CD4 count lower than 350 cells per µL (%) | 2000 | 14 (11 to 16) | 11 (9·6 to 14) | 15 (12 to 18) | 17 (14 to 20) | 14 (12 to 16) | 7·6 (6·3 to 9·0) | 5·1 (4·2 to 6·2) |
| | 2005 | 35 (29 to 42) | 30 (24 to 36) | 38 (32 to 46) | 43 (36 to 50) | 36 (29 to 43) | 21 (17 to 27) | 15 (12 to 19) |
| | 2010 | 66 (62 to 70) | 58 (54 to 63) | 71 (68 to 75) | 73 (69 to 76) | 71 (67 to 74) | 50 (45 to 54) | 38 (34 to 42) |
| | 2015 | 71 (68 to 74) | 63 (59 to 66) | 77 (74 to 79) | 76 (74 to 79) | 76 (73 to 78) | 58 (54 to 61) | 45 (41 to 49) |
| | 2020 | 77 (70 to 83) | 69 (61 to 77) | 82 (76 to 87) | 81 (74 to 87) | 81 (75 to 86) | 66 (57 to 74) | 55 (45 to 64) |
| Positivity (% of positive test among all tests) | 2000 | 11 (9·6 to 12) | 8·5 (7·6 to 9·3) | 12 (11 to 14) | 5·2 (4·6 to 5·8) | 17 (15 to 18) | 15 (13 to 16) | 5·9 (5·2 to 6·5) |
| | 2005 | 10 (9·1 to 12) | 7·9 (7·2 to 8·9) | 12 (10 to 14) | 4·5 (3·9 to 5·3) | 15 (13 to 18) | 16 (15 to 18) | 6·7 (6·1 to 7·4) |
| | 2010 | 5·6 (4·2 to 7·5) | 4·0 (3·2 to 5·2) | 6·6 (4·8 to 9·1) | 2·8 (2·1 to 3·7) | 7·3 (5·2 to 11) | 9·8 (7·6 to 13) | 4·2 (3·4 to 5·3) |
| | 2015 | 4·0 (3·5 to 4·6) | 3·5 (3·1 to 4·1) | 4·3 (3·7 to 5·0) | 2·4 (2·1 to 2·7) | 4·5 (3·8 to 5·4) | 6·7 (5·8 to 7·9) | 3·7 (2·9 to 4·6) |
| | 2020 | 2·5 (1·9 to 3·3) | 2·3 (1·8 to 2·9) | 2·6 (2·0 to 3·6) | 1·5 (1·2 to 1·9) | 2·7 (2·1 to 3·5) | 4·0 (2·9 to 5·6) | 2·8 (1·9 to 4·2) |
| Diagnosis yield (% of new diagnoses among all tests) | 2000 | 9·7 (8·8 to 11) | 7·9 (7·1 to 8·5) | 11 (10 to 12) | 4·8 (4·3 to 5·2) | 15 (14 to 16) | 14 (12 to 15) | 5·6 (5·0 to 6·1) |
| | 2005 | 7·6 (7·0 to 8·1) | 6·4 (5·9 to 6·9) | 8·5 (7·7 to 9·1) | 3·6 (3·2 to 3·8) | 10 (9·5 to 11) | 13 (12 to 14) | 5·8 (5·3 to 6·4) |
| | 2010 | 2·4 (2·1 to 2·9) | 2·2 (1·9 to 2·7) | 2·6 (2·2 to 3·0) | 1·5 (1·2 to 1·7) | 2·6 (2·1 to 3·0) | 4·8 (4·0 to 5·7) | 2·7 (2·3 to 3·3) |
| | 2015 | 1·7 (1·6 to 1·8) | 1·7 (1·5 to 1·9) | 1·7 (1·5 to 1·8) | 1·3 (1·2 to 1·4) | 1·5 (1·4 to 1·7) | 2·7 (2·4 to 3) | 2·0 (1·7 to 2·3) |
| | 2020 | 1·0 (0·8 to 1·3) | 1·0 (0·8 to 1·2) | 1·0 (0·8 to 1·3) | 0·9 (0·7 to 1·1) | 1·0 (0·8 to 1·2) | 1·4 (1·1 to 1·8) | 1·1 (0·8 to 1·5) |
| Proportion of new HIV diagnoses among all positive tests (%) | 2000 | 91 (85 to 95) | 93 (89 to 96) | 90 (83 to 95) | 92 (87 to 96) | 89 (82 to 94) | 93 (89 to 96) | 95 (92 to 97) |
| | 2005 | 74 (61 to 84) | 81 (71 to 88) | 70 (57 to 82) | 79 (68 to 88) | 67 (54 to 80) | 79 (68 to 87) | 87 (79 to 92) |
| | 2010 | 44 (32 to 59) | 55 (43 to 68) | 39 (28 to 55) | 53 (39 to 68) | 35 (24 to 50) | 49 (38 to 62) | 65 (52 to 76) |
| | 2015 | 42 (37 to 48) | 49 (43 to 55) | 39 (33 to 45) | 55 (49 to 62) | 34 (29 to 41) | 40 (34 to 46) | 53 (43 to 65) |
| | 2020 | 41 (30 to 52) | 44 (34 to 54) | 39 (28 to 52) | 59 (49 to 70) | 35 (26 to 47) | 34 (24 to 46) | 38 (25 to 56) |

PLHIV: people living with HIV, HTS: HIV testing services. Numbers in parentheses correspond to 95% credible intervals.

*Table S8*: **Progress in HIV testing related outcomes in Southern Africa stratified by sex and age group, 2000-2020**

| Outcome | Year | Overall | Men | Women | 15-24 years-old | 25-34 years-old | 35-49 years-old | 50-99 years-old |
|---|---|---|---|---|---|---|---|---|
| Proportion of individuals having ever been tested for HIV among overall population (%) | 2000 | 10 (8·3 to 12·9) | 8·5 (6·7 to 11) | 12 (9·8 to 15) | 8·8 (6·9 to 11) | 14 (11 to 17) | 11 (8·5 to 13) | 8·4 (6·8 to 11) |
| | 2005 | 30 (29 to 32) | 25 (23 to 27) | 34 (33 to 36) | 22 (21 to 24) | 39 (37 to 41) | 34 (32 to 36) | 27 (26 to 29) |
| | 2010 | 59 (58 to 61) | 52 (50 to 54) | 65 (64 to 67) | 44 (42 to 46) | 69 (67 to 70) | 68 (66 to 70) | 58 (56 to 61) |
| | 2015 | 75 (74 to 75) | 68 (67 to 69) | 80 (79 to 81) | 53 (52 to 54) | 82 (81 to 83) | 86 (85 to 87) | 79 (77 to 81) |
| | 2020 | 85 (83 to 88) | 80 (77 to 84) | 90 (87 to 92) | 62 (57 to 70) | 90 (88 to 93) | 95 (93 to 96) | 92 (90 to 93) |
| Proportion of PLHIV who know their HIV status (%) | 2000 | 9·3 (7·5 to 12) | 6·9 (5·4 to 8·7) | 11 (8·7 to 14) | 7·0 (5·5 to 8·9) | 11 (9·1 to 14) | 8·9 (7·2 to 11) | 7·8 (6·4 to 9·5) |
| | 2005 | 27 (25 to 30) | 21 (19 to 24) | 31 (28 to 34) | 18 (16 to 20) | 32 (30 to 35) | 28 (26 to 31) | 25 (23 to 27) |
| | 2010 | 60 (58 to 63) | 51 (48 to 54) | 66 (63 to 68) | 41 (38 to 44) | 65 (63 to 68) | 64 (61 to 67) | 59 (56 to 62) |
| | 2015 | 79 (77 to 80) | 72 (70 to 74) | 82 (81 to 84) | 55 (52 to 57) | 80 (78 to 81) | 84 (82 to 85) | 81 (79 to 83) |
| | 2020 | 90 (88 to 92) | 87 (84 to 90) | 92 (90 to 94) | 70 (65 to 76) | 89 (87 to 92) | 93 (92 to 95) | 93 (91 to 95) |
| Time to diagnosis or AIDS-related death, median (year) | 2000 | 7·7 (7·0 to 8·4) | 8·2 (7·6 to 8·7) | 7·4 (6·6 to 8·1) | 8·5 (7·7 to 9·3) | 7·1 (6·4 to 7·8) | 6·8 (6·3 to 7·2) | 6·4 (6·1 to 6·6) |
| | 2005 | 5·9 (5·1 to 6·6) | 6·5 (5·8 to 7·2) | 5·4 (4·6 to 6·2) | 6·4 (5·5 to 7·2) | 5·3 (4·6 to 6·1) | 5·6 (4·9 to 6·2) | 5·4 (4·9 to 5·8) |
| | 2010 | 2·8 (2·5 to 3·1) | 3·5 (3·2 to 3·8) | 2·4 (2·1 to 2·7) | 3·1 (2·8 to 3·5) | 2·4 (2·1 to 2·6) | 2·7 (2·4 to 3·0) | 3·1 (2·8 to 3·4) |
| | 2015 | 2·2 (1·9 to 2·4) | 2·7 (2·4 to 3·0) | 1·8 (1·6 to 2·1) | 2·5 (2·2 to 2·7) | 1·8 (1·6 to 2·0) | 2·0 (1·8 to 2·3) | 2·4 (2·1 to 2·6) |
| | 2020 | 1·5 (0·9 to 2·3) | 1·8 (1·1 to 2·7) | 1·3 (0·8 to 2·0) | 1·7 (1·0 to 2·7) | 1·2 (0·7 to 1·9) | 1·4 (0·9 to 2·2) | 1·7 (1·0 to 2·5) |
| Probability of getting tested within 1 year following infection (%) | 2000 | 4·2 (3·3 to 5·3) | 3·1 (2·4 to 3·9) | 4·9 (3·9 to 6·3) | 3·9 (3·0 to 5·0) | 5·1 (4·0 to 6·5) | 3·6 (2·8 to 4·6) | 3·1 (2·5 to 3·9) |
| | 2005 | 7·5 (5·8 to 9·7) | 5·7 (4·4 to 7·4) | 8·7 (6·6 to 11) | 7·1 (5·5 to 9·2) | 9·2 (7·1 to 12) | 6·4 (4·9 to 8·3) | 5·6 (4·3 to 7·3) |
| | 2010 | 23 (20 to 26) | 17 (15 to 19) | 27 (23 to 31) | 20 (18 to 23) | 29 (25 to 33) | 21 (18 to 25) | 17 (14 to 20) |
| | 2015 | 28 (25 to 32) | 22 (19 to 25) | 33 (29 to 37) | 25 (22 to 29) | 35 (31 to 39) | 27 (23 to 31) | 22 (19 to 25) |
| | 2020 | 40 (26 to 60) | 32 (20 to 50) | 44 (29 to 65) | 36 (23 to 56) | 49 (33 to 69) | 38 (24 to 57) | 32 (20 to 49) |
| Probability of getting tested before reaching a CD4 count lower than 350 cells per µL (%) | 2000 | 30 (25 to 35) | 23 (19 to 28) | 34 (29 to 40) | 34 (29 to 40) | 31 (25 to 37) | 20 (17 to 25) | 15 (12 to 18) |
| | 2005 | 43 (37 to 50) | 36 (30 to 42) | 48 (41 to 55) | 49 (43 to 55) | 45 (39 to 52) | 32 (26 to 39) | 24 (20 to 30) |
| | 2010 | 69 (66 to 72) | 61 (58 to 65) | 74 (71 to 77) | 72 (69 to 74) | 74 (71 to 77) | 62 (58 to 66) | 49 (45 to 53) |
| | 2015 | 75 (72 to 77) | 68 (65 to 71) | 79 (76 to 81) | 76 (74 to 79) | 79 (77 to 82) | 69 (65 to 72) | 57 (53 to 61) |
| | 2020 | 81 (72 to 89) | 77 (65 to 86) | 84 (75 to 91) | 83 (74 to 90) | 86 (78 to 92) | 78 (66 to 86) | 67 (53 to 78) |
| Positivity (% of positive test among all tests) | 2000 | 15 (13 to 19) | 12 (10 to 14) | 18 (15 to 22) | 14 (13 to 16) | 24 (21 to 29) | 14 (12 to 16) | 3·3 (2·7 to 4·1) |
| | 2005 | 20 (16 to 26) | 15 (13 to 19) | 23 (19 to 31) | 14 (12 to 17) | 31 (26 to 41) | 21 (18 to 27) | 5·3 (4·5 to 6·8) |
| | 2010 | 13 (9·0 to 22) | 10 (7·7 to 15) | 15 (9·9 to 25) | 9·5 (7·4 to 14) | 19 (12 to 32) | 16 (11 to 25) | 4·1 (3·0 to 5·9) |
| | 2015 | 9·2 (7·1 to 13) | 7·6 (6·2 to 9·8) | 10 (7·7 to 14) | 6·5 (5·5 to 8·0) | 12 (8·9 to 17) | 13 (9·9 to 17) | 3·5 (2·8 to 5·0) |
| | 2020 | 5·5 (3·8 to 8·4) | 5·2 (3·6 to 7·5) | 5·8 (3·9 to 9·0) | 3·5 (2·7 to 4·6) | 6·3 (4·2 to 9·8) | 8·4 (5·5 to 13) | 3·0 (2·0 to 5·4) |
| Diagnosis yield (% of new diagnoses among all tests) | 2000 | 13 (12 to 14) | 11 (9·4 to 11) | 15 (14 to 16) | 13 (12 to 14) | 20 (18 to 22) | 12 (11 to 13) | 2·8 (2·5 to 3·1) |
| | 2005 | 14 (12 to 15) | 11 (11 to 12) | 15 (13 to 16) | 11 (10 to 12) | 20 (16 to 22) | 15 (13 to 16) | 4·1 (3·8 to 4·4) |
| | 2010 | 6·9 (6·2 to 7·5) | 6·5 (5·9 to 7·1) | 7·2 (6·4 to 7·9) | 6·6 (6·1 to 7·2) | 8·3 (7·1 to 9·3) | 8·5 (7·6 to 9·4) | 2·7 (2·4 to 3·0) |
| | 2015 | 4·4 (4·1 to 4·6) | 4·3 (3·9 to 4·6) | 4·4 (4·1 to 4·7) | 4·5 (4·2 to 4·8) | 4·8 (4·4 to 5·2) | 5·5 (5·1 to 5·9) | 2·0 (1·9 to 2·2) |
| | 2020 | 2·2 (1·6 to 2·9) | 2·2 (1·5 to 3·0) | 2·2 (1·6 to 2·8) | 2·3 (1·7 to 2·9) | 2·2 (1·5 to 2·9) | 2·8 (1·9 to 3·8) | 1·2 (0·9 to 1·8) |
| Proportion of new HIV diagnoses among all positive tests (%) | 2000 | 87 (70 to 97) | 91 (77 to 98) | 86 (67 to 97) | 91 (78 to 98) | 85 (65 to 97) | 88 (72 to 98) | 87 (70 to 98) |
| | 2005 | 70 (48 to 87) | 78 (58 to 91) | 67 (44 to 86) | 81 (61 to 93) | 64 (40 to 84) | 72 (50 to 88) | 78 (58 to 90) |
| | 2010 | 53 (31 to 76) | 64 (41 to 82) | 49 (27 to 73) | 70 (46 to 88) | 44 (23 to 70) | 54 (33 to 75) | 67 (46 to 84) |
| | 2015 | 47 (34 to 60) | 56 (43 to 68) | 43 (31 to 57) | 70 (57 to 80) | 40 (28 to 54) | 43 (31 to 56) | 58 (41 to 73) |
| | 2020 | 39 (25 to 55) | 42 (28 to 56) | 37 (23 to 55) | 66 (50 to 79) | 35 (22 to 51) | 34 (20 to 49) | 41 (22 to 61) |

PLHIV: people living with HIV, HTS: HIV testing services. Numbers in parentheses correspond to 95% credible intervals.
